# *“*Validation of a novel ordinal endpoint for an adaptive platform trial in paediatric haematopoietic stem cell transplant*”*

**DOI:** 10.1101/2025.03.21.25324374

**Authors:** Hannah Walker, Chris J Selman, Li-yin Goh, Katherine J Lee, Kristene Rombaldo, Lorna McLeman, Peter Summers, Julian Stolper, Diane Hanna, David Hughes, Stacie S Wang, Claudia Toro, Deborah Meyran, Beth Williams, Roxanne Dyas, Lori Chait Rubinek, Kaitlyn Taylor, Tom Snelling, Theresa Cole, Amanda Gwee, Anneke Grobler, Rachel Conyers

**Author notes:** Designates co-first authorship (Hannah Walker, Chris J Selman). **Correspondence:** A/Prof Rachel Conyers, Cancer Therapies Group, Stem Cell Medicine Theme Murdoch Children’s Research Institute, 50 Flemington Rd, Parkville, Victoria, Australia 3052. **Author Contributions:** All authors have approved the final version of the manuscript for publication.

## Abstract

**Background:** Haematopoietic stem cell transplant (HCT) is a curative therapy for various paediatric conditions but is associated with significant morbidity and mortality, particularly in children requiring intensive care, facing delayed immune reconstitution or prolonged viral reactivation. Due to the rarity and heterogeneity of paediatric HCT, traditional randomised controlled trials are challenging. Adaptive platform trials (APTs), which evaluate multiple interventions across multiple subgroups, offer a solution, but typically rely on a shared short-term primary outcome that is relevant to clinicians, patients and families and can be used for all interventions/subgroups. No such outcome currently exists in paediatric HCT. In this article we propose and validate four novel ordinal outcomes to assess HCT-related morbidity and mortality within the first 100 days post-transplant for use in ***A****n adaptive platform trial **D**esigned to **I**mprove the **CO**mplications, cost-effectiveness and health **<u>O</u>**utcomes for children receiving a stem cell **T**ransplant (BANDICOOT)* APT, designed.

**Methods:** The proposed outcomes were validated using real-world data from *n*=202 paediatric patients who underwent allogeneic HCT. The validation process included examining the distribution of patients across outcome categories, assessing the association with key long-term outcomes post HCT, and evaluating whether exposures with known efficacy had the expected associations with the proposed endpoints and whether the proportional odds assumption used in the analysis is likely to be reasonable. We also sought feedback on the outcomes from clinicians and family representatives.

**Results:** The results showed strong associations between each ordinal endpoint and long-term HCT complications, including relapse, chronic-graft-versus-host disease, and death. Associations with key exposures (e.g. donor type and positive minimal residual disease pre HCT) were mostly in the expected direction. Moreover, expert feedback from clinicians and family representatives indicates that one of the proposed endpoints, which incorporates viral-related patient states and single/multi-organ support days, was both feasible and relevant for use in BANDICOOT.

**Conclusions:** The selected ordinal endpoint provides a robust and clinically applicable framework for evaluating interventions in paediatric HCT that offers broad applicability across various HCT outcomes.

## INTRODUCTION

Haematopoietic stem cell transplant (HCT) is a potentially curative therapy for a range of rare paediatric medical conditions, including malignant diseases such as leukemia, inborn errors of immunity and metabolic disorders ^1^. Allogeneic HCT involves the administration of high dose chemotherapy, and/or radiation and serotherapy (for T-cell depletion), followed by an infusion of healthy haematopoietic stem cells from a carefully selected donor. Despite the curative potential of allogeneic HCT, it carries with it significant treatment related morbidity and mortality ^2–7^. The highest risk cohorts are paediatric patients requiring intensive care level support, patients who fail to achieve early immune reconstitution and patients who experience prolonged viral reactivation ^2–7^. Unfortunately, due to several factors, which include the rarity of paediatric transplant and the heterogeneity of its disease groups, there is a lack of randomised clinical trials (RCTs) of interventions to improve HCT related morbidity and mortality ^8–11^.

Traditional two-arm RCTs are not well suited to HCT given the heterogeneity of the patients, the variety of interventions at different stages of the process and the limited participant pool. More flexible trial designs such as a platform trial are better suited to a HCT population, as they allow for the evaluation of multiple treatment modalities (or ‘domains’) simultaneously to answer research questions across heterogeneous populations, offer much greater flexibility and may be of more use in this context. For example, platform trials can include adaptations, a so-called adaptive platform trial (APT), to increase trial efficiency, such as the inclusion of stopping rules for efficacy and/or futility. Another benefit of platform designs is the ability to borrow information between subgroups of patients, which can further increase efficiency. These efficiency gains mean the expected sample size of the trial is decreased compared to traditional “fixed” design, making such designs appealing in HCT given its rarity. APTs are particularly appealing in the setting of paediatric HCT because of the clearly defined phases of HCT; e.g. pre HCT (preparation of the patient and conditioning phase) and post HCT (complications), each of which provides a potential point in an APT for enrolment and randomisation.

In most APTs, all domains share a common primary outcome. To be useful for guiding adaptations, this endpoint must occur in a relatively short time frame post-randomisation so that adaptations can be implemented into the platform without delay. Whilst short-term endpoints are often used as the first timepoint of evaluation in paediatric HCT RCTs (e.g. measured at day 100 such as time to develop acute graft versus host disease), the primary outcome has often focused on a time to event endpoint (e.g. event free or overall survival) that is measured at five years post HCT. Such endpoints fail to take into account other patient related events that may be of clinical interest and do not encapsulate other endpoints that are of interest to consumers and clinicians alike. The need for a short-term outcome, that captures mortality as well morbidity and is relevant to consumers, demanded the need to develop a novel outcome to be used in a platform trial in HCT.

One approach to combine multiple health states into a single endpoint is to use an ordinal outcome. An ordinal outcome is an endpoint that is categorical in nature and assigns patients a ranking, e.g. from level 1 representing a “favourable” outcome to level 5 representing an “unfavourable” outcome, where proximate categories are not necessarily evenly spaced. An ordinal endpoint is appealing since it can combine various efficacy and safety endpoints in a single hierarchy and can capture different patient states that come from heterogeneous populations. Compared to dichotomised ordinal outcomes, ordinal outcomes can increase statistical power to detect a treatment effect in a RCT, although their use and analysis may be less familiar to clinicians and statisticians alike.

The use of ordinal outcomes in RCTs has risen to prominence particularly since the COVID-19 pandemic which led to the derivation of the Clinical Progression Scale^14^ (and its various adaptations), developed by the World Health Organisation. The increased uptake of ordinal outcomes during the pandemic also coincided with the need for efficient trial designs that could expedite conclusions, in particular APTs. Ordinal outcomes have been used in numerous APTs, including the randomised embedded multifactorial adaptive platform for community acquired pneumonia (REMAP-CAP)^15^ that evaluated the effectiveness of various treatments for patients with CAP, and the Australasian COVID-19 Trial (ASCOT)^16^ that aimed to provide evidence-based guidance on how best to treat COVID-19 and improve patient outcomes.

When an ordinal outcome is used in an RCT, there are many possible target estimands and statistical approaches available to analyse the outcome, with the proportional odds ratio (OR) estimated using a proportional odds model^17^ most commonly used^18^. In this context, the proportional OR represents the odds of a more favourable outcome in the intervention arm compared to a control arm representing the ‘global’ average treatment effect across the distribution of the ordinal outcome^19^. The proportional odds model relies on the assumption that the intervention exerts the same effect across each binary split of the ordinal outcome. This assumption may or may not be true in practice. If this assumption does not hold, the proportional OR can potentially mask important treatment effects. There are various methods that can be used to assess the proportional odds assumption, such as plotting the differences in predicted logits between different categories of the ordinal outcome or using a formal hypothesis testing approach such as the Brant test or likelihood ratio test.^20,21^ However, such statistical approaches tend to lack statistical power^22^. An informal and potentially more useful approach than formal statistical tests is to estimate and graphically compare the OR for each binary split of the ordinal scale and its confidence interval (CI) which can be used to determine whether the point estimates are of similar magnitude and its variability.

Our group recently developed two novel short-term ordinal outcomes for use in an APT in HCT, BANDICOOT, “*An adaptive platform trial Designed to Improve the Complications, cost-effectiveness and health <u>O</u>utcomes for children receiving a stem cell Transplant”*. These outcomes capture HCT related morbidity and mortality occurring in the first 100 days post HCT ^23^. We recently published the process for developing these outcomes^23^. In the current manuscript, we aim to validate these proposed primary outcomes, as well as two additional versions which arose through the validation process (four in total) using real-world data. We aim to address four specific objectives in this validation paper:

1. To describe the distribution of potential patients across the categories of the proposed ordinal outcomes in a similar population to the planned APT population. The ideal scenario is a balanced distribution of patients spread across all categories, avoiding categories with few patients.
2. To ascertain whether the proposed ordinal endpoints are predictive of long term (five year) outcomes traditionally used in RCTs in HCT (e.g. relapse, overall survival, chronic graft versus host disease)^2,24^.
3. To determine whether exposures with known efficacy demonstrate an effect on the proposed ordinal endpoints and whether the proportional odds assumption holds for these exposures.
4. To obtain expert and consumer feedback on the feasibility of collecting data for the proposed endpoints to inform the final choice for the APT.

## METHODS

The four ordinal endpoints validated in this paper are summarised in Table 1. The process and rationale behind the Endpoints 1 and 2 are described in our recent publication.^23^ Endpoint 1 is comprised of five categories related to minor and major organ support and hospital admitted days. Endpoints 2 and 3 also consisted of five categories but extended the definition of the first endpoint to incorporate organ support in addition to immune reconstitution and viral reactivation. The difference between Endpoint 2 and 3 is the definition of organ support as minor or major organ support in Endpoint 2 (Supplementary Table 1) compared to single or multi-organ support in Endpoint 3 (Supplementary Table 2). Endpoints 4 was included in this validation since preliminary validation work showed a high concentration of patients in some categories and no patients being assigned to other categories. This raised a question as to whether an eight-category outcome that comprises categories from Endpoints 1 and 2 would have better statistical properties than the five-category outcomes, resulting in Endpoint 4 which combines immune reconstitution <u>and</u> viral reactivation into a single category and sub-divides the CD4+ count cut-offs.

**Table 1:**
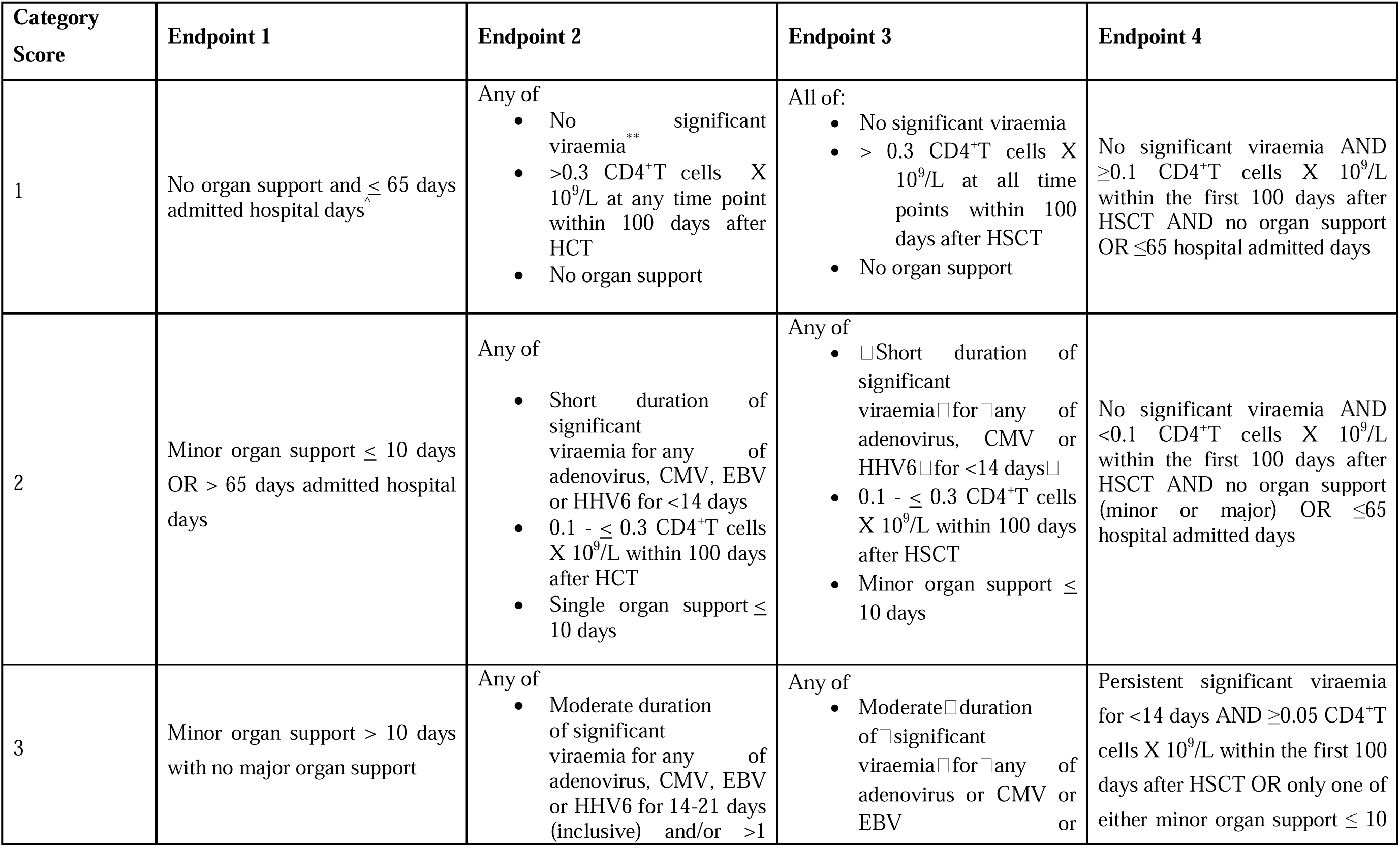
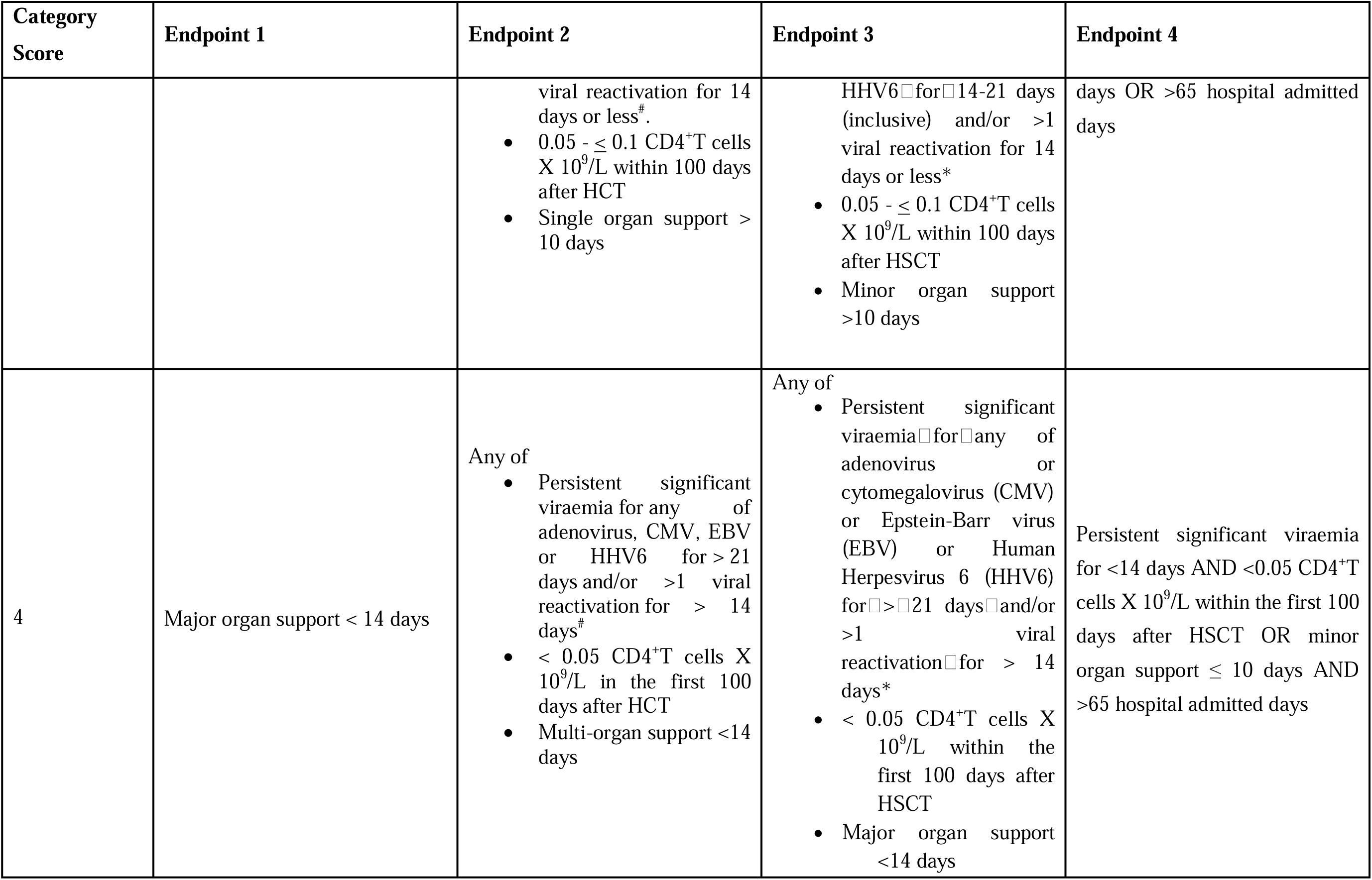
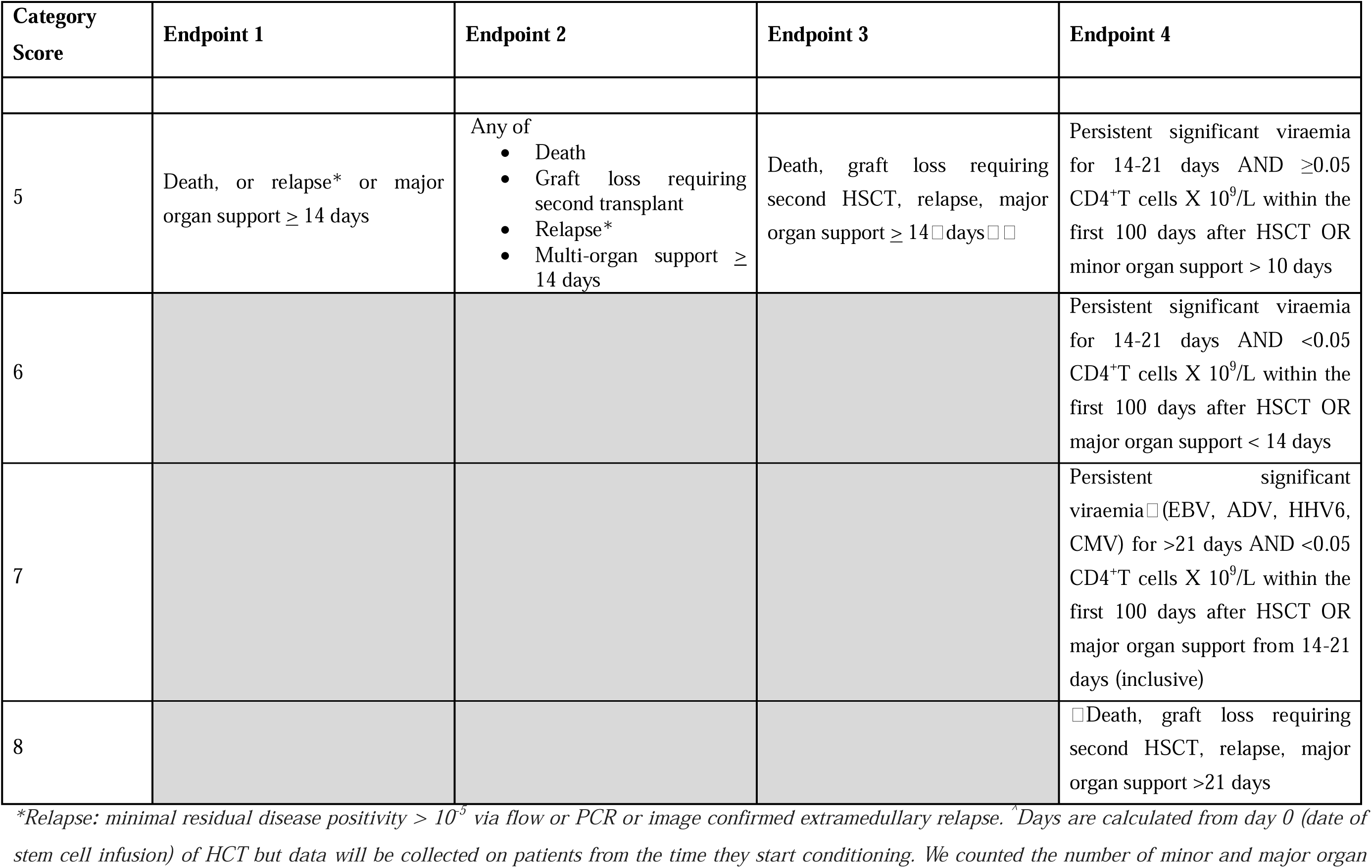
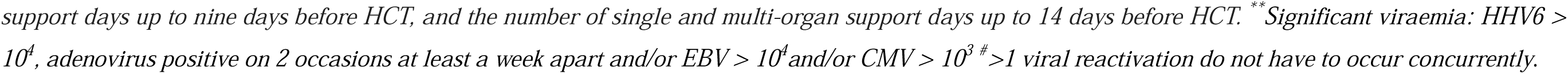
Proposed ordinal endpoints.

| Category Score | Endpoint 1 | Endpoint 2 | Endpoint 3 | Endpoint 4 |
| --- | --- | --- | --- | --- |
| 1 | No organ support and $\leq 65$ days admitted hospital days | Any of<br>No significant viraemia**<br>$>0.3$ CD4<sup>+</sup>T cells X $10^9/L$ at any time point within 100 days after HCT<br>No organ support | All of:<br>No significant viraemia<br>$> 0.3$ CD4<sup>+</sup>T cells X $10^9/L$ at all time points within 100 days after HSCT<br>No organ support | No significant viraemia AND $\geq 0.1$ CD4<sup>+</sup>T cells X $10^9/L$ within the first 100 days after HSCT AND no organ support OR $\leq 65$ hospital admitted days |
| 2 | Minor organ support $\leq 10$ days OR $> 65$ days admitted hospital days | Any of<br>Short duration of significant viraemia for any of adenovirus, CMV, EBV or HHV6 for $<14$ days<br>$0.1 - \leq 0.3$ CD4<sup>+</sup>T cells X $10^9/L$ within 100 days after HCT<br>Single organ support $\leq 10$ days | Any of<br>Short duration of significant viraemia for any of adenovirus, CMV or HHV6 for $<14$ days<br>$0.1 - \leq 0.3$ CD4<sup>+</sup>T cells X $10^9/L$ within 100 days after HSCT<br>Minor organ support $\leq 10$ days | No significant viraemia AND $<0.1$ CD4<sup>+</sup>T cells X $10^9/L$ within the first 100 days after HSCT AND no organ support (minor or major) OR $\leq 65$ hospital admitted days |
| 3 | Minor organ support $> 10$ days with no major organ support | Any of<br>Moderate duration of significant viraemia for any of adenovirus, CMV, EBV or HHV6 for 14-21 days (inclusive) and/or $>1$ | Any of<br>Moderate duration of significant viraemia for any of adenovirus or CMV or EBV | Persistent significant viraemia for $<14$ days AND $\geq 0.05$ CD4<sup>+</sup>T cells X $10^9/L$ within the first 100 days after HSCT OR only one of either minor organ support $\leq 10$ |
|  |  | viral reactivation for 14 days or less<sup>#</sup>.<br>• 0.05 - ≤ 0.1 CD4<sup>+</sup>T cells X 10<sup>9</sup>/L within 100 days after HCT<br>• Single organ support > 10 days | HHV6 for 14-21 days (inclusive) and/or >1 viral reactivation for 14 days or less*<br>• 0.05 - ≤ 0.1 CD4<sup>+</sup>T cells X 10<sup>9</sup>/L within 100 days after HSCT<br>• Minor organ support >10 days | days OR >65 hospital admitted days |
| 4 | Major organ support < 14 days | Any of<br>• Persistent significant viraemia for any of adenovirus, CMV, EBV or HHV6 for > 21 days and/or >1 viral reactivation for > 14 days<sup>#</sup><br>• < 0.05 CD4<sup>+</sup>T cells X 10<sup>9</sup>/L in the first 100 days after HCT<br>• Multi-organ support <14 days | Any of<br>• Persistent significant viraemia for any of adenovirus or cytomegalovirus (CMV) or Epstein-Barr virus (EBV) or Human Herpesvirus 6 (HHV6) for > 21 days and/or >1 viral reactivation for > 14 days*<br>• < 0.05 CD4<sup>+</sup>T cells X 10<sup>9</sup>/L within the first 100 days after HSCT<br>• Major organ support <14 days | Persistent significant viraemia for <14 days AND <0.05 CD4<sup>+</sup>T cells X 10<sup>9</sup>/L within the first 100 days after HSCT OR minor organ support ≤ 10 days AND >65 hospital admitted days |
| 5 | Death, or relapse* or major organ support ≥ 14 days | Any of<br>• Death<br>• Graft loss requiring second transplant<br>• Relapse*<br>• Multi-organ support ≥ 14 days | Death, graft loss requiring second HSCT, relapse, major organ support ≥ 14 days | Persistent significant viraemia for 14-21 days AND $\geq 0.05$ CD4<sup>+</sup>T cells X 10<sup>9</sup>/L within the first 100 days after HSCT OR minor organ support > 10 days |
| 6 |  |  |  | Persistent significant viraemia for 14-21 days AND <0.05 CD4<sup>+</sup>T cells X 10<sup>9</sup>/L within the first 100 days after HSCT OR major organ support < 14 days |
| 7 |  |  |  | Persistent significant viraemia (EBV, ADV, HHV6, CMV) for >21 days AND <0.05 CD4<sup>+</sup>T cells X 10<sup>9</sup>/L within the first 100 days after HSCT OR major organ support from 14-21 days (inclusive) |
| 8 |  |  |  | Death, graft loss requiring second HSCT, relapse, major organ support >21 days |

\*Relapse: minimal residual disease positivity $> 10^{-5}$ via flow or PCR or image confirmed extramedullary relapse. ^ Days are calculated from day 0 (date of stem cell infusion) of HCT but data will be collected on patients from the time they start conditioning. We counted the number of minor and major organ
*support days up to nine days before HCT, and the number of single and multi-organ support days up to 14 days before HCT. \*\*Significant viraemia: HHV6 > 10<sup>4</sup>, adenovirus positive on 2 occasions at least a week apart and/or EBV > 10<sup>4</sup> and/or CMV > 10<sup>3</sup> #>1 viral reactivation do not have to occur concurrently.*

### Data extraction

We used data from the BILBY database supplemented with medical records to validate the endpoints. The BILBY database consists of 243 patients who underwent allogeneic HCT at either the Royal Children’s Hospital (RCH) in Melbourne or Perth Children’s Hospital between 2016 and 2022, with follow-up ceasing in May 2023. Ethics approval for the collection of deidentified patient information was obtained from the local ethics committee; HREC/91777. The BILBY database included over 700 data fields including deidentified demographic data, transplant episode-specific data, data on various exposures with known efficacy on outcomes post-transplant, and transplant long-term outcomes including graft versus host disease (GVHD), transplant associated thrombotic microangiopathy (TA-TMA), relapse, graft failure, need for a second HCT and survival. For the validation, we included patients who were admitted to RCH and were <18 years old (*n* = 202) and considered the first HCT only Patients from Perth Children’s Hospital were excluded as they do not have an EMR to facilitate additional data extraction required for the validation. The Centre for Health Analytics (CHA) at the RCH extracted additional data required for the derivation of the ordinal endpoints that were not collected in the BILBY database from the EMR. The data required for extraction spanned from 14 days prior to the patient receiving a HCT through to 100-days following HCT. The data points extracted by CHA included maximum CD4+ count in the first 100 days post HCT, plasma/blood viral polymerase chain reaction (PCR) results including titre, organ support status and data related to hospital admitted days. Plasma viral PCR for Epstein Barr virus (EBV), Human herpes virus 6 (HHV-6), Cytomegalovirus (CMV) and Adenovirus are routinely performed weekly in all patients at RCH, increasing to twice weekly in patients who have viral reactivation. We merged the extracted data from the EMR with data from the BILBY database to proceed with the validation.

### Statistical methods

Baseline characteristics of the dataset are summarised using counts and percentages for categorical variables and means and standard deviations for continuous variables. We describe the distribution of patients across the categories of the proposed endpoints by reporting the number and percentage of patients that fall in each category of each endpoint (as well as the number of patients with missing endpoint data) and visualise the distribution using bar charts.

We assessed whether the four proposed primary endpoints were associated with long-term outcomes by using the proposed endpoint as the exposure in a survival analysis. The long-term outcomes of interest were time to relapse, secondary graft failure requiring a second HCT, TA-TMA, chronic GVHD, and death from time of HCT. Kaplan-Meier estimates of survival were plotted by each category of the 4 proposed endpoints. We estimated the hazard ratio (HR) of each category of each endpoint compared to the lowest category using a Cox proportional hazards model censoring patients who did not experience the event within the 5-year follow-up period or beyond 31 May 2023 (whichever came first), or at the time of death (for all long-term outcomes apart from death). The proportional hazards assumption was assessed visually by comparing the log(-log) of the survival function for different categories of the ordinal endpoint over time in conjunction with the results of a global statistical test based on the Schoenfeld residuals. We also summarised the proportion of patients who had each of these long-term outcomes in each category of each endpoint.

To determine whether the proposed ordinal endpoints can distinguish the effect of historic exposures with known efficacy, we investigated the effect of the following exposures using each of the ordinal endpoints: transplant donor source (matched sibling, matched unrelated and haploidentical donors with separate analyses in patients with malignant and non-malignant indications); whether the patient was a high-risk cytomegalovirus (CMV) mismatch (donor CMV negative, D-, recipient CMV positive R+) compared to low risk CMV mismatch or matched donor recipient (where a patient was considered high risk if the recipient is negative but the donor is positive); whether the patient received a cord blood transplant vs other (bone marrow or peripheral blood stem cell transplant), and among those with a B-acute lymphoblastic leukemia (ALL), we compared those with and without positive minimal residual disease (MRD) within four weeks of HCT. We calculated the effects of each of these exposures on the four proposed endpoints by estimating a proportional OR using a proportional odds model. Within these models we adjusted for potential confounders for each exposure where possible; which are noted in the footnote of Table 5. We reverse coded the ordinal outcome categories for this analysis so that a proportional OR > 1 is interpreted as the exposed group having higher odds of a more favourable outcome. This was done so that the interpretation of the exposure effect was based on a positive outcome to reduce negative perceptions. For each analysis we assessed the proportional odds assumption by calculating an OR for each binary cut-point by fitting a logistic regression model to the binary outcome (including adjustment for confounders) as well as Grotta bar charts, which were also used to visualise the (unadjusted) cumulative probabilities across the categories of the outcome by exposure group. The results from the logistic regression models were presented as forest plots of the OR with corresponding 95% confidence interval (CI) for each of the binary splits.

### Obtaining expert and consumer feedback

Once the statistical validation process had been conducted, we presented the results to a focus group of expert paediatric HCT physicians, nurses and consumer representatives (families of children who underwent HCT) who were not directly involved in the design of the ordinal endpoints. We asked the group for feedback on a.) the feasibility of collecting the data prospectively to calculate the primary ordinal endpoints, b.) the applicability of the ordinal endpoints to outcomes they consider most important for patients and families, and c) whether each endpoint was feasible (yes/no) and applicable to consumers (yes/no). We describe the general consensus for a) and b), and the proportion stating yes for feasibility and applicability for each endpoint for c).

## RESULTS

The BILBY database identified 202 patients who fit the inclusion criteria. The majority of patients were less than 13 years of age (151, 75%) and had a malignant indication (118, 58%) (Table 2). Matched unrelated donor was the most common type of donor (81, 40%) followed by matched sibling donor (68, 34%) and haploidentical donors (53, 26%). Thirty-one (16%) patients were considered to have been a high risk CMV mismatch. Median follow-up time for patients was 3.3 years post HCT.

**Table 2:** Summary of baseline characteristics at time of transplant (N=202)

| Characteristic | Number (Percentage) or Mean (SD) |
| --- | --- |
| **Age (years)** | Mean = 7.9 (SD = 5.6) |
| 13 years old or less | 151 (74.8%) |
| >13 years old | 51 (25.2%) |
| **Gender** |  |
| Female | 84 (41.6%) |
| Male | 118 (58.4%) |
| **Type of donor** |  |
| Matched sibling | 68 (33.7%) |
| Matched unrelated donor | 81 (40.1%) |
| Haploidentical donor | 53 (26.2%) |
| **Indication for HCT** |  |
| Malignant | 118 (58.4%) |
| Non-malignant | 84 (41.6%) |
| **Conditioning regimen** |  |
| Myeloablative conditioning | 151 (75.5%) |
| Reduced intensity conditioning | 49 (24.5%) |
| Missing | 2 observations |
| **High risk CMV mismatch** |  |
| Not mismatched | 169 (84.5%) |
| Mismatched (D-/R+) | 31 (15.5%) |
| Missing | 2 observations |
| **Type of transplant** |  |
| Peripheral blood or bone marrow transplant | 187 (92.6%) |
| Cord blood transplant | 15 (7.4%) |

HCT = haematopoietic stem cell transplant; SD = standard deviation.

### Distribution of patients across categories of the proposed endpoints

The number and proportion of patients in each category of the proposed endpoints are shown in Figure 1. There was one patient with unavailable hospital admission data which resulted in missing data for Endpoints 1 and 4. The distribution of patients was fairly evenly spread across the categories for Endpoints 2 and 3 (Figure 1), with slightly higher probabilities of being in the second and fourth categories for each. In contrast, most individuals fell in the ‘best’ health states (the first and second categories) for Endpoints 1 and 3 and no patients were classified in the third category of Endpoint 1, or in the first category of Endpoint 4.

**Figure 1:**
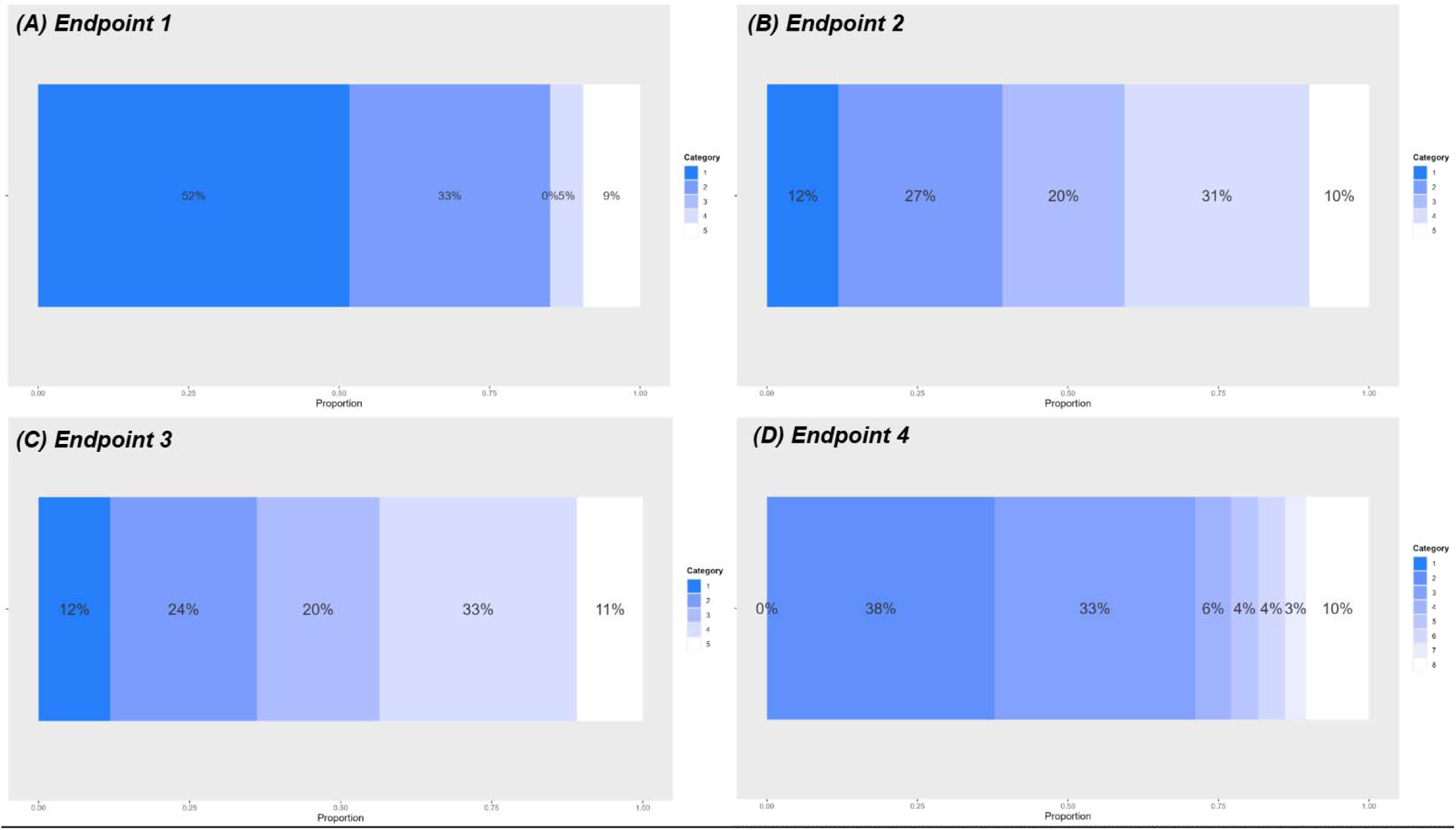
Stacked bar chart for the proposed ordinal endpoints.

### Association between proposed endpoints and long-term outcomes

Within the five years of follow-up (median follow-up time 3.3 years), 21/117 (18.0%) of patients experienced relapse among those with malignant indications (n=117), 6/202 (3.0%) required a second HCT due to secondary graft failure, 10/202 (5.0%) received treatment for TA-TMA, 22/202 (10.9%) had cGVHD, and 37/202 (18.4%) died. All four proposed endpoints were strongly associated with death, with patients classified in higher categories at higher risk of death compared to those in the first (most healthy) category (all HRs > 1), and HRs increasing in magnitude as the category increased, albeit with considerable uncertainty (Table 4 and Figure 2). The 4 endpoints were also strongly associated with other long-term outcomes of time to initiating treatment for TA-TMA, cGVHD, and relapse with the exception of Endpoints 2 and 3, where patients classified in categories 2-4 were less likely to experience a relapse compared to those in the first category among patients with malignant indications (HRs < 1). Despite low event rates, the Kaplan-Meier survival curves for long-term outcomes of time to secondary graft failure requiring second HCT and time to initiating TA-TMA indicate that those in the worst category have the worst survival probabilities, although there is significant uncertainty in these estimates (see Supplementary Material 2).

**Figure 2:**
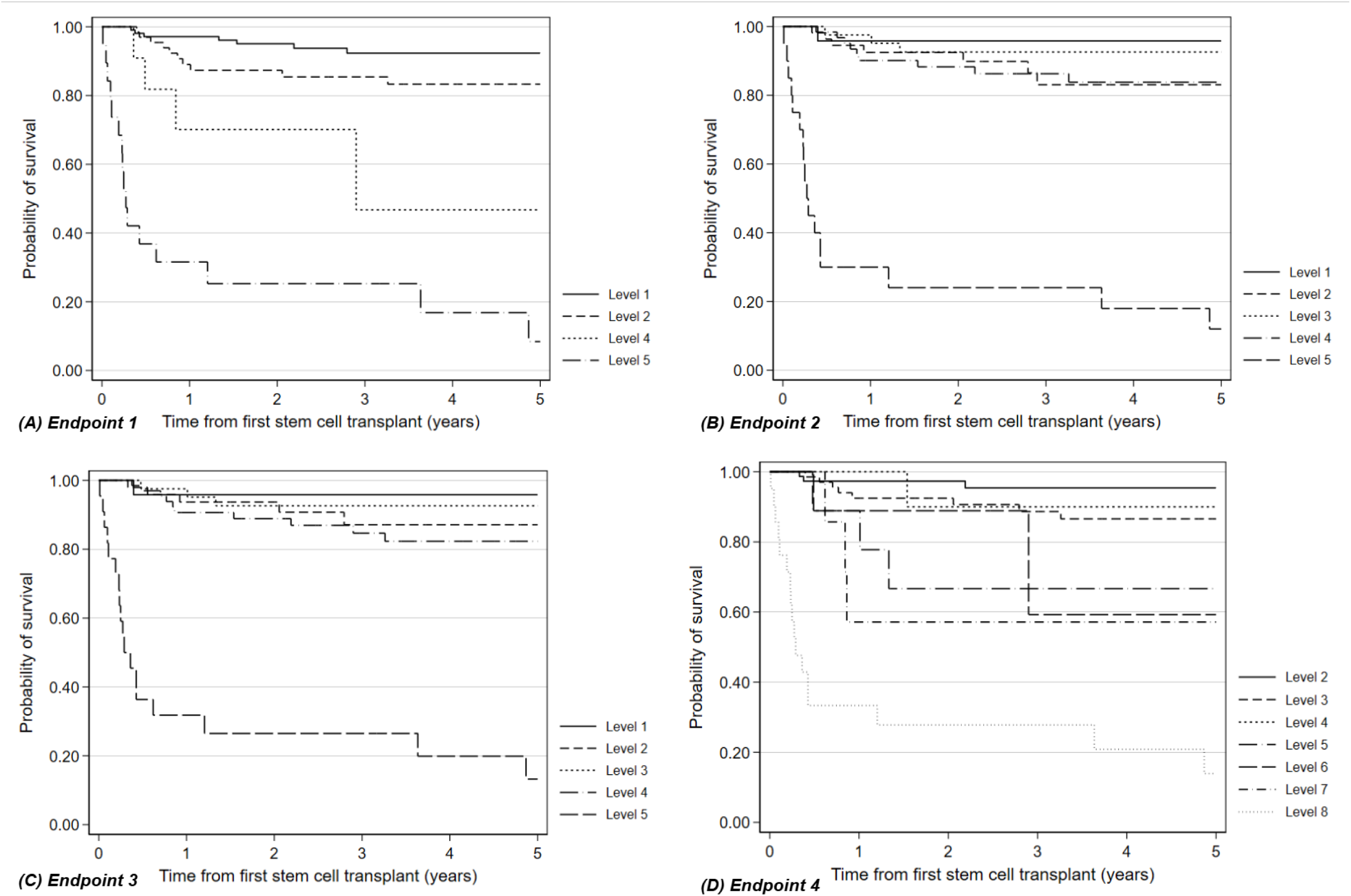
Kaplan-Meier survival curve for time to death by endpoint category for the 4 proposed ordinal endpoints.

**Table 4:**
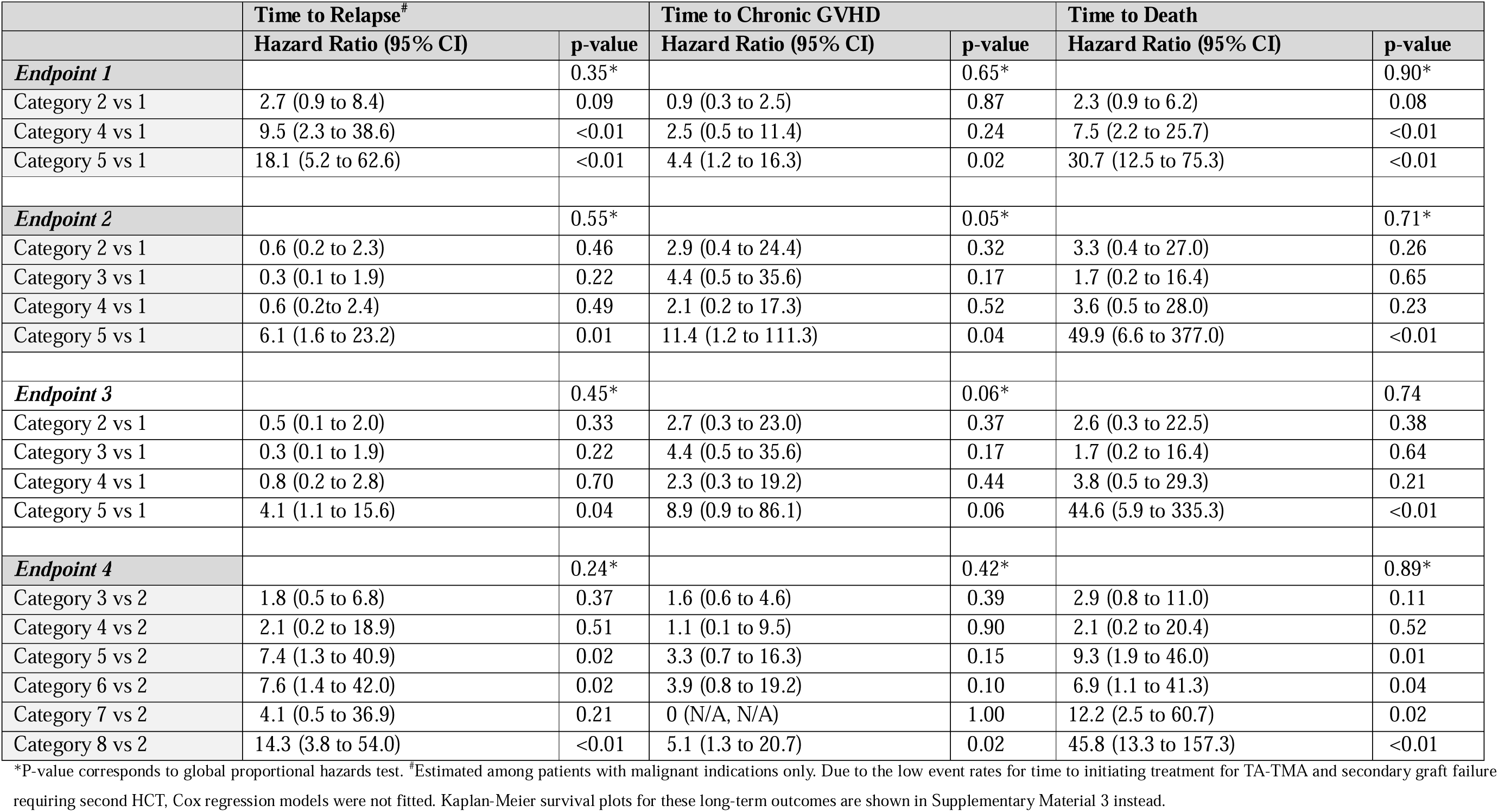
Summary of associations between the proposed ordinal endpoint categories and long-term outcomes.

### Effect of exposures with known efficacy using the proposed endpoints

Using the proposed endpoints to estimate the effect of donor type found that sibling matched donors had higher odds of a more favourable outcome compared to matched unrelated and haploidentical donors among those with malignant indications for all four endpoints which would be expected (Table 5; and Figure 3). Among those with non-malignant indications, all proportional ORs were < 1 assessing the effect of haploidentical to sibling matched donors for Endpoints 2, 3 and 4. However, among patients with non-malignant indications the proportional OR = 1.13 for Endpoint 1 suggests sibling matched donors had slightly worse outcomes than matched unrelated donors which would not be expected. For all endpoints, patients who received a cord blood transplant had higher odds of less favourable outcomes compared to those who did not receive a cord blood transplant. For all endpoints, among patients with B-ALL those with minimal residual disease had higher odds of less favourable outcomes compared to those who did not report any minimal residual disease (all proportional ORs close to 0.50).

**Figure 3:**
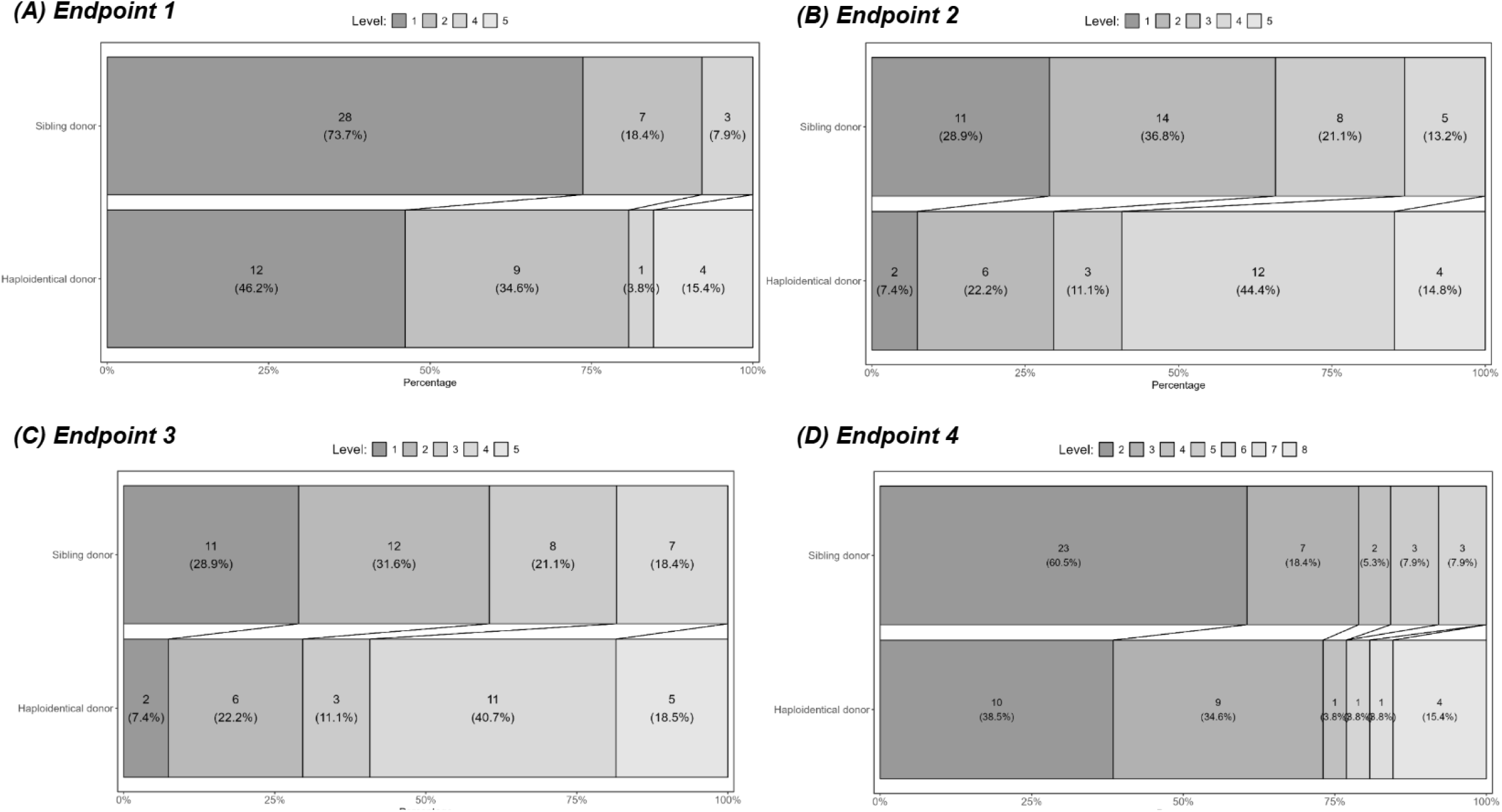
Grotta bar charts for all proposed ordinal endpoints by transplant type (haploidentical vs sibling donor) among those with malignant indications.

Among those with a non-cord blood transplant, patients who had high risk CMV mismatch were more likely to have less favourable outcomes compared to those who were not a high-risk mismatch for Endpoints 2 and 3. However, the proportional OR were in the opposite direction than expected for the Endpoints 1 and 4, such that patients who were a high-risk mismatch reported better outcomes compared to those who were not.

Visualising the ORs and corresponding 95% CI estimated from separate (adjusted) logistic regression models and the Grotta bar charts suggested that the proportional odds assumption was plausible in most scenarios (see Figure 3 and Supplementary Material 3 and 4).

**Table 5:**
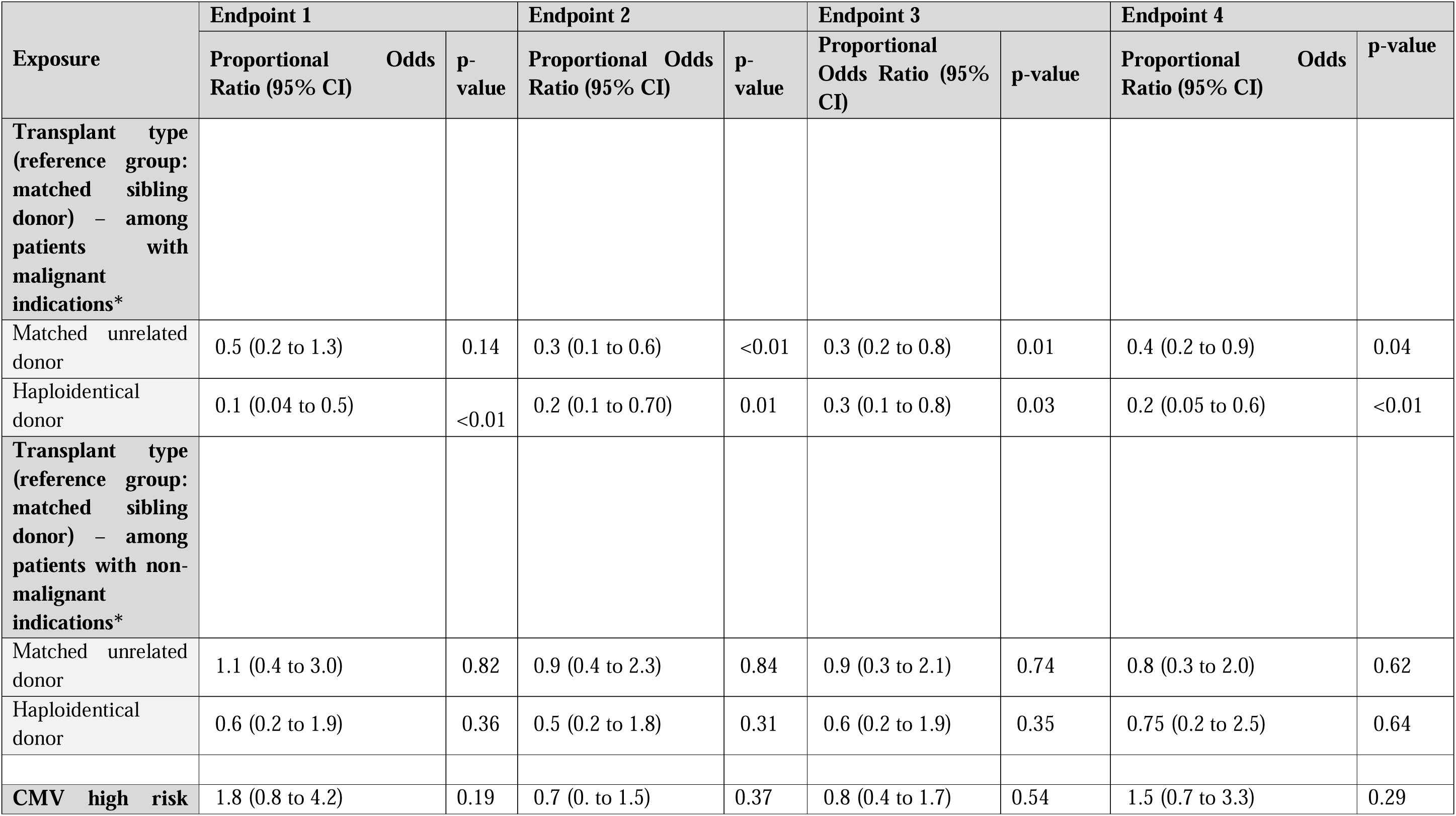
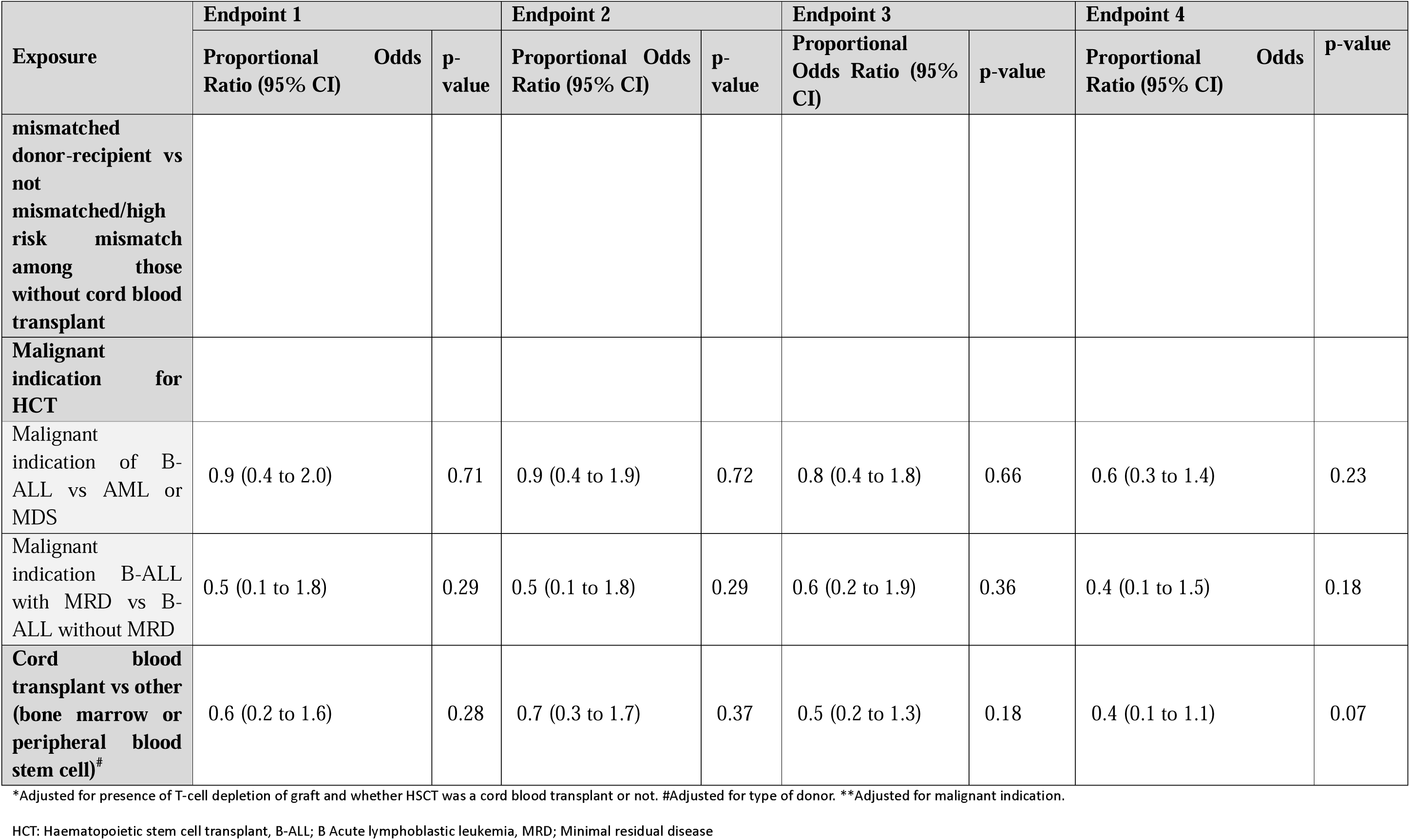
Summary of relationship between exposures with known efficacy and proposed ordinal endpoints estimated using proportional odds models.

### Expert and consumer feedback

Feedback on the 4 proposed endpoints was collected for 14 individuals spread across a range of clinicians in HCT and patient/family consumers. This cohort identified some concerns that data collection for the eight-point scale was more complex than for the five-point scales. They felt that organ support would be more feasible if defined as single or multi-organ support (Endpoint 2) compared to major and minor organ support (Endpoint 1). Consumers were most influenced by whether the endpoints correlated with overall survival and cGHVD. In total, 93% (13/14) of consumers considered Endpoint 2 to be both feasible and applicable to consumers in the HCT cohort.

Based on the results of both the statistical validation and consumer feedback the decision was therefore made to proceed with the Endpoint 2 for the planning of BANDICOOT

## DISCUSSION

We have statistically validated four possible short-term ordinal endpoints for use in a paediatric HCT APT. We have demonstrated that the chosen primary ordinal endpoint, Endpoint 2, is strongly associated with long-term outcomes, key exposures with known efficacy are shown as such using the selected outcome, and that data can be feasibly and routinely collected. This is the first paper to develop an ordinal endpoint for a paediatric HCT APT, and the first novel paediatric HCT endpoint that has been co-developed by consumers and a broad range of healthcare workers and researchers. The developed endpoint incorporates recent literature and key clinical states from the past decade that significantly impact transplant outcomes (e.g., CD4+ immune reconstitution, viral load, and ICU support) and therefore reflects modern HCT practices and advances ^2–7^.

When selecting an ordinal outcome, it is important that participants are spread across the different categories of the outcome as this provides the greatest power and means that there is the opportunity for interventions to shift participants up and down the scale. Compellingly, the patients were relatively evenly spread across the categories of Endpoint 2 and 3. This was not the case with Endpoints 1 and 4, where patients were concentrated in the lowest categories. The implications of using such an endpoint in practice would be that an intervention would not be able to show any improvement, since the patients are already concentrated in the ‘best’ or ‘healthiest’ patient states.

One of the key long-term outcomes that is predicted by the chosen ordinal endpoint is chronic GVHD, which occurs as a complication after HCT in 6-33% of paediatric patients^25^. Chronic GVHD is associated with a reduced risk of disease relapse but unfortunately contributes significantly to post HCT morbidity accounting for up to 37.5% of non-relapse mortality.^25,26^ Chronic GVHD develops late post HCT (beyond the 100-day censor point) with the most common risk factor being the development of acute GVHD early post HCT. Our validation results demonstrate that Endpoint 2 is strongly associated with the development of chronic GVHD with patients classified in a higher category (or ‘worse’ health state) associated with an increased likelihood of developing chronic GVHD, compared to a lower category. This is likely driven by the fact that low CD4 count and prolonged viral reactivation are part of the endpoint criteria^2–4^, both of which are known to be strongly associated with the development of moderate to severe acute GVHD and, in turn, chronic GVHD.

Similarly to chronic GVHD, disease relapse post HCT often occurs beyond 100 days post HCT and is associated with post HCT mortality.^27^ In patients who undergo HCT to treat an underlying malignancy, the main purpose of the HCT is to maintain lifelong graft versus leukaemia effect and avoid relapse. Our validation results show that in patients with a malignant indication for HCT, the highest level in Endpoint 2 was associated with a higher risk of relapse. Interestingly, however, patients classified in the first category had a lower risk of relapse compared to categories two through to four. This is in contrast with the risk of death for Endpoint 2 where time to death is lowest in patients in the first category compared to all other categories, with the greatest difference seen for patients in the highest category. We hypothesise that this is due to more robust immune reconstitution (reflected by higher CD4+ counts) and graft versus leukaemic effect ^28^. Most immune reconstitution and other significant events (e.g. survival and reduced relapse) have been associated with CD4+. Of importance, whilst after HCT, early CD4+ recovery is mostly driven by peripheral expansion of memory T cells, as time progresses patients begin thymic regeneration of naïve T cells. These naïve T cells are responsible for developing new adaptive immune responses against leukaemic antigens and support the development of tumour specific CD8+T cells responsible for graft versus leukaemic effects ^29^.

Importantly, the emergence of prolonged viral infections in the immediate post HCT period is a risk factor for morbidity and mortality, the clearance of which is intrinsically linked to immune reconstitution ^4^. The lower risk of death in the first category of Endpoint 2 (for those patients without relapsed disease) is likely also related to robust immune reconstitution and lack of significant viral infection within this category. The converse is true for category five, which was associated with a higher risk of death, poorer immune reconstitution and prolonged significant viral exposures compared with the earlier categories. In Australia, disease relapse is the most common cause of death following HCT. Disease relapse accounts for 62% of deaths from a matched sibling donor HCT, 68% from a matched unrelated donor and 47% from a haploidentical donor transplants (ANZTCT 2022). Viral reactivations are the leading cause of non-relapse mortality^1^, accounting for 56% and 67% of deaths in children receiving a HCT from a matched unrelated donor or haploidentical donor respectively. Therefore, by including CD4+ reconstitution together with viral reactivations (incorporating significant levels and time of exposure), Endpoint 2 accounts for the drivers of disease morbidity and mortality in the paediatric HCT cohort.

The finding that the proposed endpoints demonstrated the expected effect for most transplant-related exposures was also a key factor for determining which endpoint score would be used in BANDICOOT. One of these factors was transplant donor, specifically haploidentical versus matched sibling donor. In the majority of HCT indications a matched sibling donor is preferable due to the lowest rates of non-relapse morbidity ^30^. Unfortunately, less than half of patients have an available matched sibling donor compared to the more readily available haploidentical donor. The finding that Endpoint 2 showed a strong positive effect in those who received a matched sibling donor HCT compared to haploidentical HCT is consistent with this known effect. Endpoint 2 also showed the poor prognostic effect consistant with what is expected for patients with diagnoses of B-ALL, with positive MRD in the month prior to HCT ^31,32^.

Three critical enablers have enhanced this detailed validation. Firstly, access to a granular and detailed modern day paediatric HCT database (BILBY) from the start of the design process. Secondly, input from the CHA to obtain large-scale missing data, to avoid further manual data collection, using efficient data coding. The investment of time and funding from the CHA to support this work is an important consideration for other groups wanting to perform a similar validation process for an APT. Thirdly, the invaluable regular input from statisticians has allowed for informed collaboration from the initial design of the four potential ordinal endpoints, whilst ensuring their relevance for use in the APT. The above facilitated both efficient and comprehensive results to present to consumers. In future it would also be important to consider validating this primary endpoint in other populations, such as in other paediatric HCT centres outside of Australia. There are also potential future applications of this primary ordinal endpoint in patients receiving an autologous transplant, who are often managed in the same HCT units as allogeneic HCT recipients.

As well as needing to have a large granular database of patient information to validate the endpoints, one limitation of using a retrospectively collected database is that some of the data required for specific endpoints were documented as free text in the medical record, such as the need for gastrointestinal organ support (e.g. ascitic drain insertion) making them difficult to obtain from the EMR. This may be overcome by prospectively collecting this information in a dedicated database, or through the use of “smart phrases” where patient information is documented in a reproducible way in the EMR and can be identified more reliably by data analysts, however this requires a significantly longer investment of time and resources to collect an equivalent number of patients. Another limitation of this work is that for transplant outcomes; TA-TMA requiring treatment and secondary graft failure requiring subsequent transplants there was considerable uncertainty in the estimates of associations. This reflects the low numbers of patients in both categories. It is important to note that the historical cohort was not undergoing routine screening for TA-TMA with sC5b-9 at the time of data collection. Therefore, we have concluded that this result was biased by both limited sample size and likely does not reflect the true denominator of disease; with an increase in awareness of TA-TMA and the ability to screen with sC5b-9 our incidence of TA-TMA has increased.

## Data Availability

All data produced in the present work are contained in the manuscript and supplementary file

## SUMMARY

In summary, we describe the validation process for determining the primary ordinal endpoint chosen to be included for the paediatric HCT APT, BANDICOOT. This involved the application of data from ‘real-world’ patients who have undergone allogeneic HCT, obtained through both manual data extraction and large dataset exports. This dataset was used to statistically validate 4 potential ordinal endpoints measured at 100 days post HCT, and determine which was best suited to the APT based on the distribution of patients and correlation with ‘traditional’ long term transplant endpoints e.g. cGVHD and OS. This process identified Endpoint 2 as both statistically valid and the most appropriate endpoint by consumers and clinicians. We believe this endpoint has the potential to create a paradigm shift in paediatric HCT RCTs due to its broad applicability across a range of HCT interventions and outcomes, trumping the time-to-event approach of modern HCT endpoints.

