## Supplemental file for "*“*Validation of a novel ordinal endpoint for an adaptive platform trial in paediatric haematopoietic stem cell transplant*”*"

**SUPPLEMENTARY MATERIAL 1: Definition of endpoints**

**Supplementary Table 1:** Major and minor organ support for Ordinal Endpoint 1, in Table 2

| Organ | Support | Classification |
| --- | --- | --- |
| Respiratory | Mechanical Ventilation or ECMO | Major |
|  | Non Invasive Ventilation (CPAP/BiPAP) | Major |
| Hepatic | Haemofiltration | Major |
|  | Ascitic drain | Minor |
| Renal | Haemofiltration, dialysis, ECP | Major |
|  | Intravenous antihypertensives | Minor |
| Cardiovascular | Inotropes and/or vasopressors and/or pericardial drain without other major end organ support | Minor |
|  | Inotropes and/or vasopressors with other major organ support | Major |
| Neurologic | Neurological protection in ICU, mechanical ventilation +/- neuroprotective measures e.g. cooling, tilting | Major |
| Gastrointestinal tract | Bowel perforation or pneumotosis intestinalis or ischaemia present on gross inspection (surgical) or by plain abdominal x-ray, CT or MRI abdomen. | ? Major |

**Supplementary Table 2**: Multi-organ support; defined as two or more organ involvement above

| Organ | Support |
| --- | --- |
| Respiratory | Mechanical Ventilation and/or ECMO |
|  | Non Invasive Ventilation (CPAP/BiPAP) |
|  | High flow O2 |
| Hepatic | Haemofiltration |
| Renal | Haemofiltration, dialysis, extracorporeal photopheresis. |
| Cardiovascular | Inotropes and/or vasopressors and/or ECMO* |
| Neurologic | Neurological protection in ICU, mechanical ventilation +/- neuroprotective measures |
| Gastrointestinal | Bowel perforation or pneumotosis intestinalis or ischaemia present on gross inspection (surgical) or by plain abdominal x-ray, CT or MRI abdomen. |

**SUPPLEMENTARY MATERIAL 2: Distribution of patients across categories of the proposed endpoints**

**Table 3a:** Distribution of patients across categories of the proposed 5-point ordinal endpoints

|  | **Endpoint 1** | **Endpoint 2** | **Endpoint 3** |
| --- | --- | --- | --- |
| **Level 1** | 105 (52.2%) | 24 (11.9%) | 24 (11.9%) |
| **Level 2** | 66 (32.8%) | 55 (27.2%) | 49 (24.3%) |
| **Level 3** | 0 (0.0%) | 41 (20.3%) | 41 (20.3%) |
| **Level 4** | 11 (5.5%) | 62 (30.7%) | 66 (32.7%) |
| **Level 5 (death)** | 19 (9.5%) | 20 (9.9%) | 22 (10.9%) |
| **Missing** | 1 observation | 0 observations | 0 observations |

**Table 3b:** Distribution of patients across categories of Endpoint 4

|  | **Endpoint 4** |
| --- | --- |
| **Level 1** | 0 (0.0%) |
| **Level 2** | 75 (37.3%) |
| **Level 3** | 68 (33.8%) |
| **Level 4** | 12 (6.0%) |
| **Level 5** | 9 (4.5%) |
| **Level 6** | 9 (4.5%) |
| **Level 7** | 7 (3.5%) |
| **Level 8 (death)** | 21 (10.4%) |
| **Missing** | 1 observation |

**SUPPLEMENTARY MATERIAL 3: Kaplan-Meier Survival Curves for Longer-Term Outcomes**

1. ***Time to relapse***

We present Kaplan-Meier survival curves for time to relapse in Figures 1a-d.

**Figure 1a:** Kaplan-Meier survival curve for time to relapse for categories of the first proposed ordinal endpoint

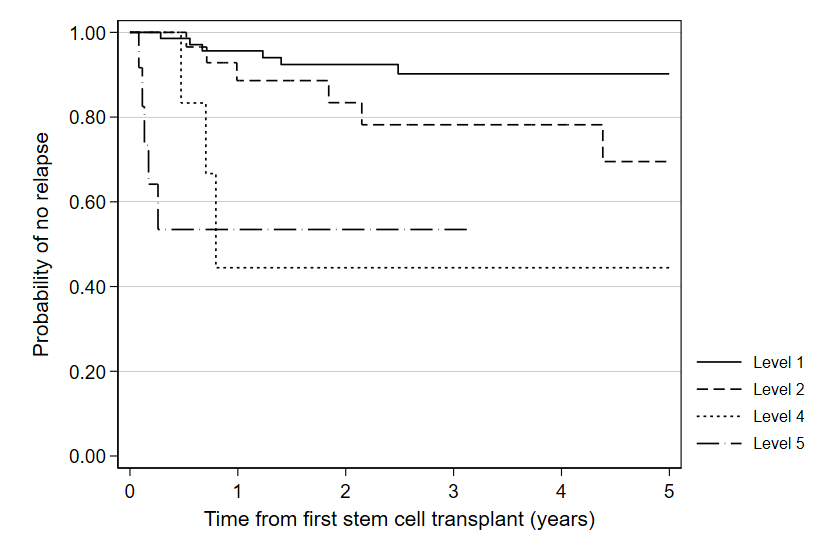

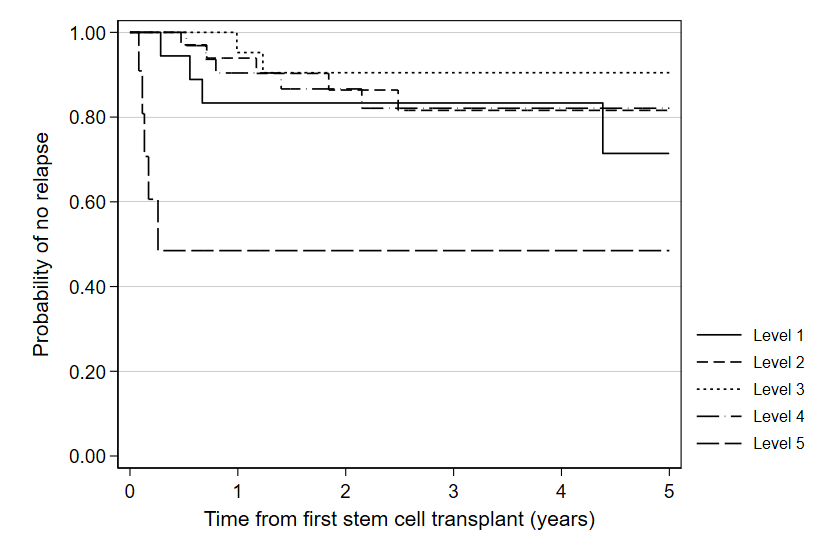
**Figure 1b:** Kaplan-Meier survival curve for time to relapse for categories of the second proposed ordinal endpoint

**Figure 1c:** Kaplan-Meier survival curve for time to relapse for categories of the third proposed ordinal endpoint

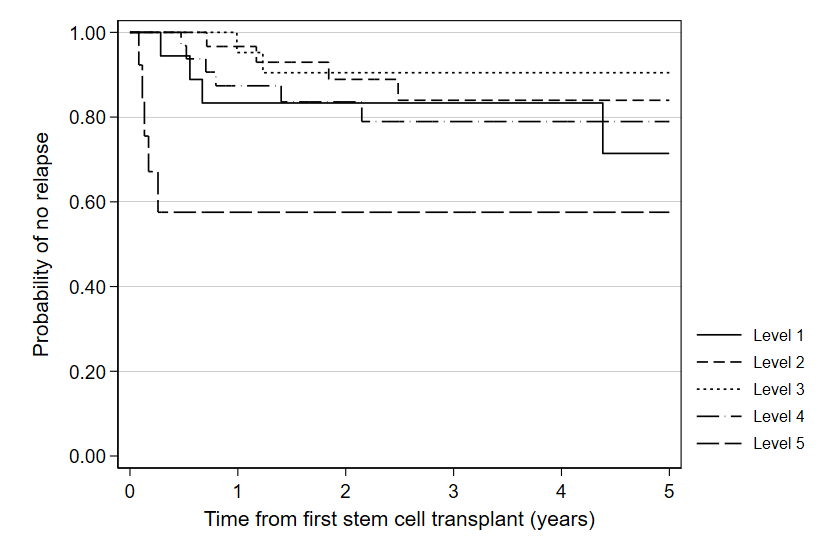

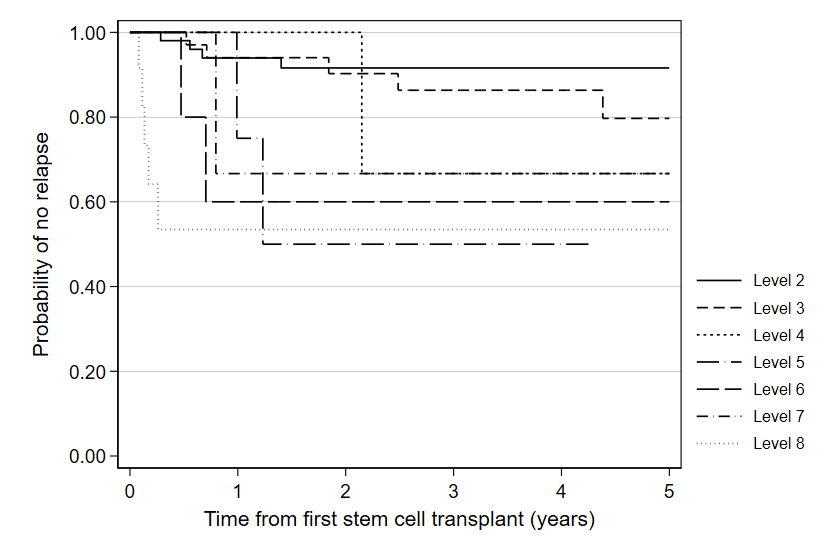
**Figure 1d:** Kaplan-Meier survival curve for time to relapse for categories of the fourth proposed ordinal endpoint

2. ***Time to secondary graft failure requiring second HSCT***

We first present Kaplan-Meier survival curves for time to secondary graft failure requiring second HSCT in Figures 2a-d.

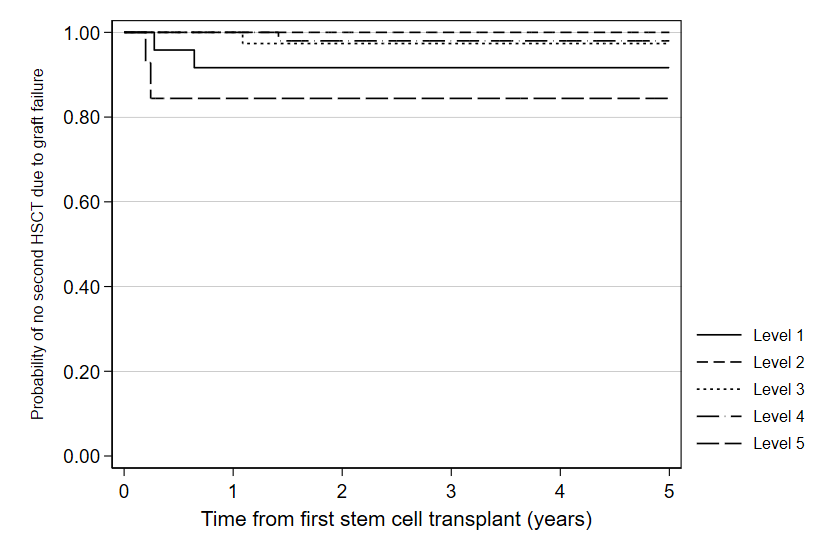
**Figure 2a:** Kaplan-Meier survival curve for time to secondary graft failure requiring second HSCT for categories of the first proposed ordinal endpoint

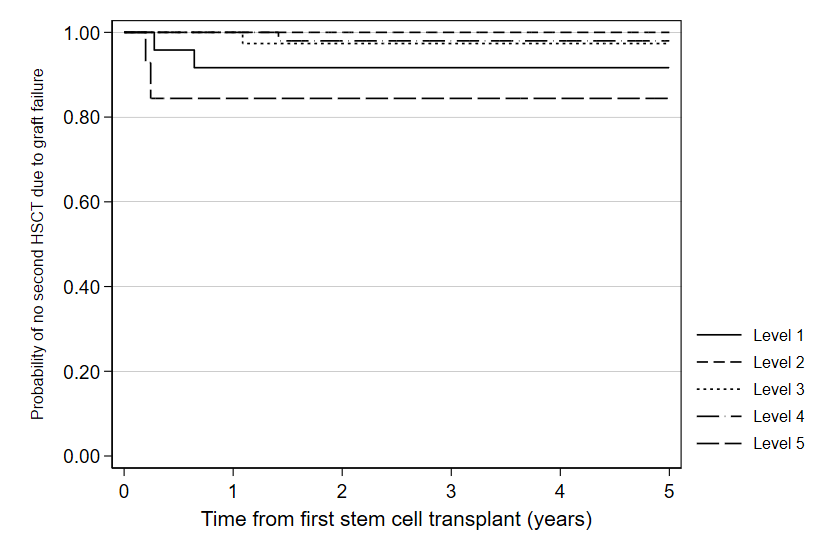
**Figure 2b:** Kaplan-Meier survival curve for time to secondary graft failure requiring second HSCT for categories of the second proposed ordinal endpoint

**Figure 2c:** Kaplan-Meier survival curve for time to secondary graft failure requiring second HSCT for categories of the third proposed ordinal endpoint

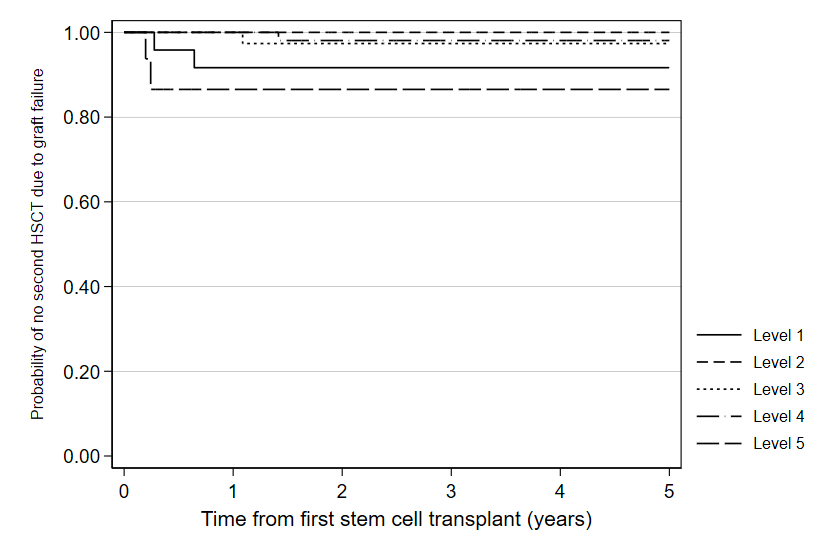

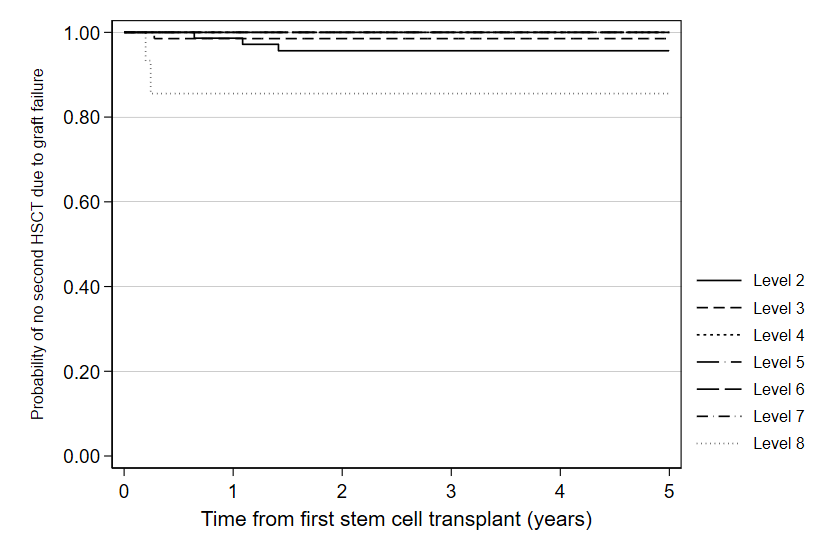
**Figure 2d:** Kaplan-Meier survival curve for time to secondary graft failure requiring second HSCT for categories of the fourth proposed ordinal endpoint

3. ***Time to initiating treatment for TA-TMA***

We first present Kaplan-Meier survival curves for time to initiating treatment for TA-TMA in Figures 3a-d.

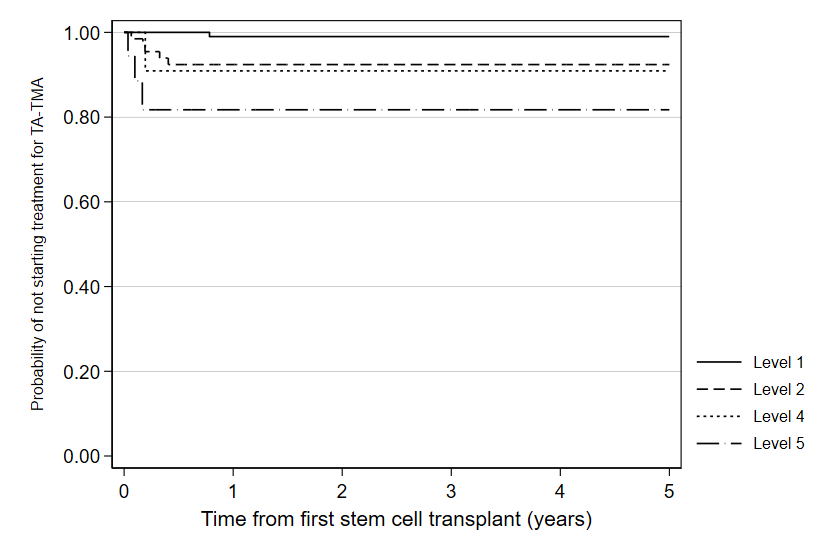
**Figure 3a:** Kaplan-Meier survival curve for time to initiating treatment for TA-TMA for categories of the first proposed ordinal endpoint

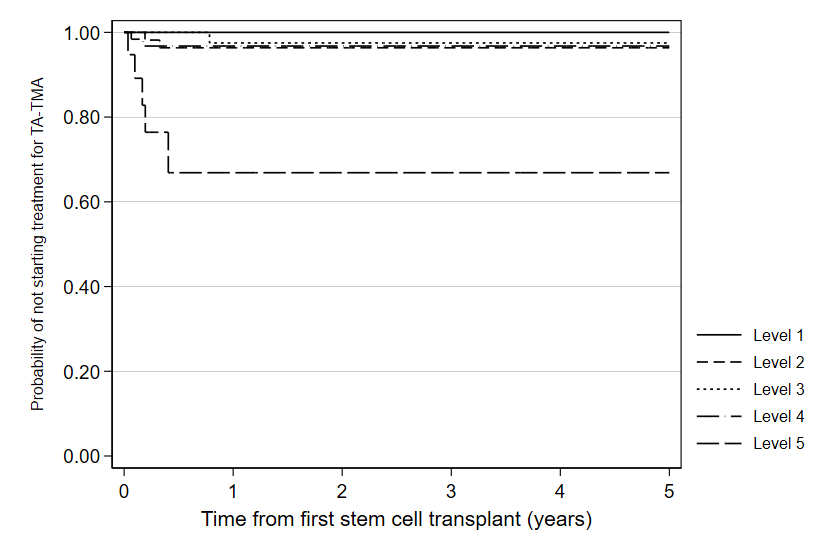
**Figure 3b:** Kaplan-Meier survival curve for time to initiating treatment for TA-TMA for categories of the second proposed ordinal endpoint

**Figure 3c:** Kaplan-Meier survival curve for time to initiating treatment for TA-TMA for categories of the third proposed ordinal endpoint

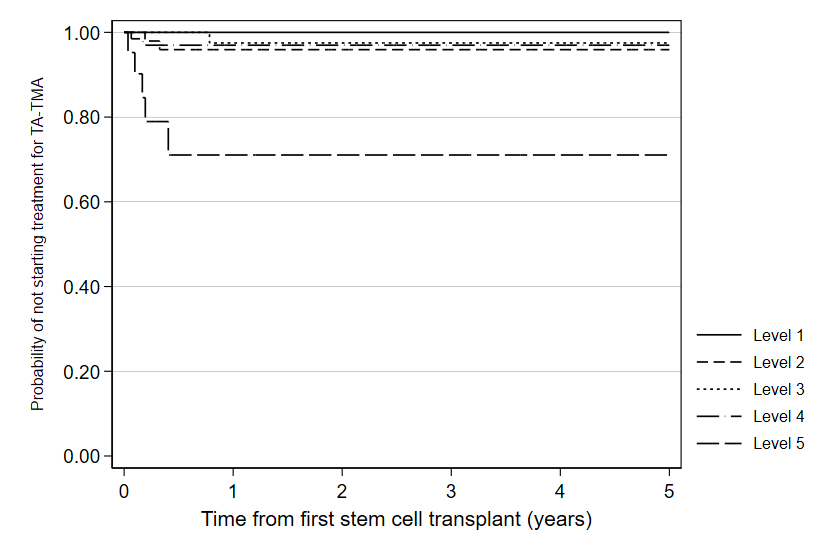

**Figure 3d:** Kaplan-Meier survival curve for time to initiating treatment for TA-TMA for categories of the fourth proposed ordinal endpoint

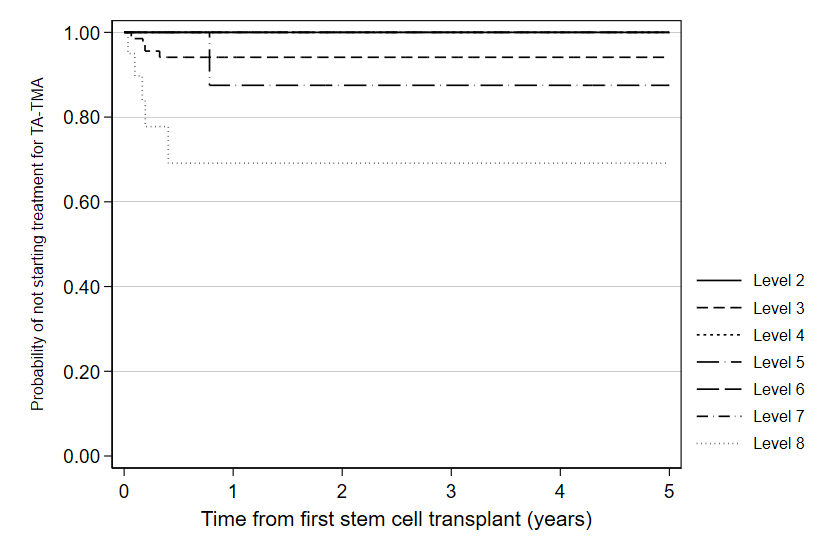

4. ***Time to chronic GVHD***

We first present Kaplan-Meier survival curves for time to chronic GVHD in Figures 4a-d.

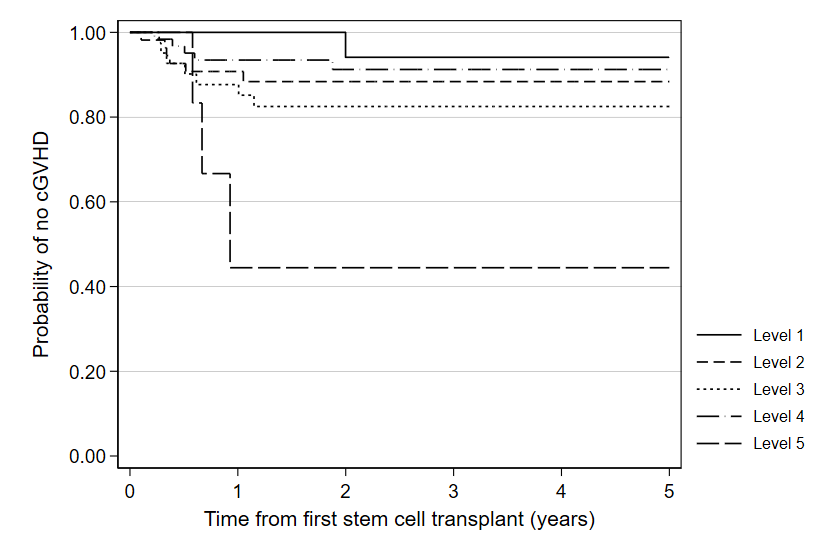
**Figure 4a:** Kaplan-Meier survival curve for time to chronic GVHD for categories of the first proposed ordinal endpoint

**Figure 4b:** Kaplan-Meier survival curve for time to chronic GHVD for categories of the second proposed ordinal endpoint

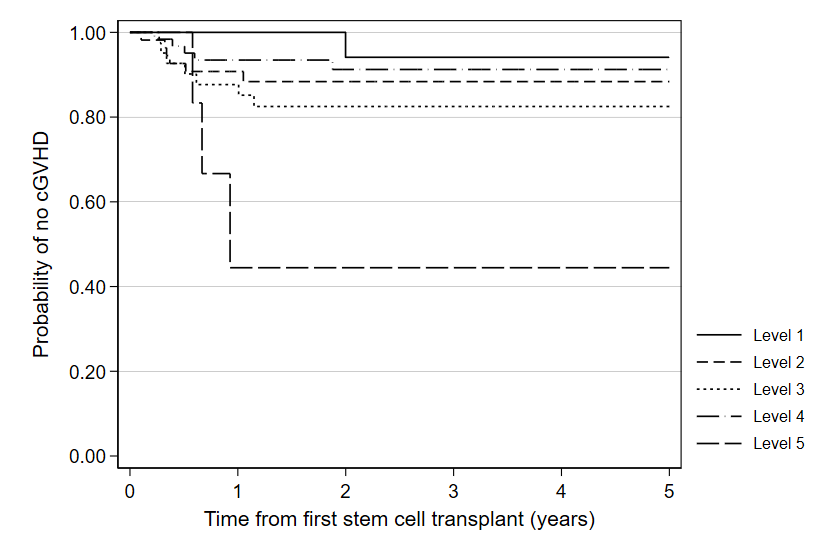

**Figure 4c:** Kaplan-Meier survival curve for time to chronic GHVD for categories of the third proposed ordinal endpoint

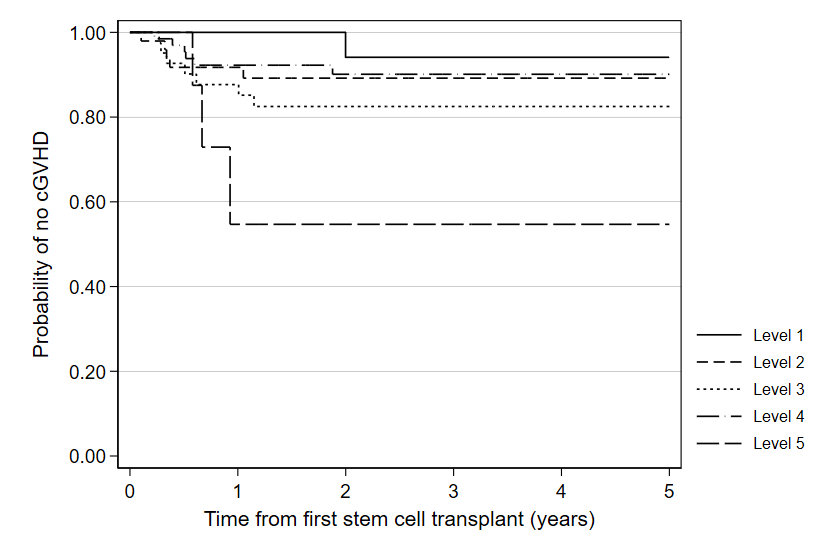

**Figure 4d:** Kaplan-Meier survival curve for time to chronic GHVD for categories of the fourth proposed ordinal endpoint

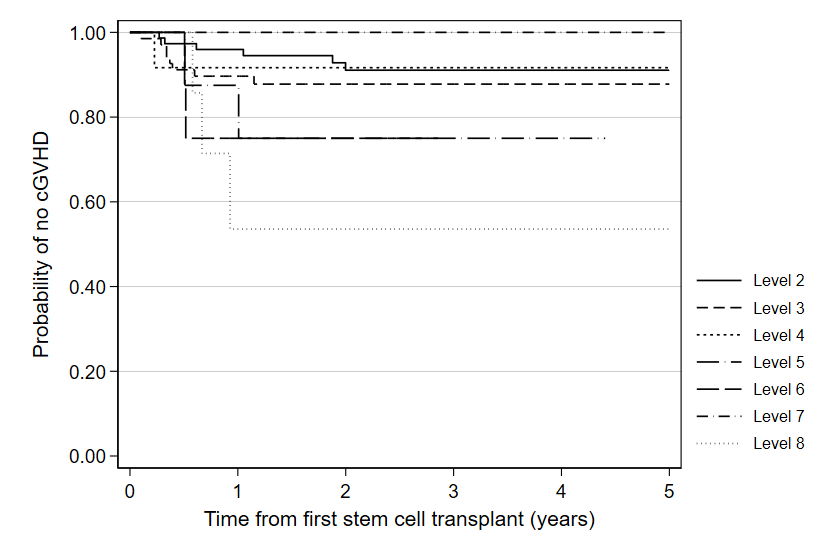

5. ***Time to death***

We first present Kaplan-Meier survival curves for time to death in Figures 5a-d. All endpoints appear strongly correlated with mortality by five years.

**Figure 5a:** Kaplan-Meier survival curve for time to death for categories of the first proposed ordinal endpoint

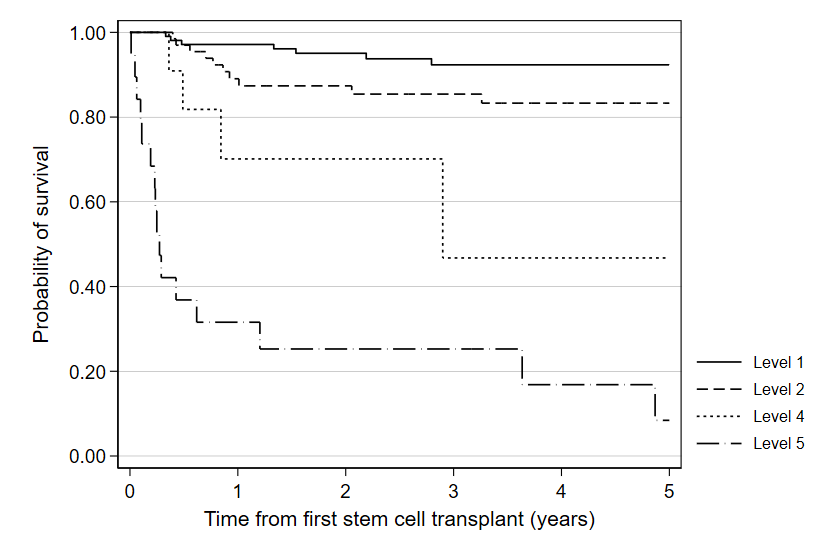

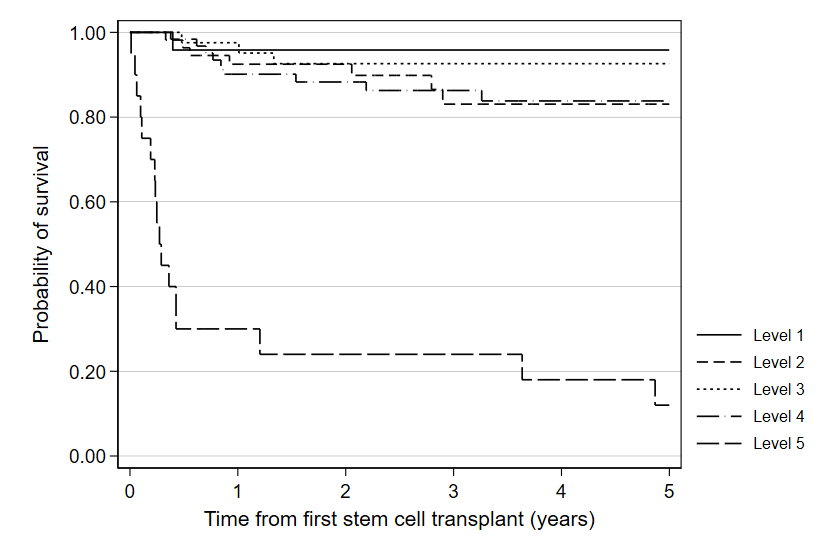
**Figure 5b:** Kaplan-Meier survival curve for time to death for categories of the second proposed ordinal endpoint

**Figure 5c:** Kaplan-Meier survival curve for time to death for categories of the third proposed ordinal endpoint

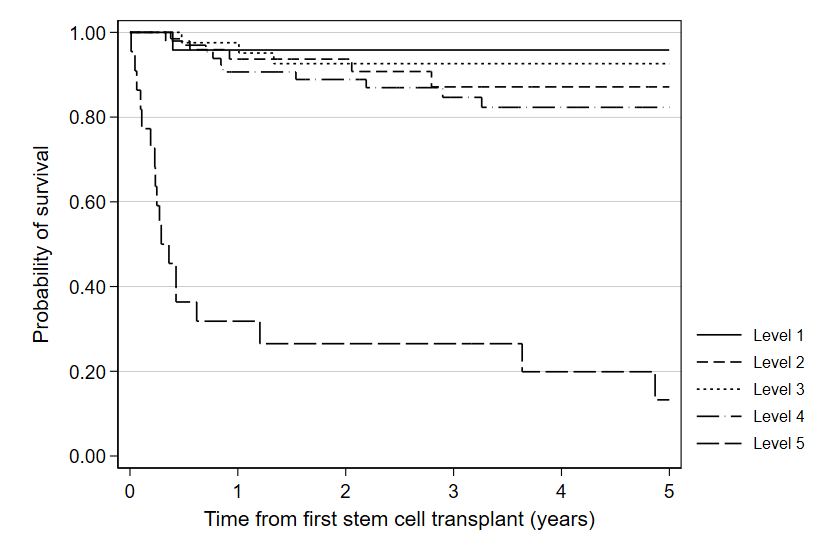

**Figure 5d:** Kaplan-Meier survival curve for time to death for categories of the fourth proposed ordinal endpoint

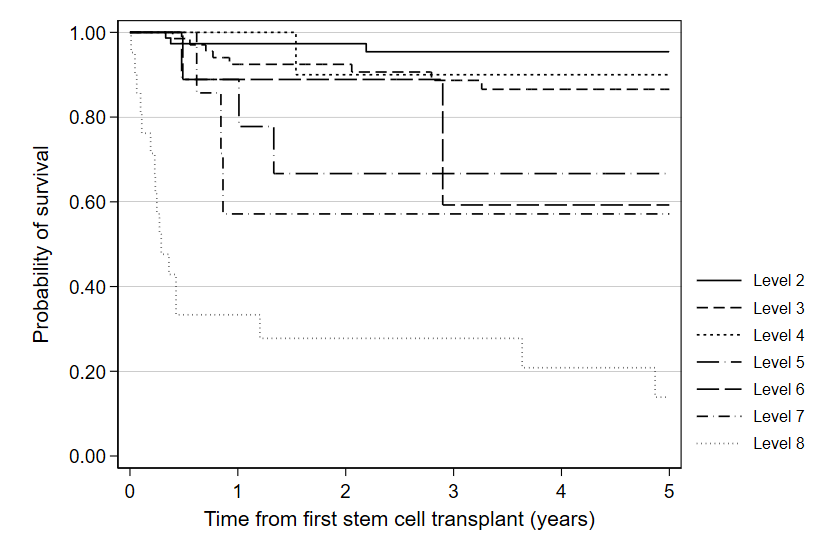

**SUPPLEMENTARY MATERIAL 4: Assessment of Proportional Odds Assumption for Exposure-Outcome Associations**

When interpreting the figures of the cut-point ORs for each exposure, all estimated cut-point ORs should be greater or less than 1 for proportional odds and stochastic ordering to hold (if the OR jumps above and below 1, this indicates the exposure exerts a different effect across the distribution of the ordinal categories). The proportional OR was added to each graph that is represented by the same colour as the exposure shown in the legend of the figure.

**Figure 6a:** Cut-point odds ratios for CMV mismatched donor status to assess proportional odds assumption for first ordinal endpoint

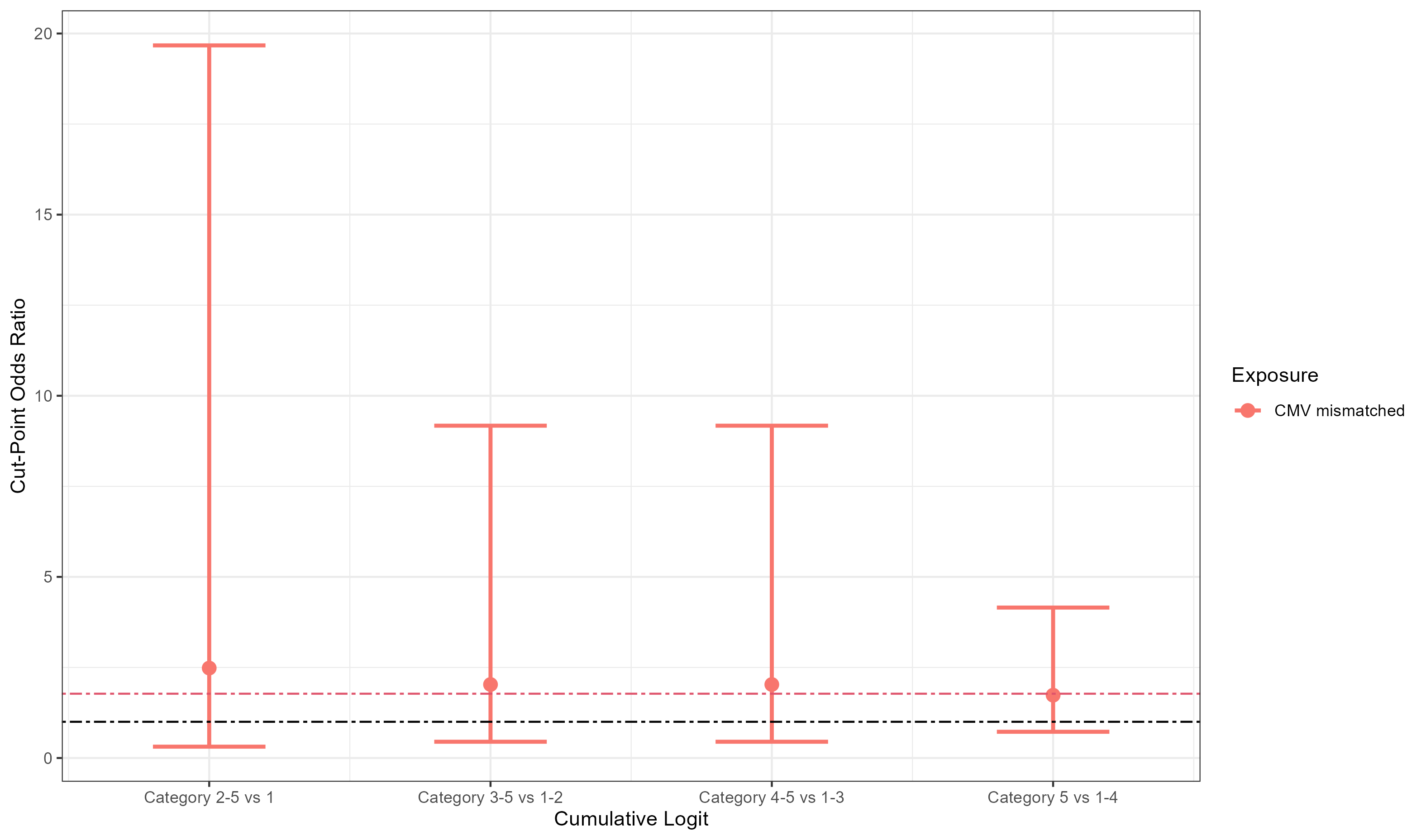

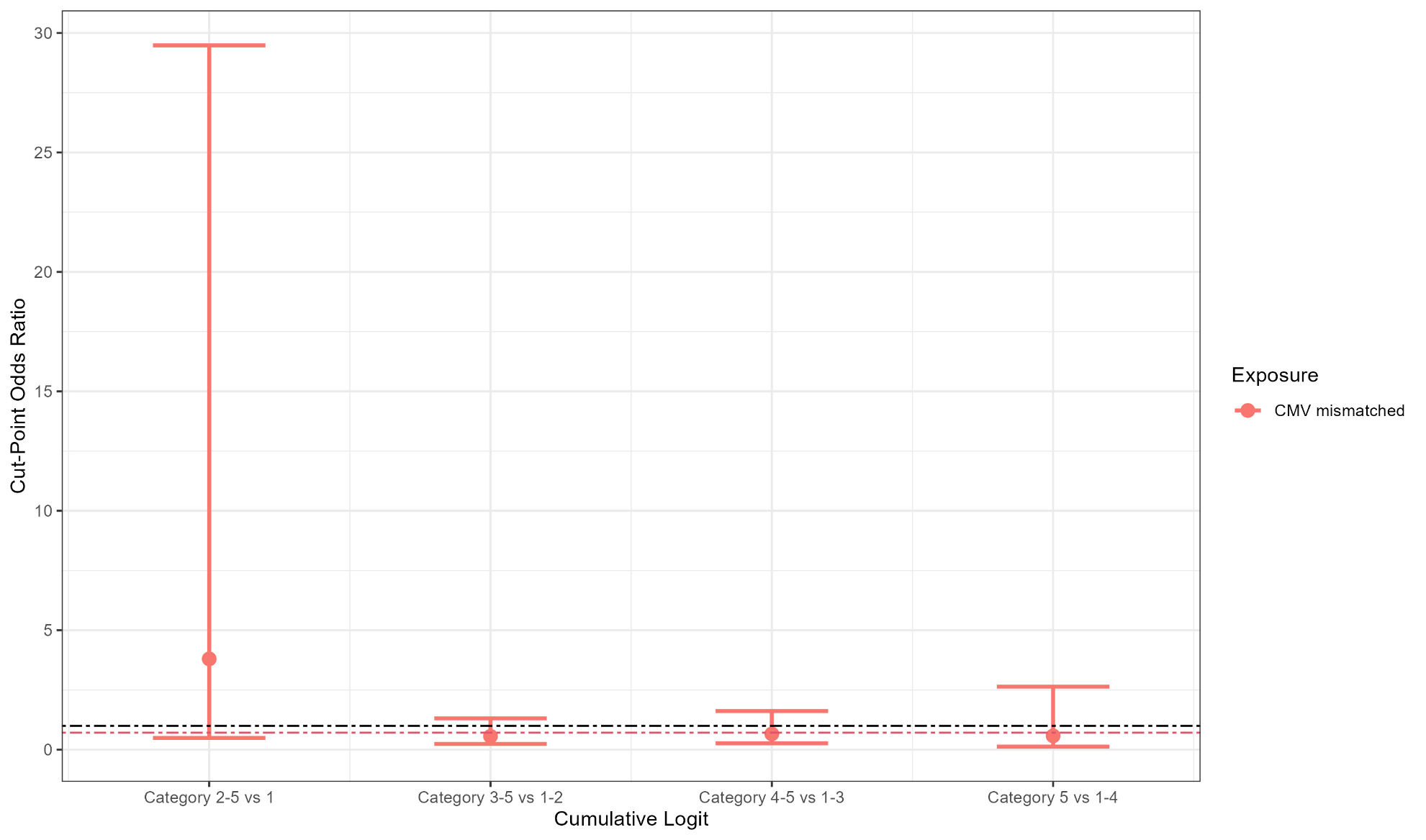
**Figure 6b:** Cut-point odds ratios for CMV mismatched donor status to assess proportional odds assumption for second ordinal endpoint

**Figure 6c:** Cut-point odds ratios for CMV mismatched donor status to assess proportional odds assumption for third ordinal endpoint

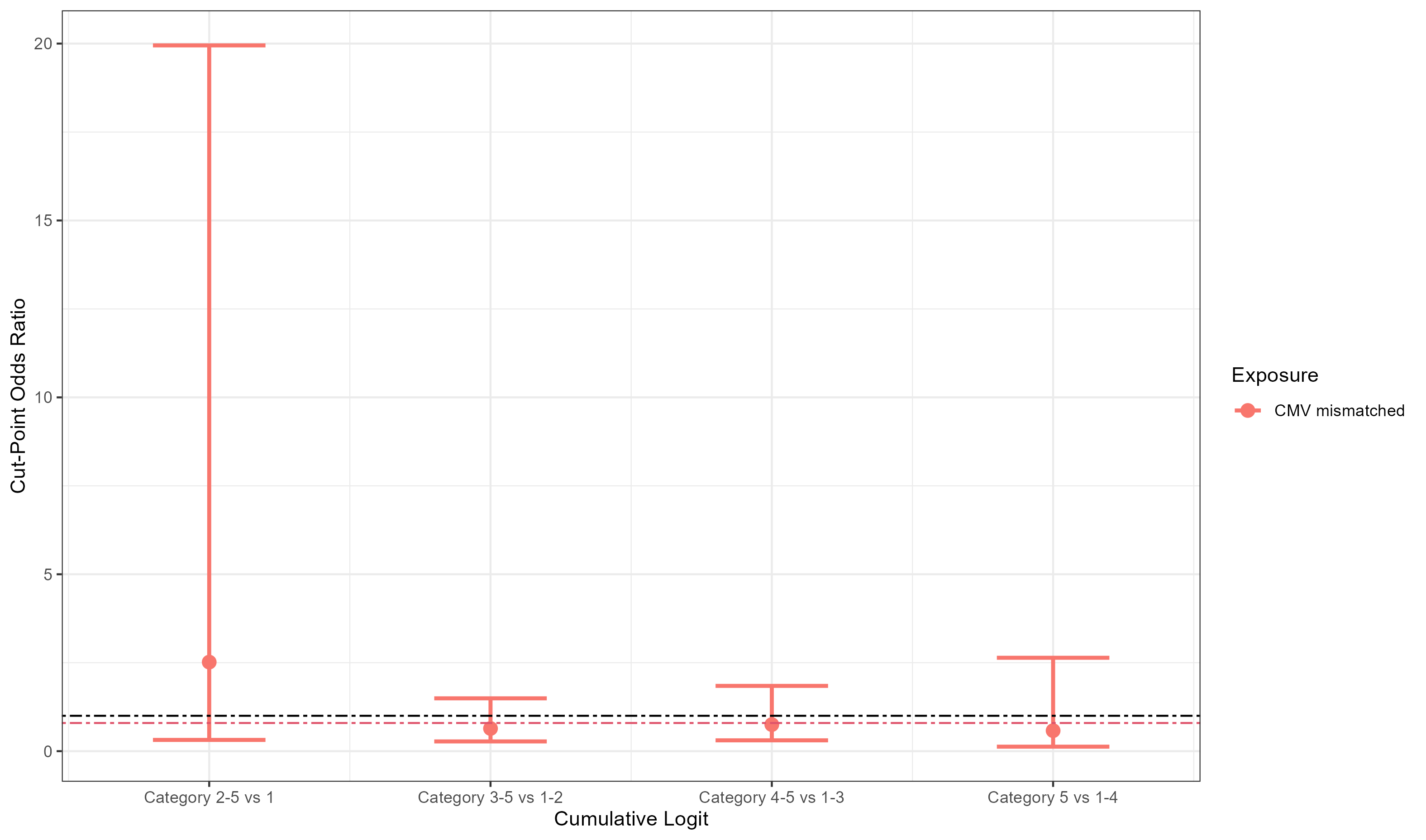

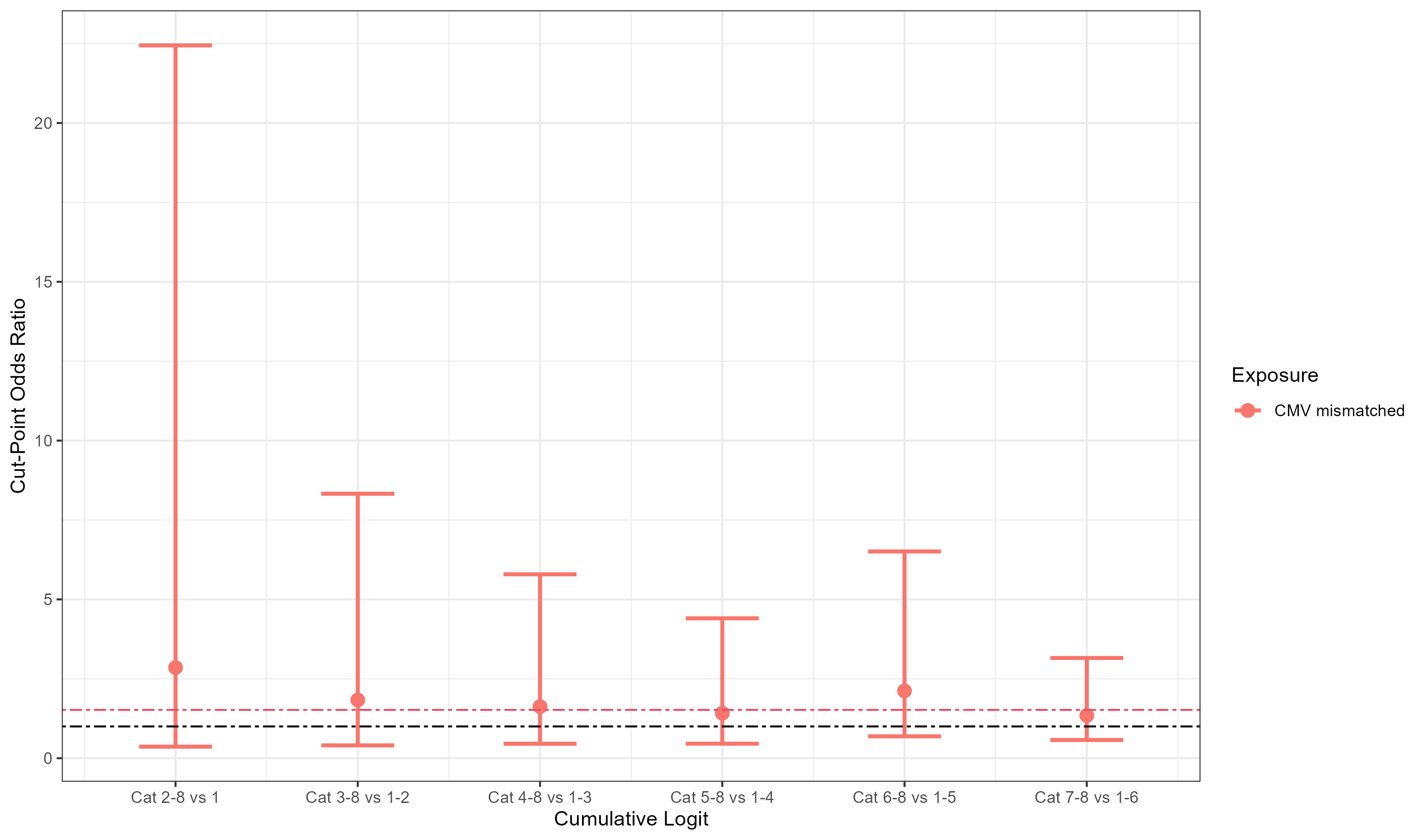
**Figure 6d:** Cut-point odds ratios for CMV mismatched donor status to assess proportional odds assumption for fourth ordinal endpoint

**Figure 7a:** Cut-point odds ratios for transplant donor type to assess proportional odds assumption for first ordinal endpoint (among those with malignant indication)

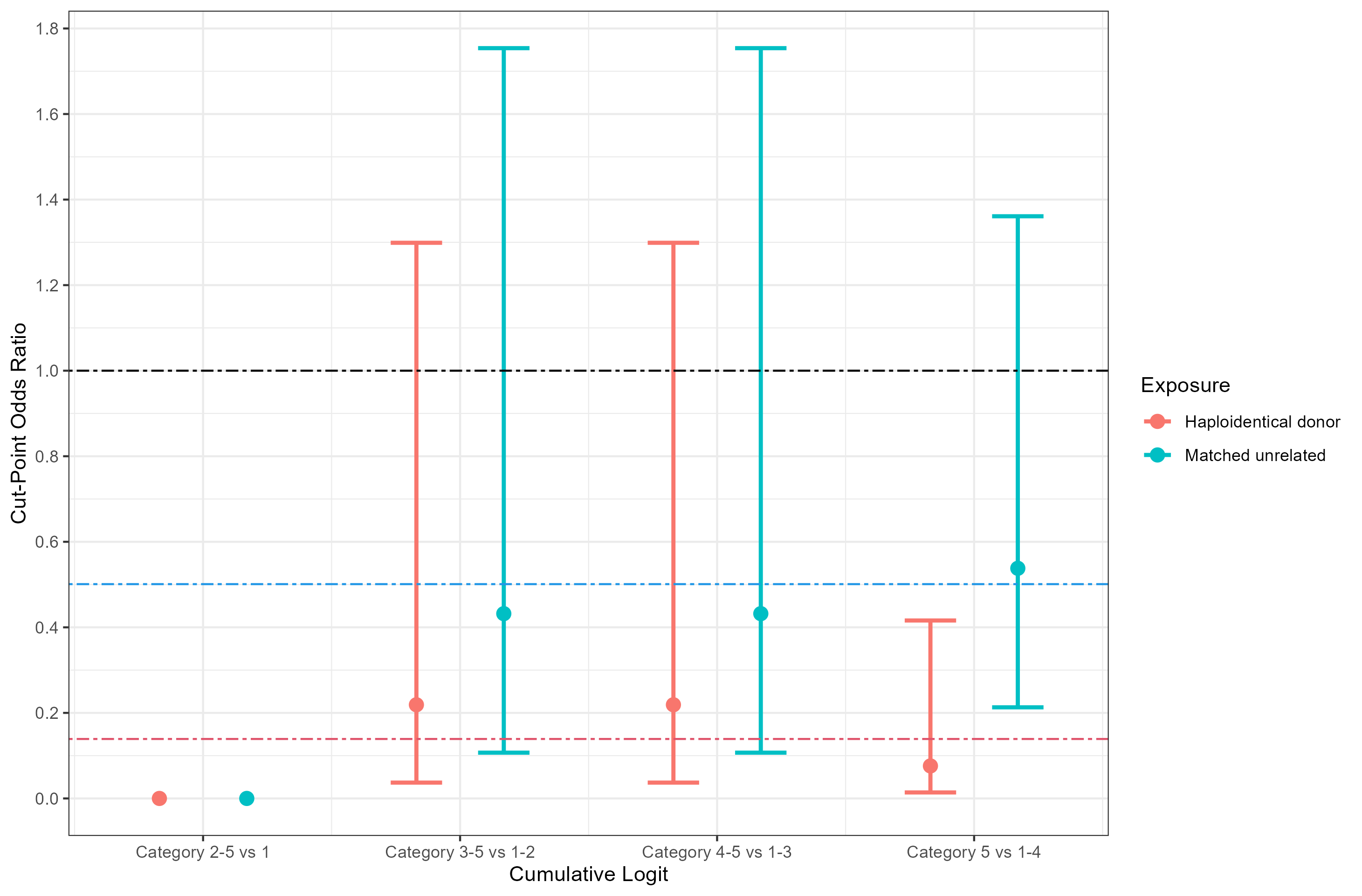

**Figure 7b:** Cut-point odds ratios for transplant donor type to assess proportional odds assumption for second ordinal endpoint (among those with malignant indication)

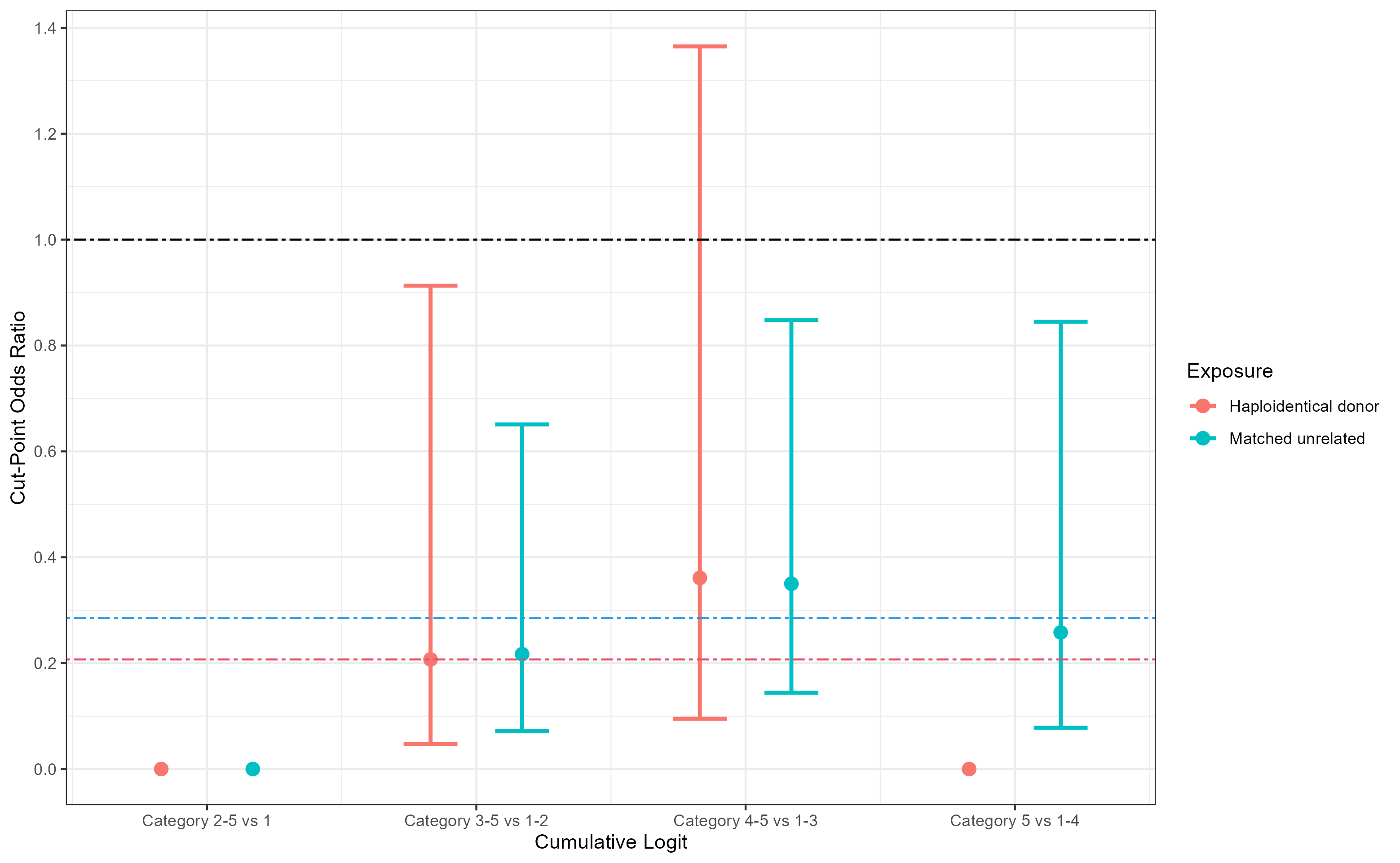

**Figure 7c:** Cut-point odds ratios for transplant donor type to assess proportional odds assumption for third ordinal endpoint (among those with malignant indication)

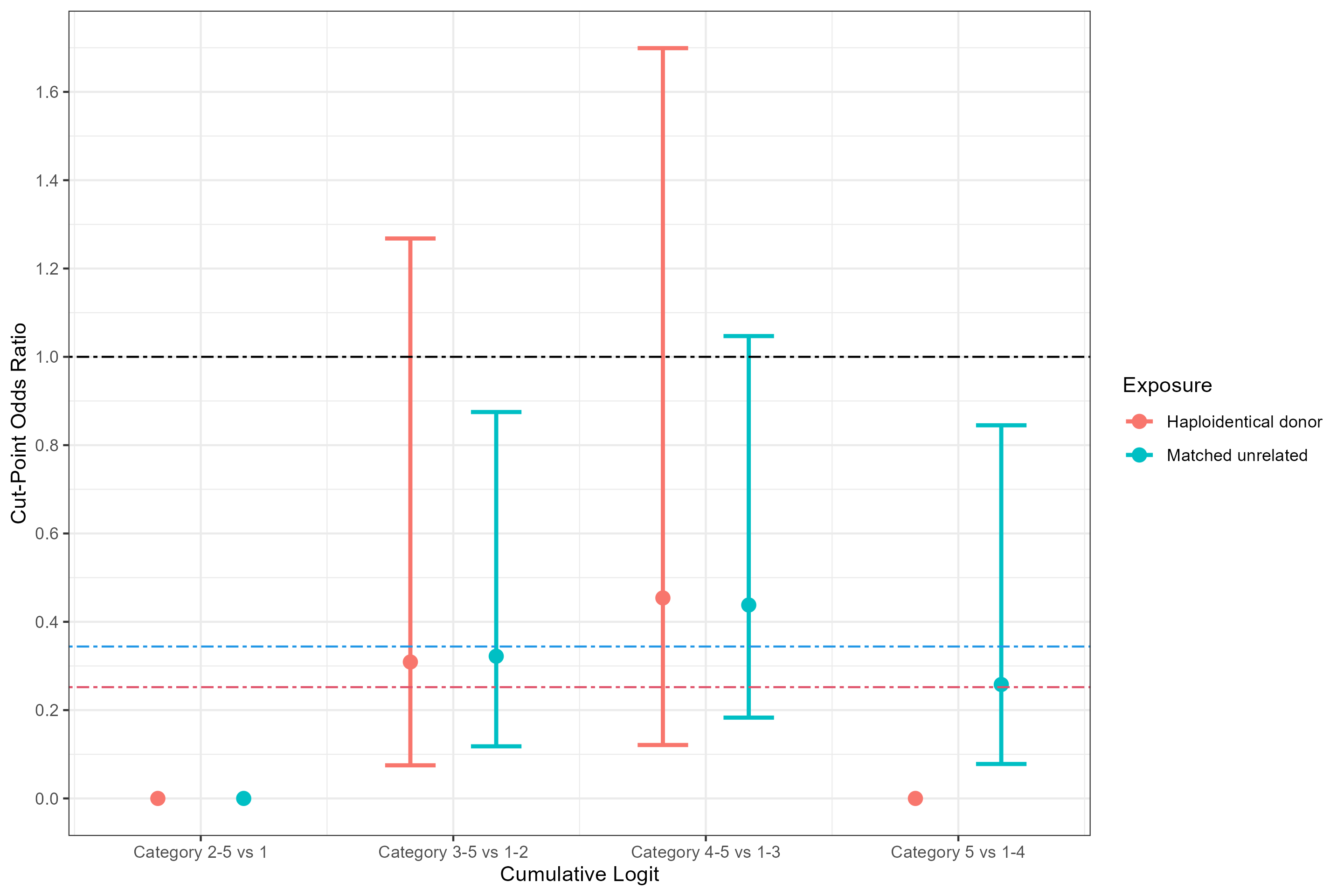

**Figure 7d:** Cut-point odds ratios for transplant donor type to assess proportional odds assumption for fourth ordinal endpoint (among those with malignant indication)

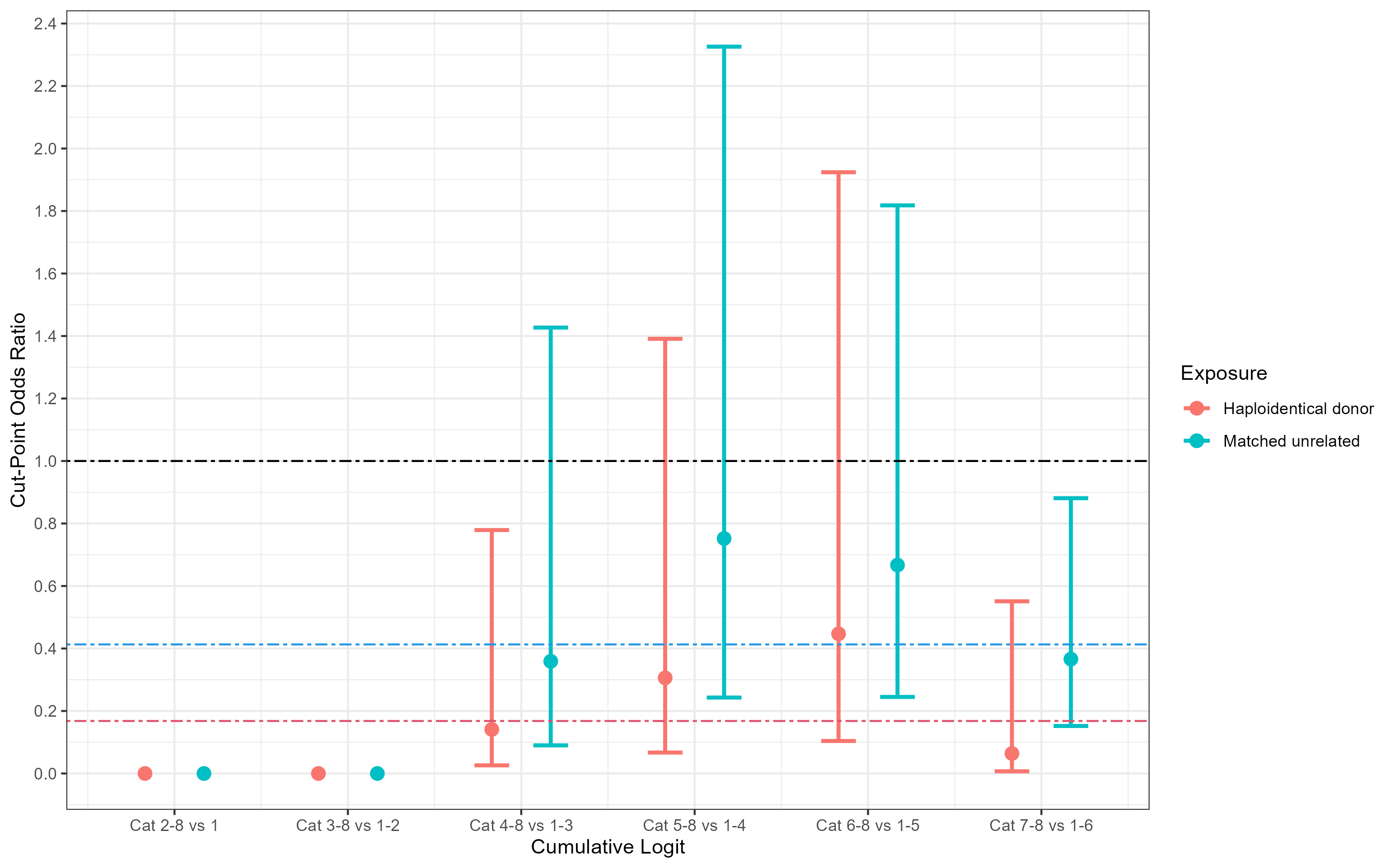

**Figure 8a:** Cut-point odds ratios for transplant donor type to assess proportional odds assumption for first ordinal endpoint (among those with non-malignant indication)

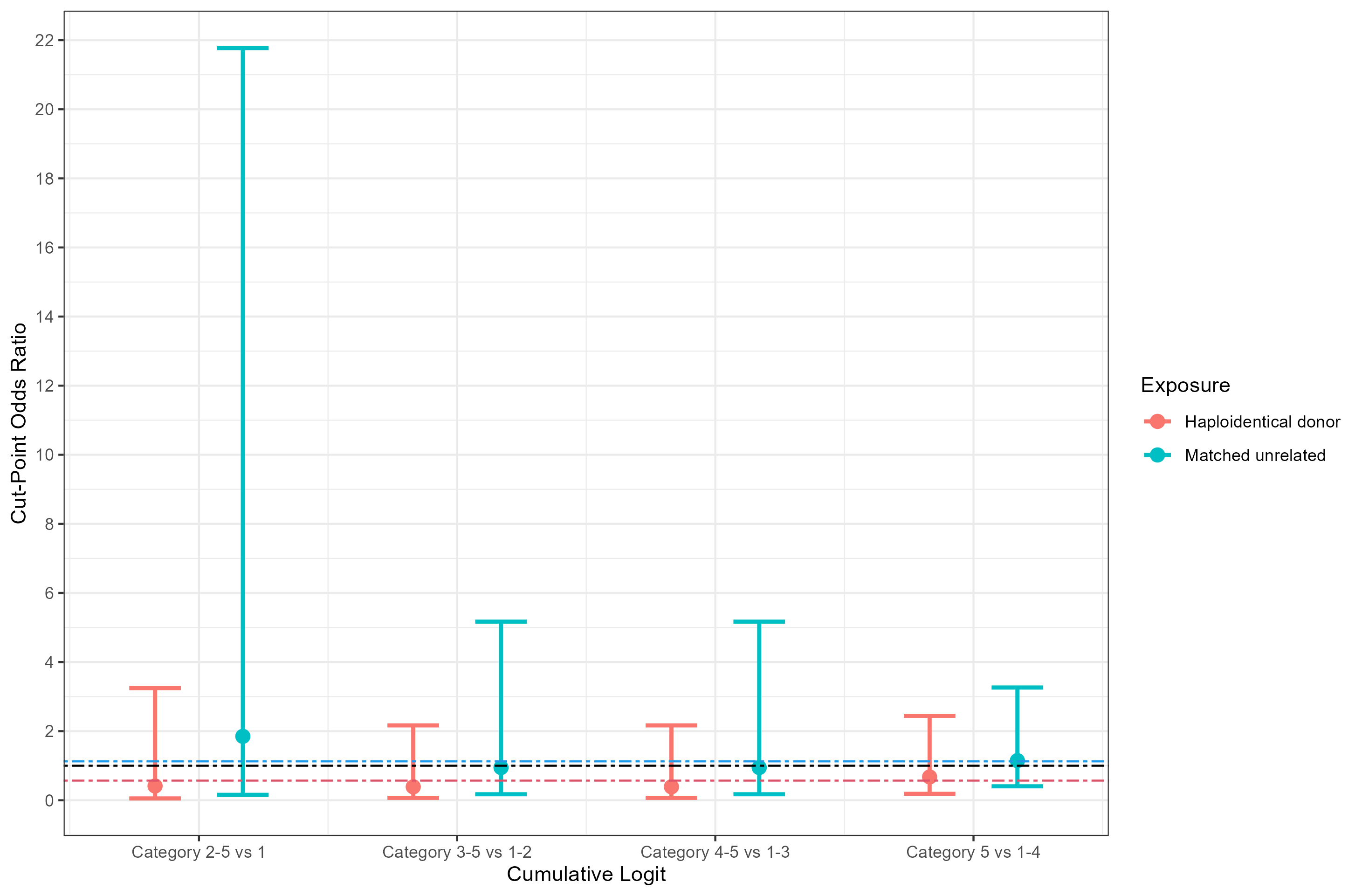

**Figure 8b:** Cut-point odds ratios for transplant donor type to assess proportional odds assumption for second ordinal endpoint (among those with non-malignant indication)

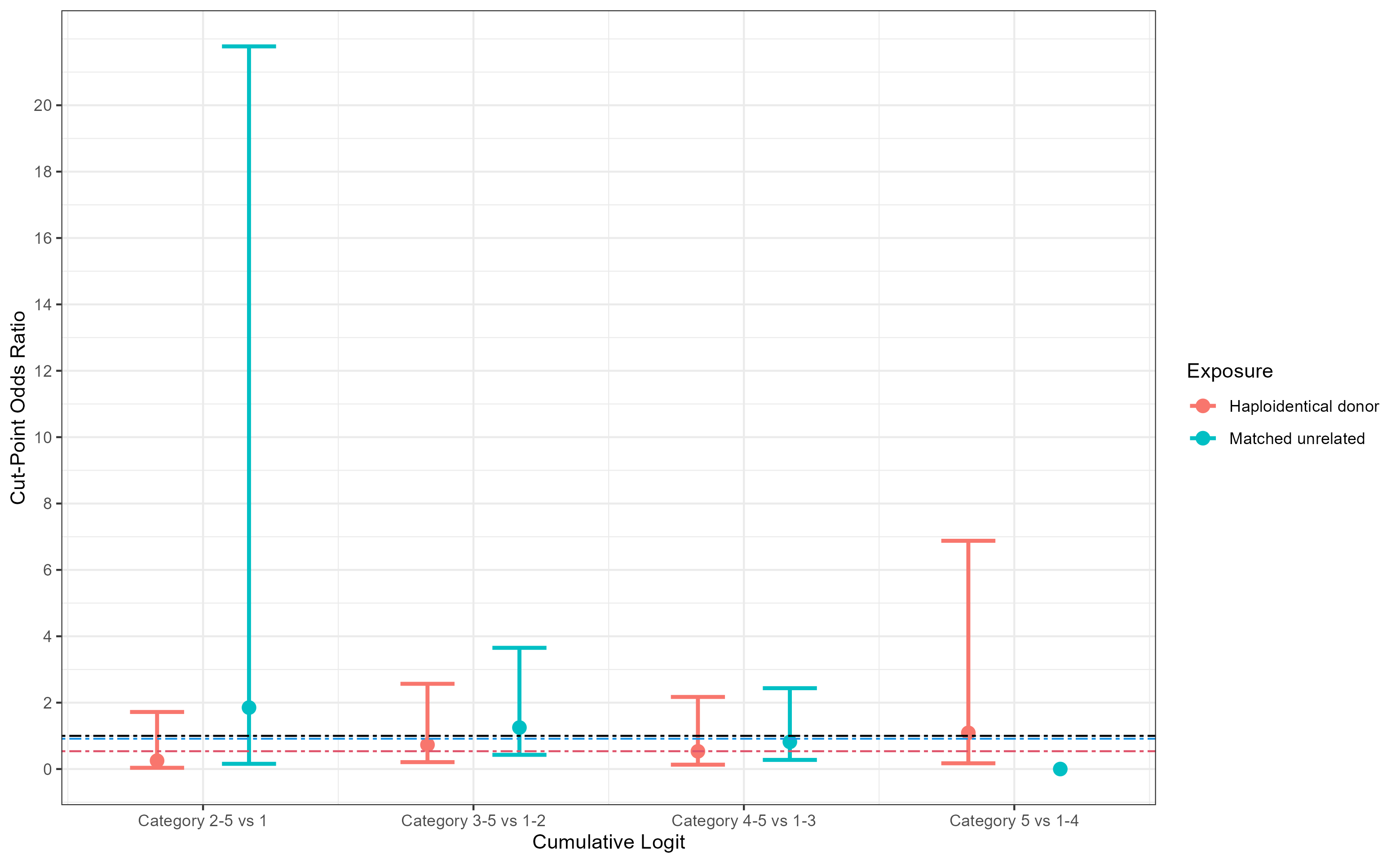

**Figure 8c:** Cut-point odds ratios for transplant donor type to assess proportional odds assumption for third ordinal endpoint (among those with non-malignant indication)

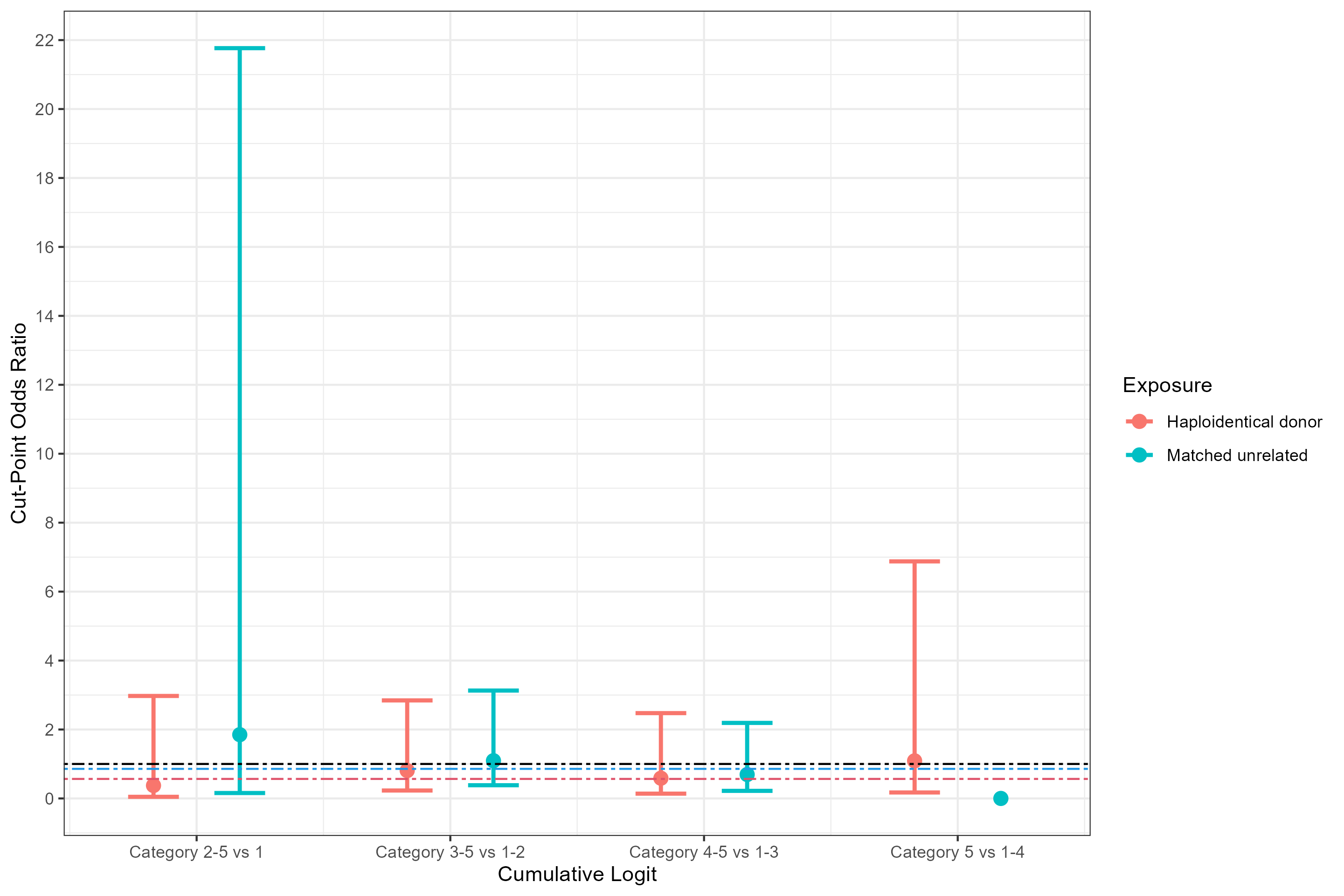

**Figure 8d:** Cut-point odds ratios for transplant donor type to assess proportional odds assumption for fourth ordinal endpoint (among those with non-malignant indication)

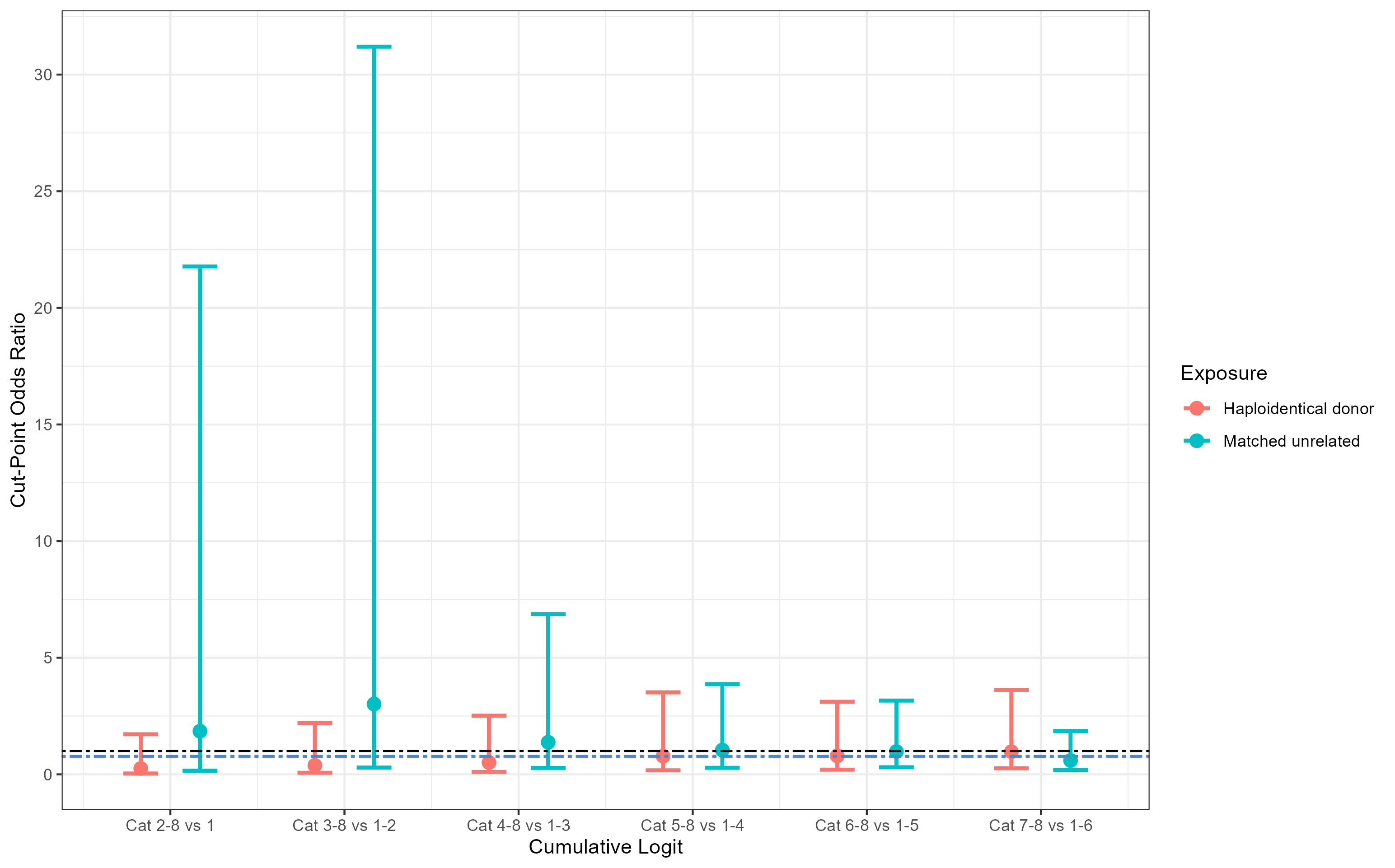

**Figure 9a:** Cut-point odds ratios for malignant indication of HSCT to assess proportional odds assumption for first ordinal endpoint

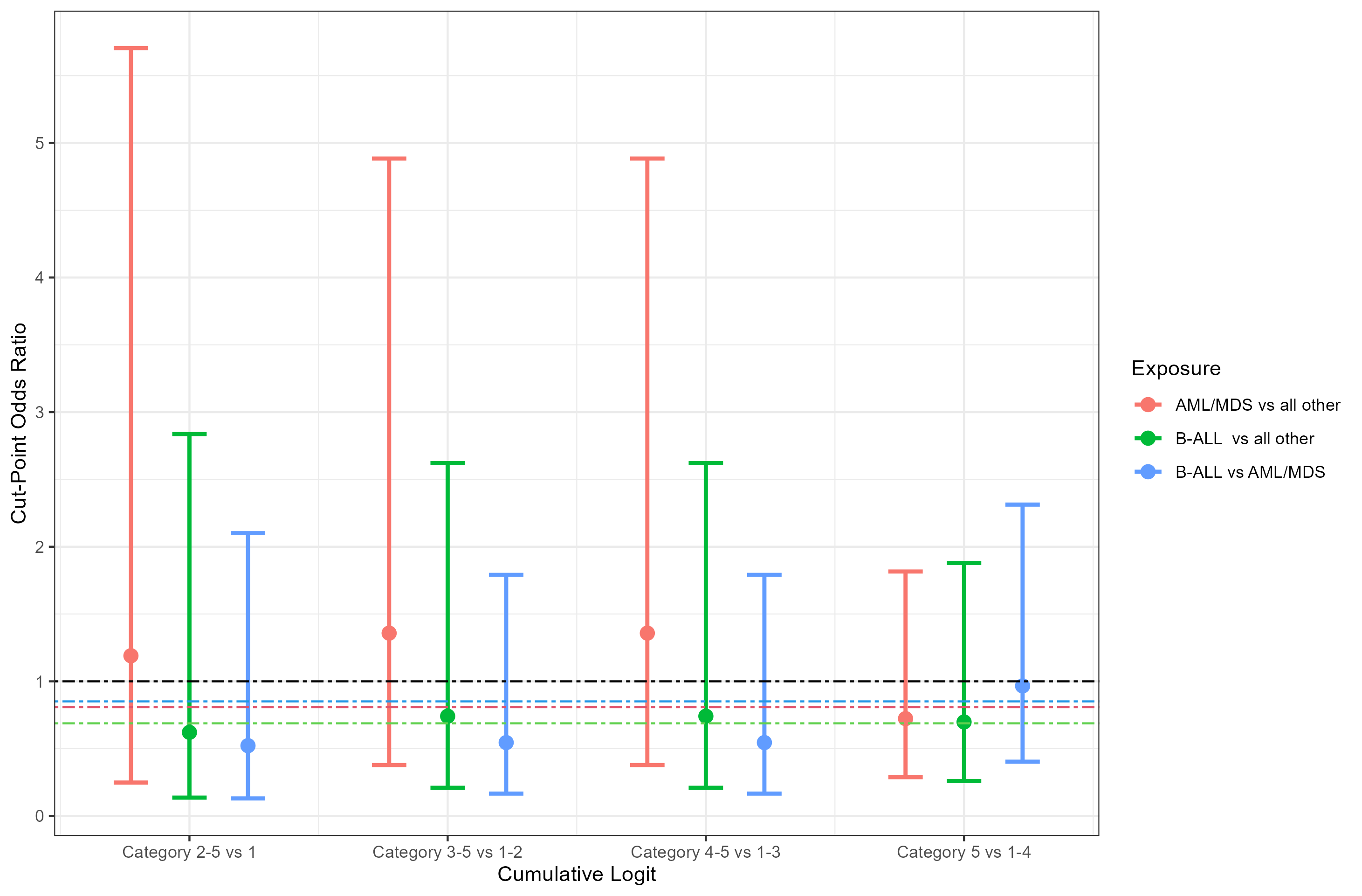

**Figure 9b:** Cut-point odds ratios for malignant indication of HSCT to assess proportional odds assumption for second ordinal endpoint

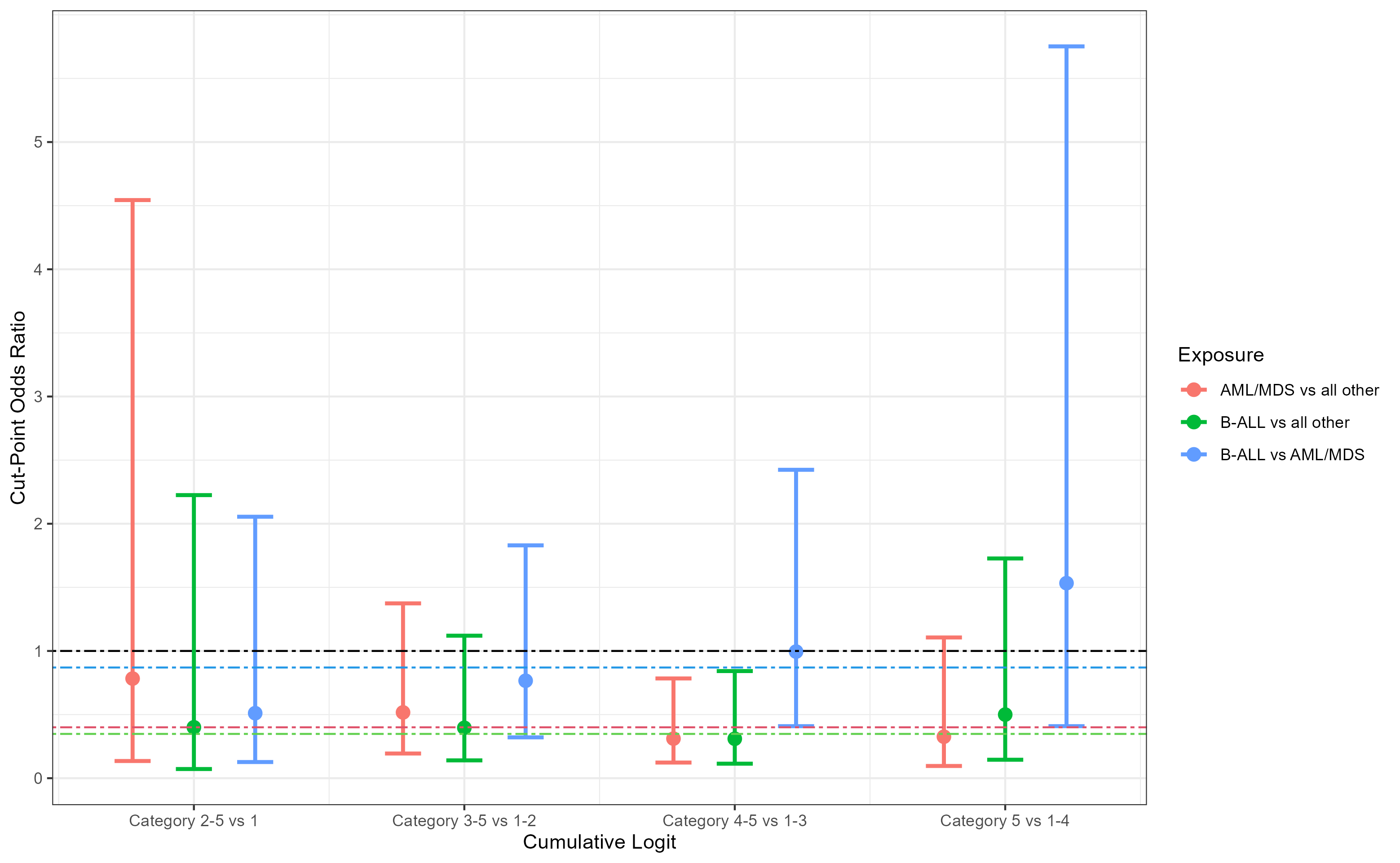

**Figure 9c:** Cut-point odds ratios for malignant indication of HSCT to assess proportional odds assumption for third ordinal endpoint

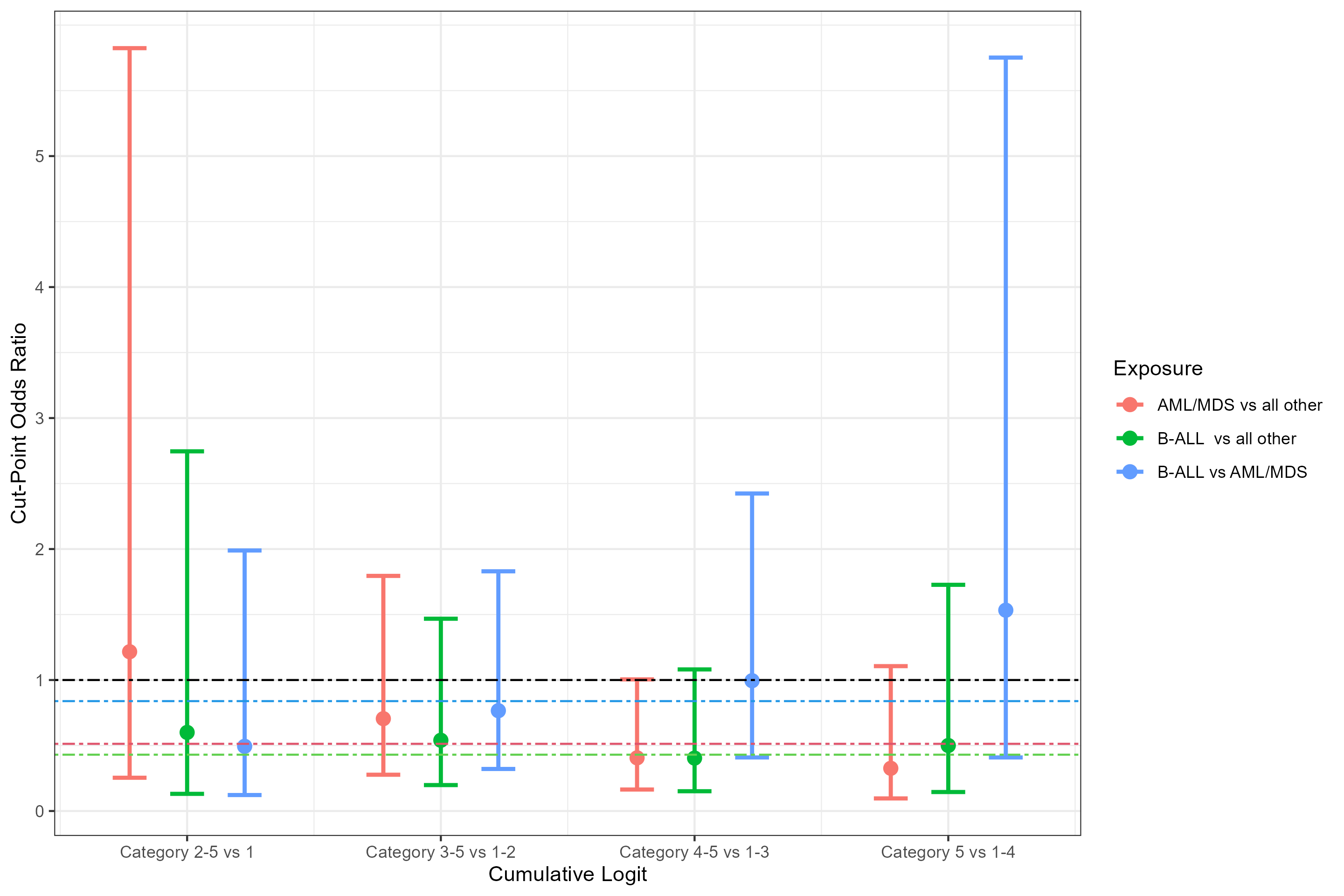

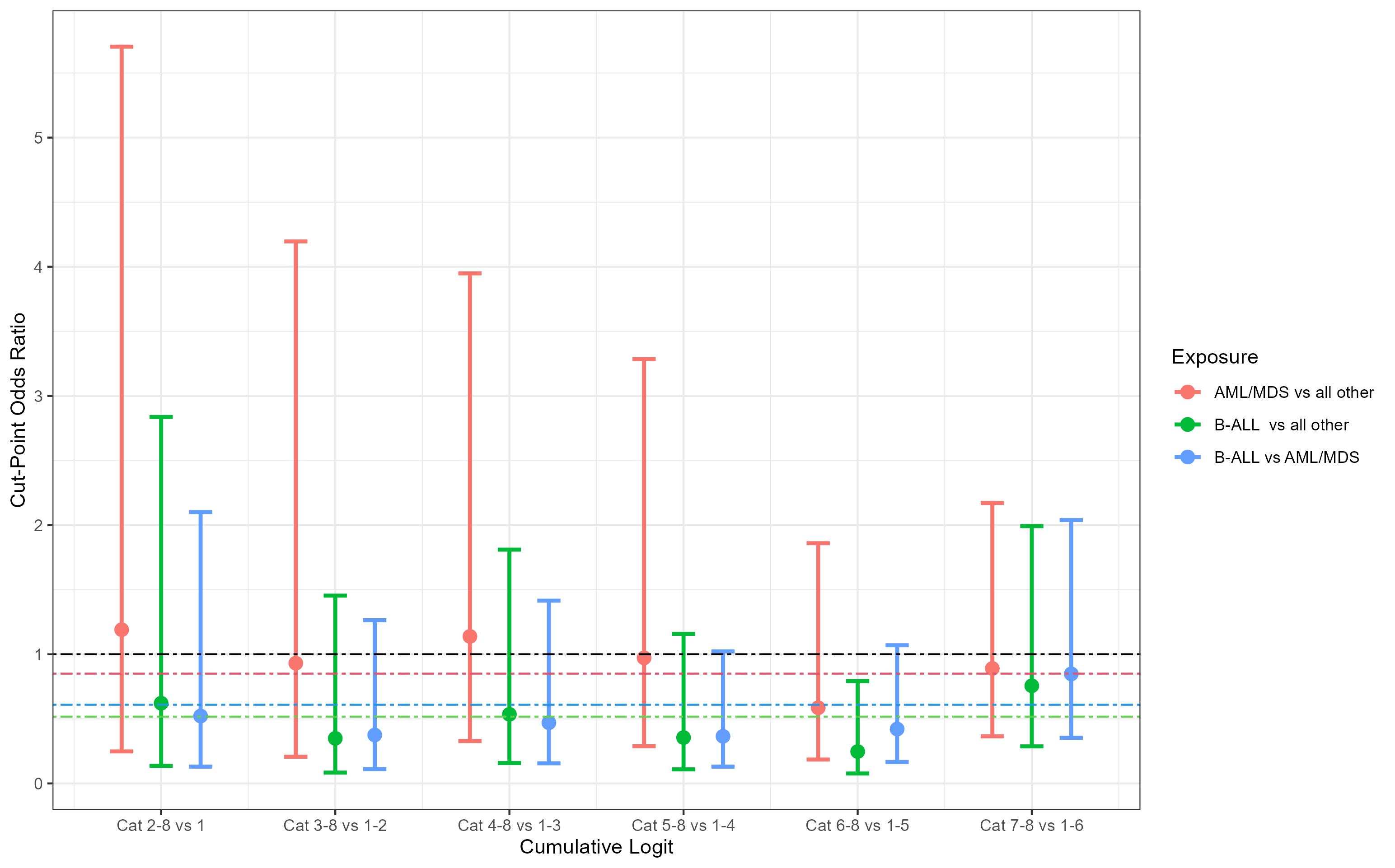
**Figure 9d:** Cut-point odds ratios for malignant indication of HSCT to assess proportional odds assumption for third ordinal endpoint

**Figure 10a:** Cut-point odds ratios for cord blood transplant status to assess proportional odds assumption for first ordinal endpoint

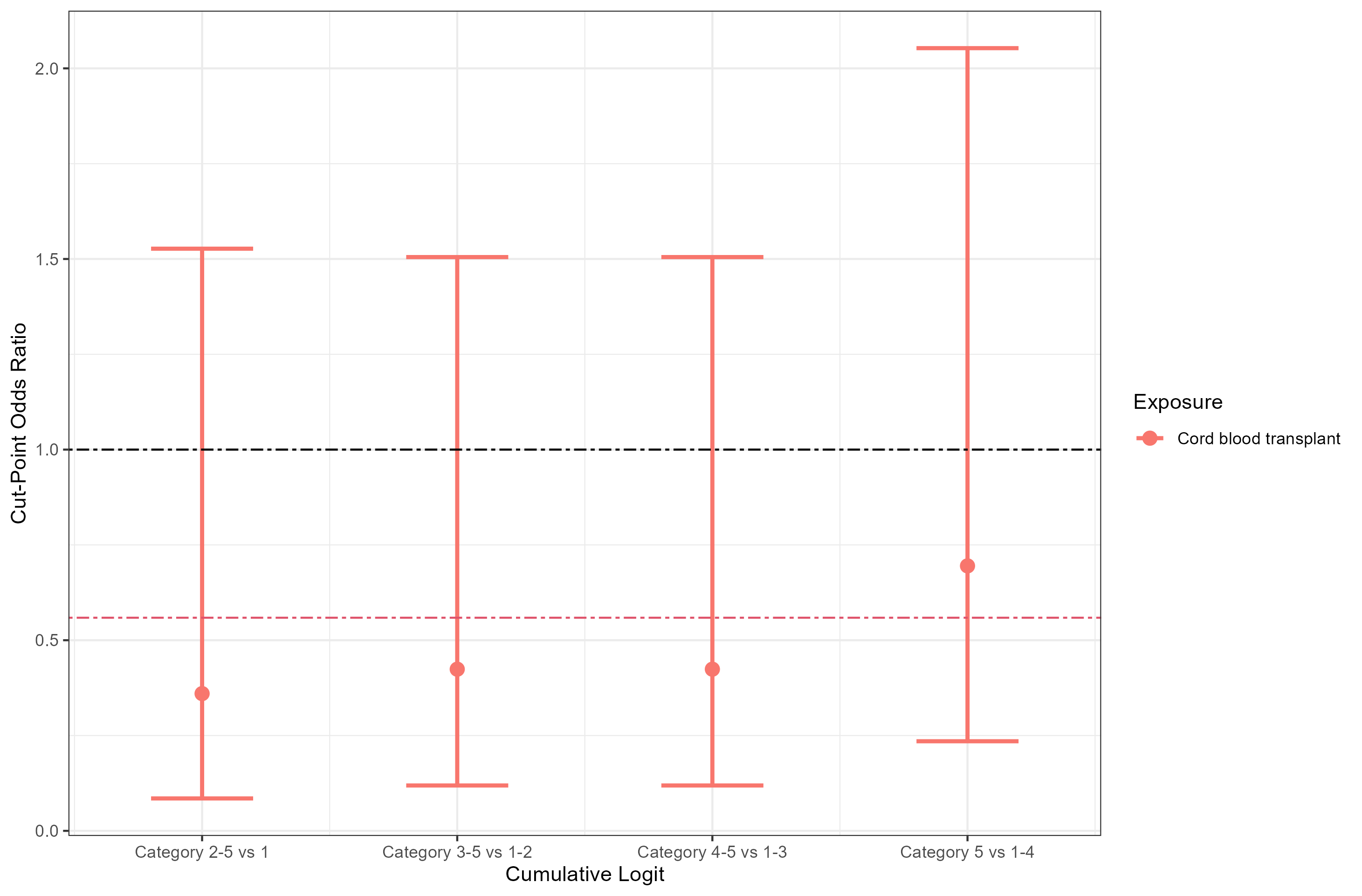

**Figure 10b:** Cut-point odds ratios for cord blood transplant status to assess proportional odds assumption for second ordinal endpoint

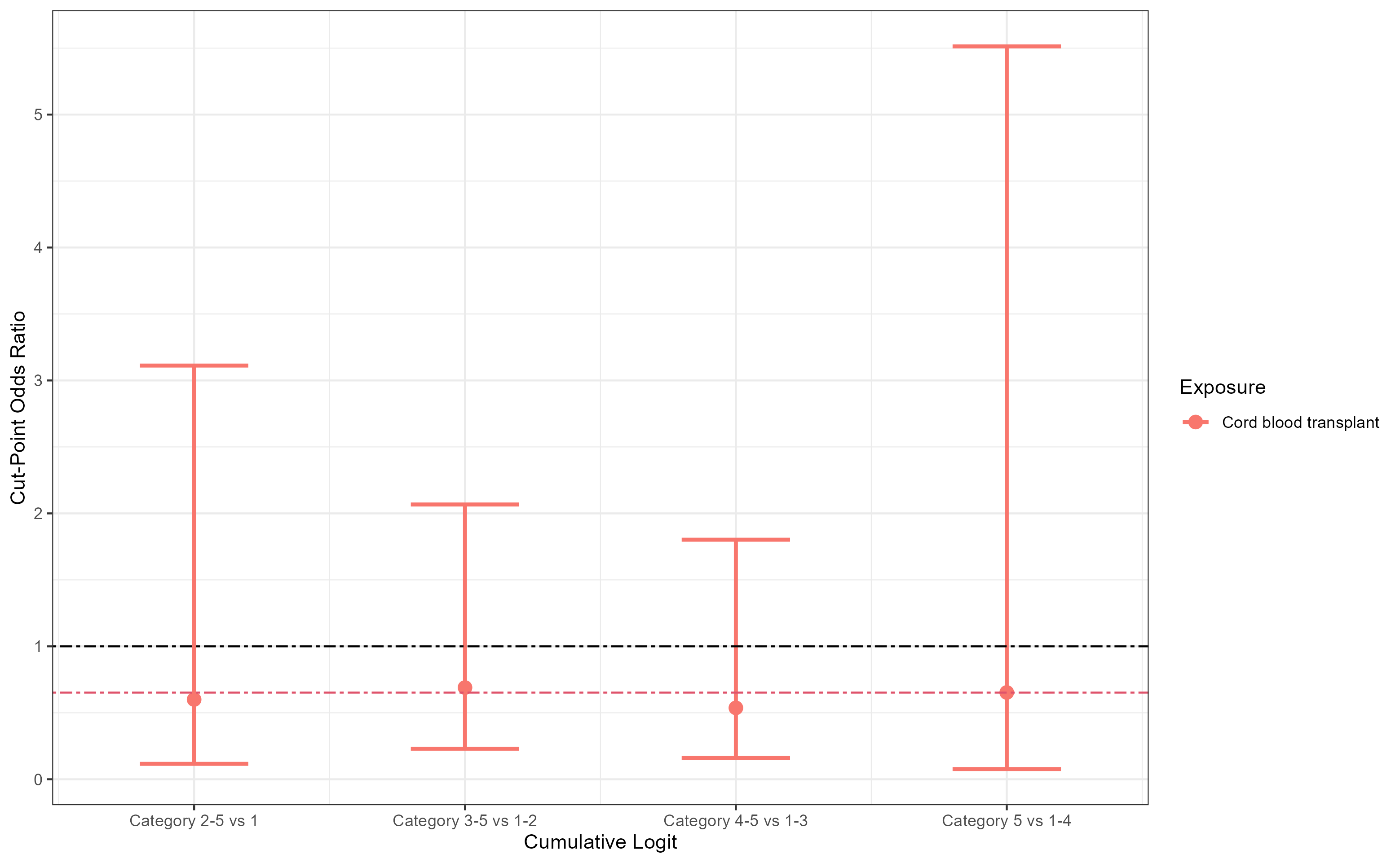

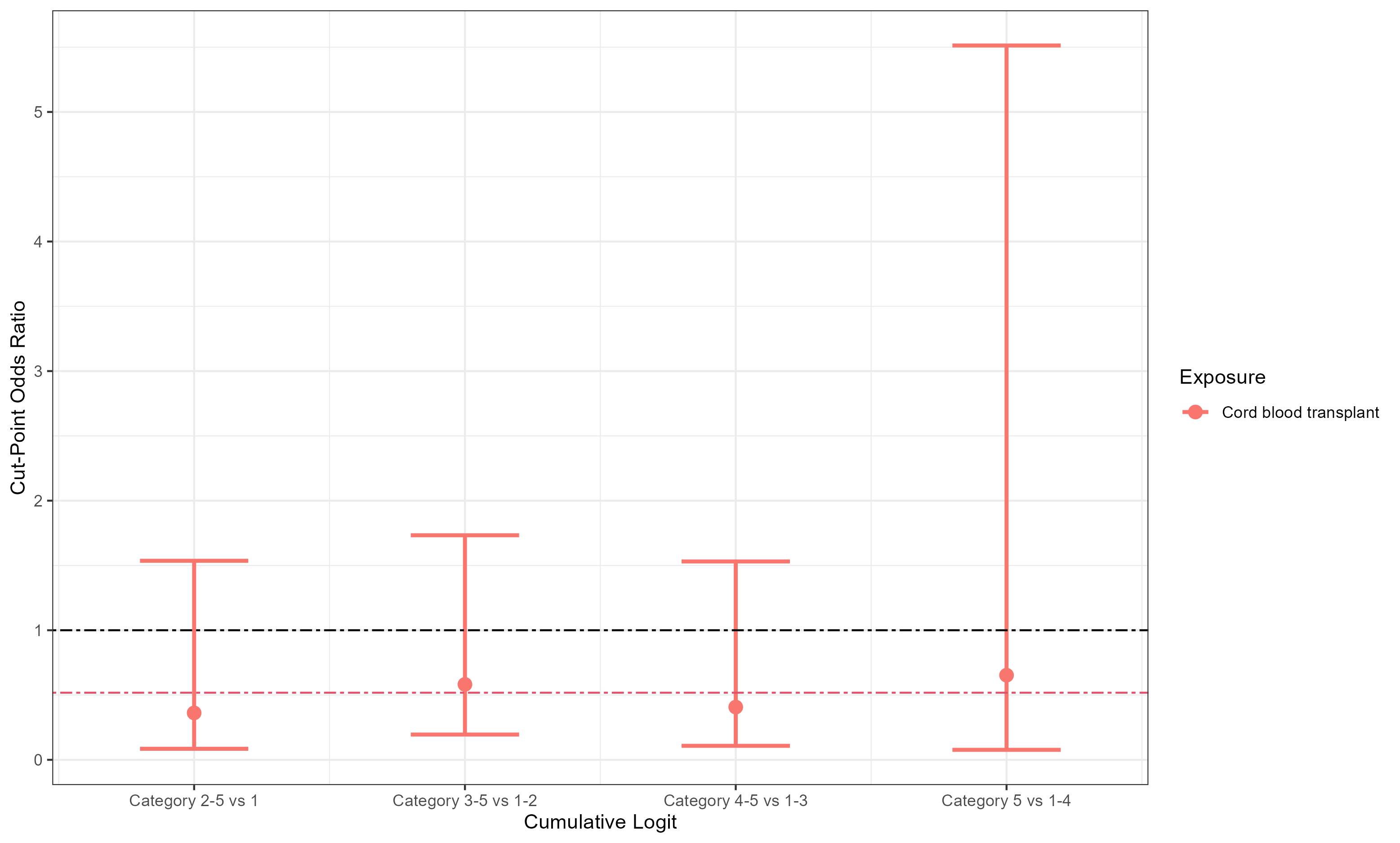
**Figure 10c:** Cut-point odds ratios for cord blood transplant status to assess proportional odds assumption for third ordinal endpoint

**Figure 10d:** Cut-point odds ratios for cord blood transplant status to assess proportional odds assumption for fourth ordinal endpoint

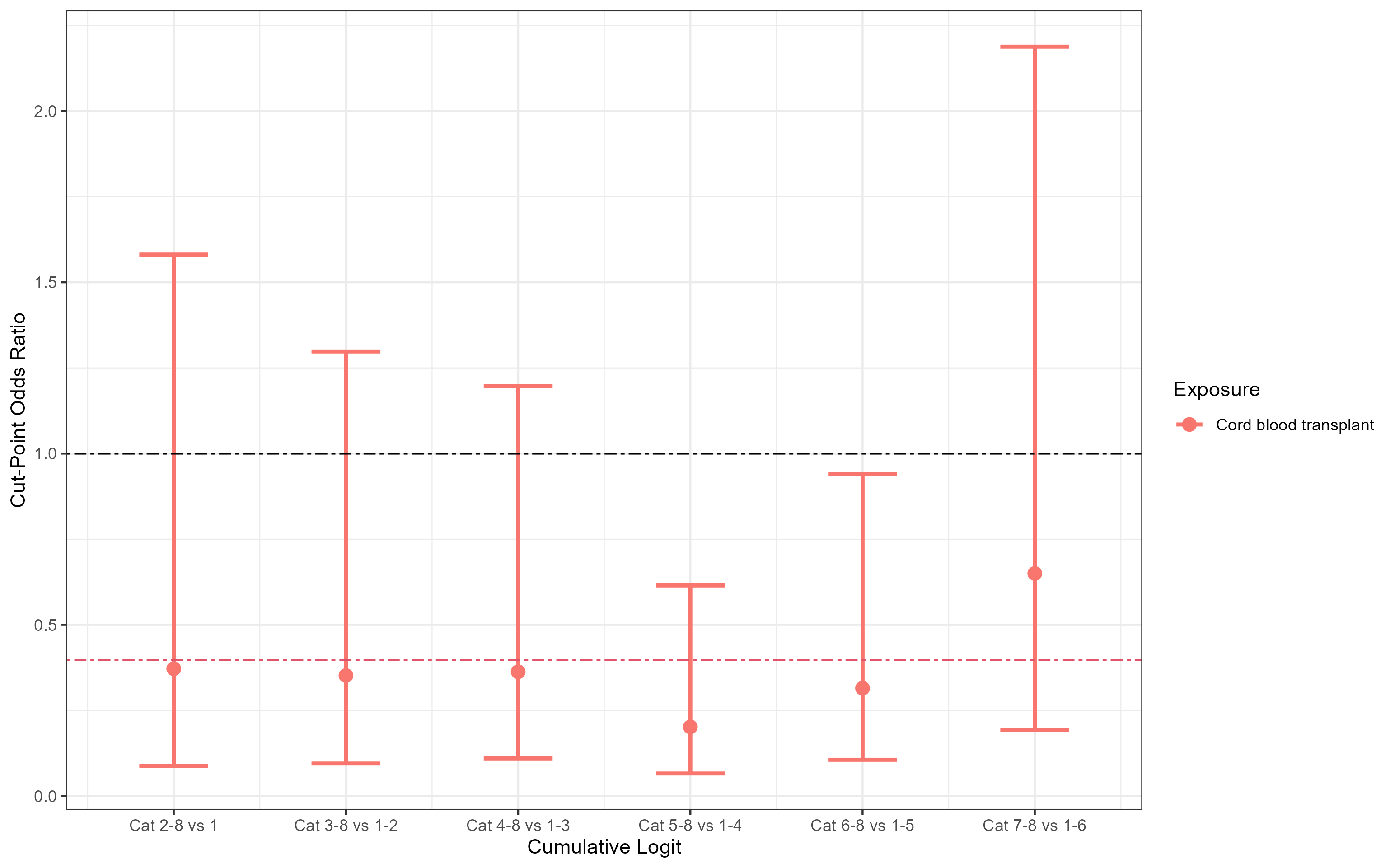

**Figure 11a:** Cut-point odds ratios for presence of MRD or not among patients with B-ALL to assess proportional odds assumption for first ordinal endpoint

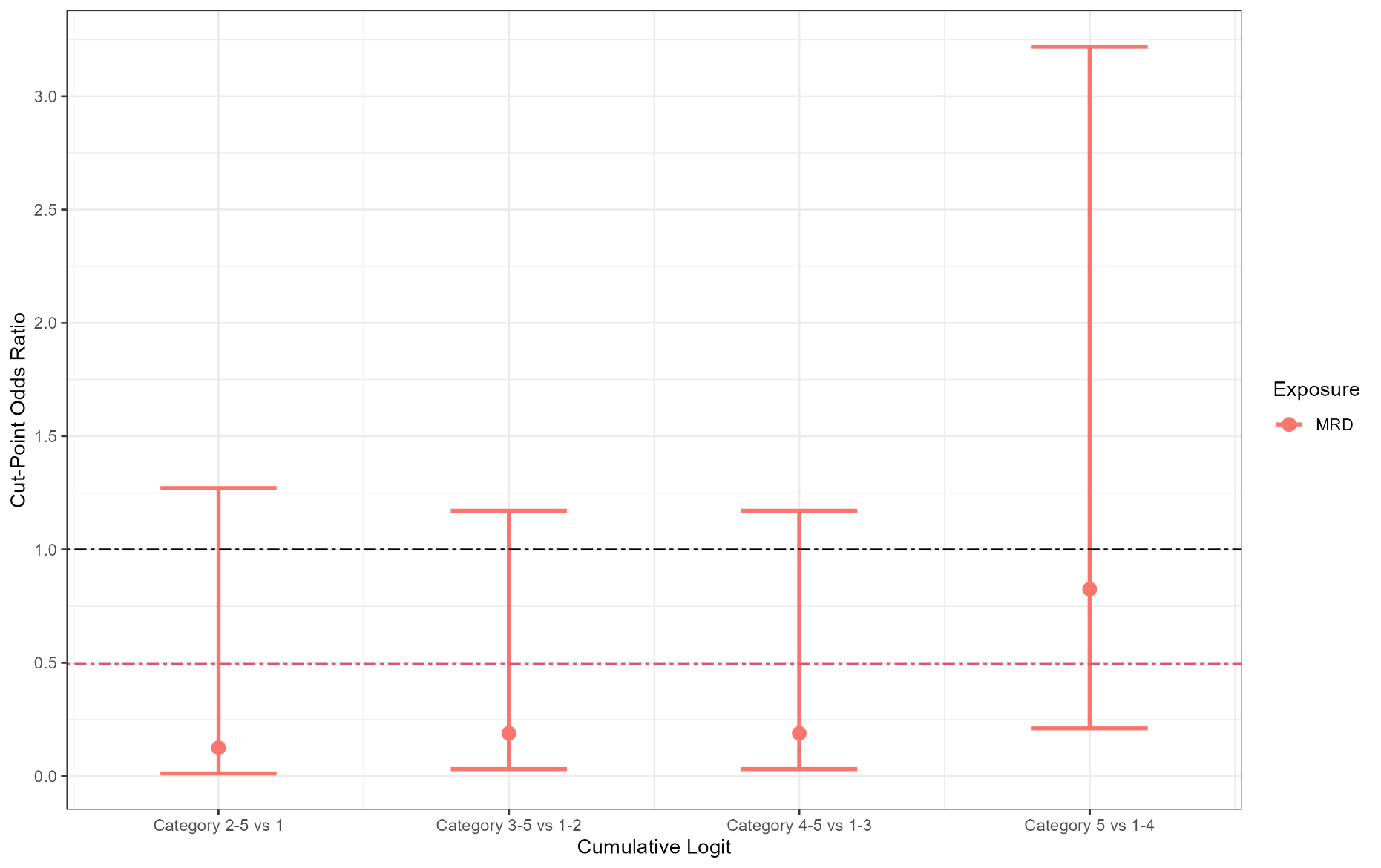

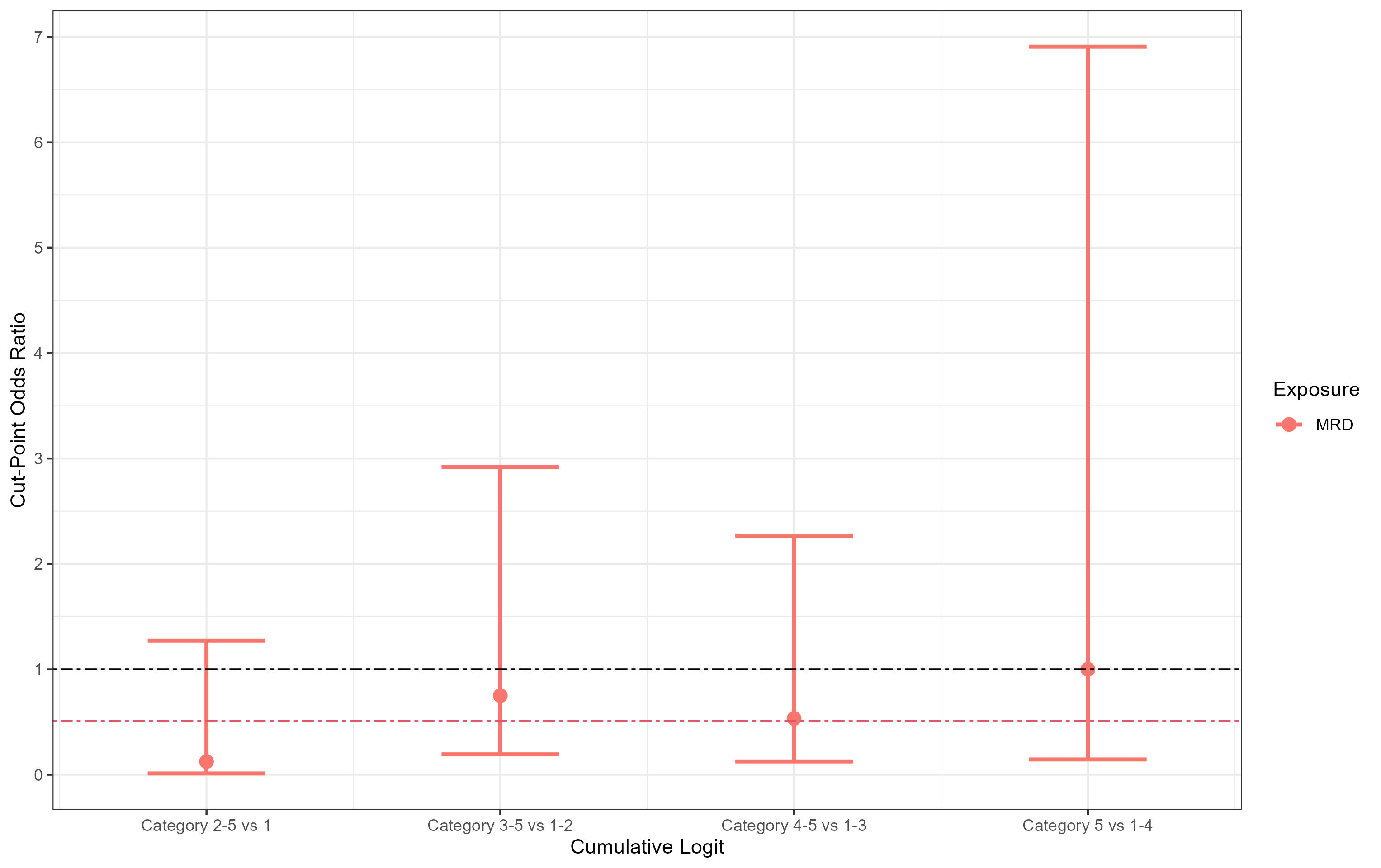
**Figure 11b:** Cut-point odds ratios for presence of MRD or not among patients with B-ALL to assess proportional odds assumption for second ordinal endpoint

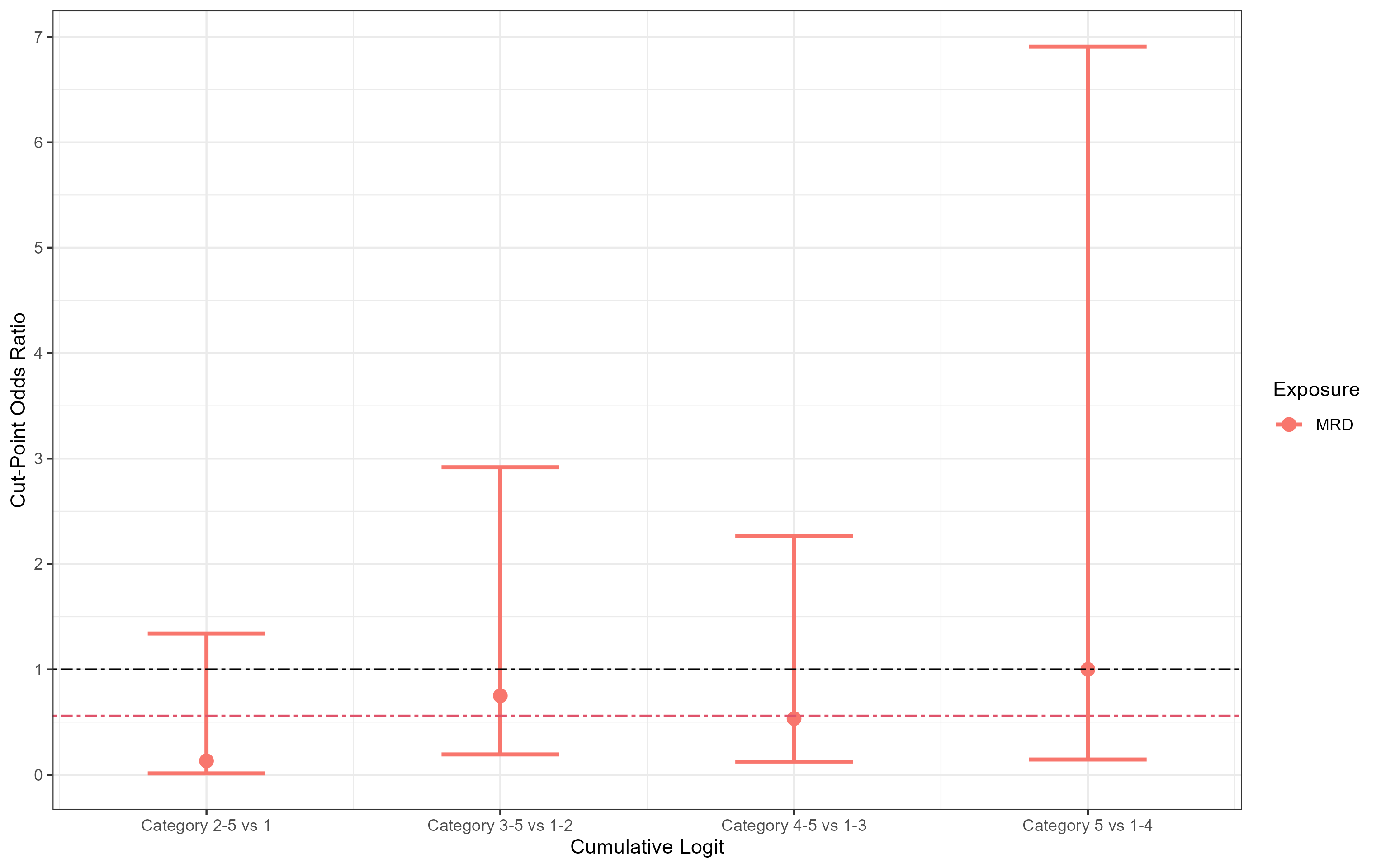
**Figure 11c:** Cut-point odds ratios for presence of MRD or not among patients with B-ALL to assess proportional odds assumption for third ordinal endpoint

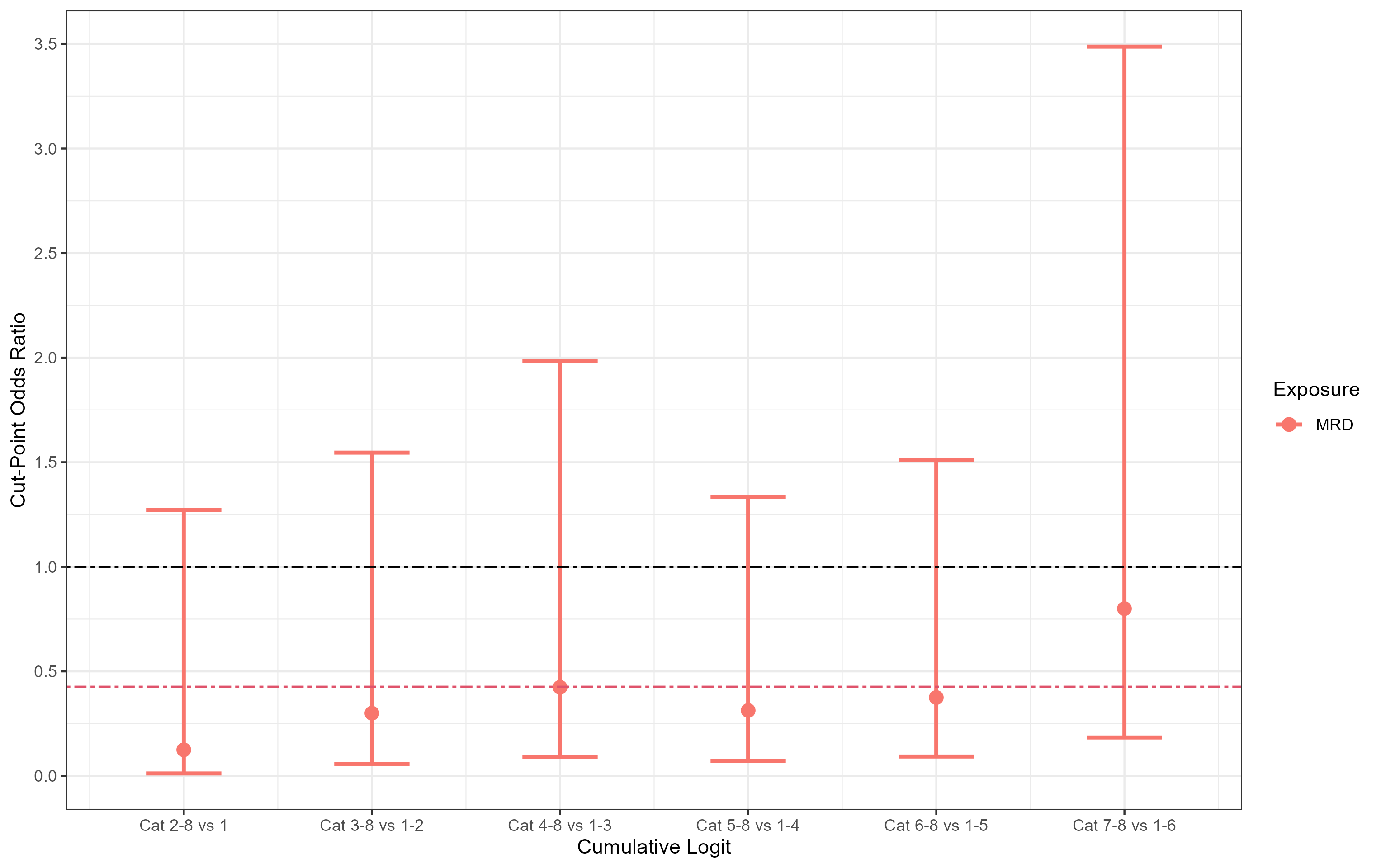
**Figure 11d:** Cut-point odds ratios for presence of MRD or not among patients with B-ALL to assess proportional odds assumption for fourth ordinal endpoint

**SUPPLEMENTARY MATERIAL 5: Cumulative Distributions of Proposed Endpoints by Exposure Group**

In the following section, we present the percentages that fall in each category by exposure group in Grotta bar charts. When interpreting the Grotta bar charts, if the black lines between exposure groups are consistently in the same direction (and similar angle), this is consistent with the proportional odds assumption.

1. ***CMV mis-matched donor recipient***

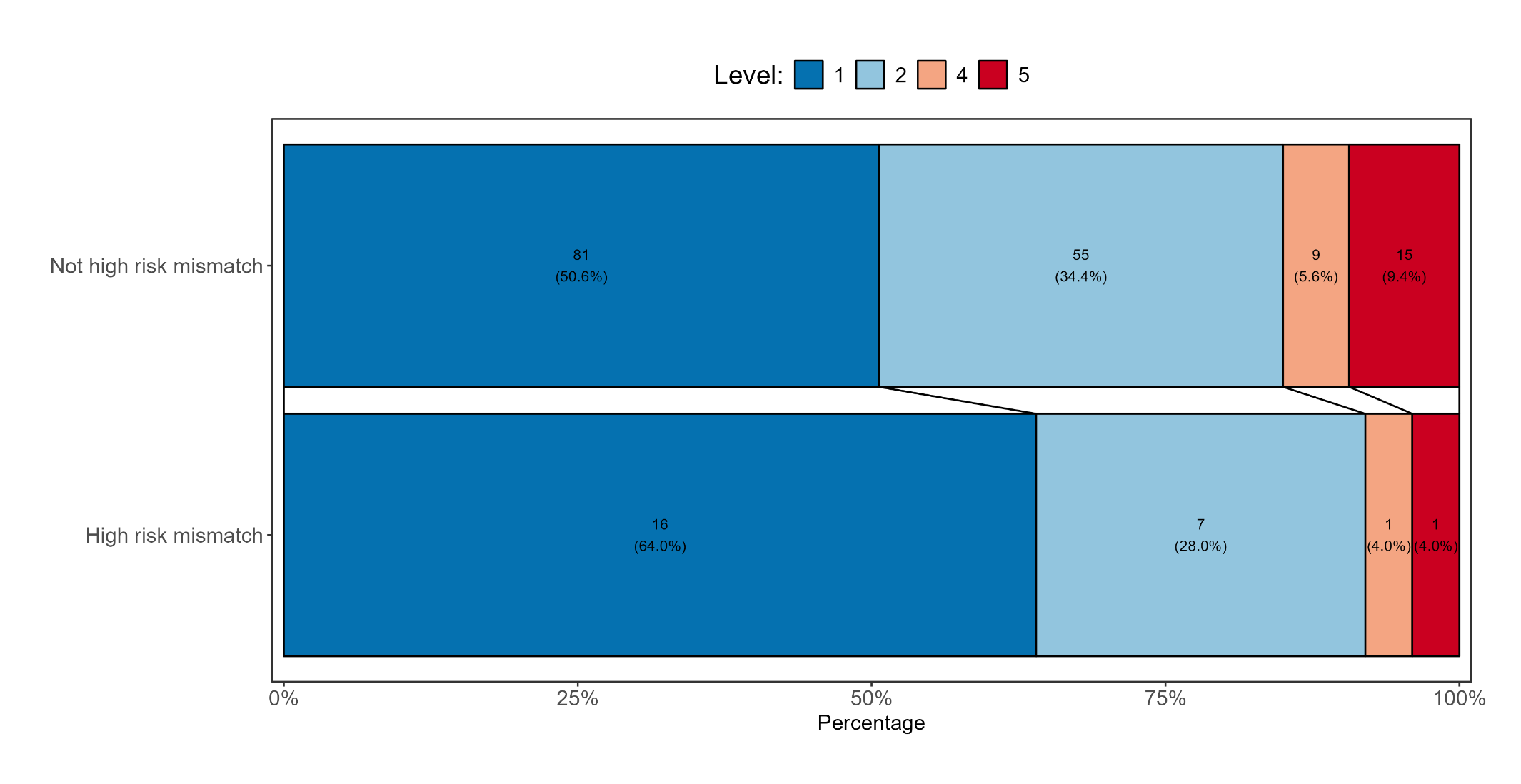
**Figure 12a:** Grotta bar for first ordinal endpoint by CMV mismatched status

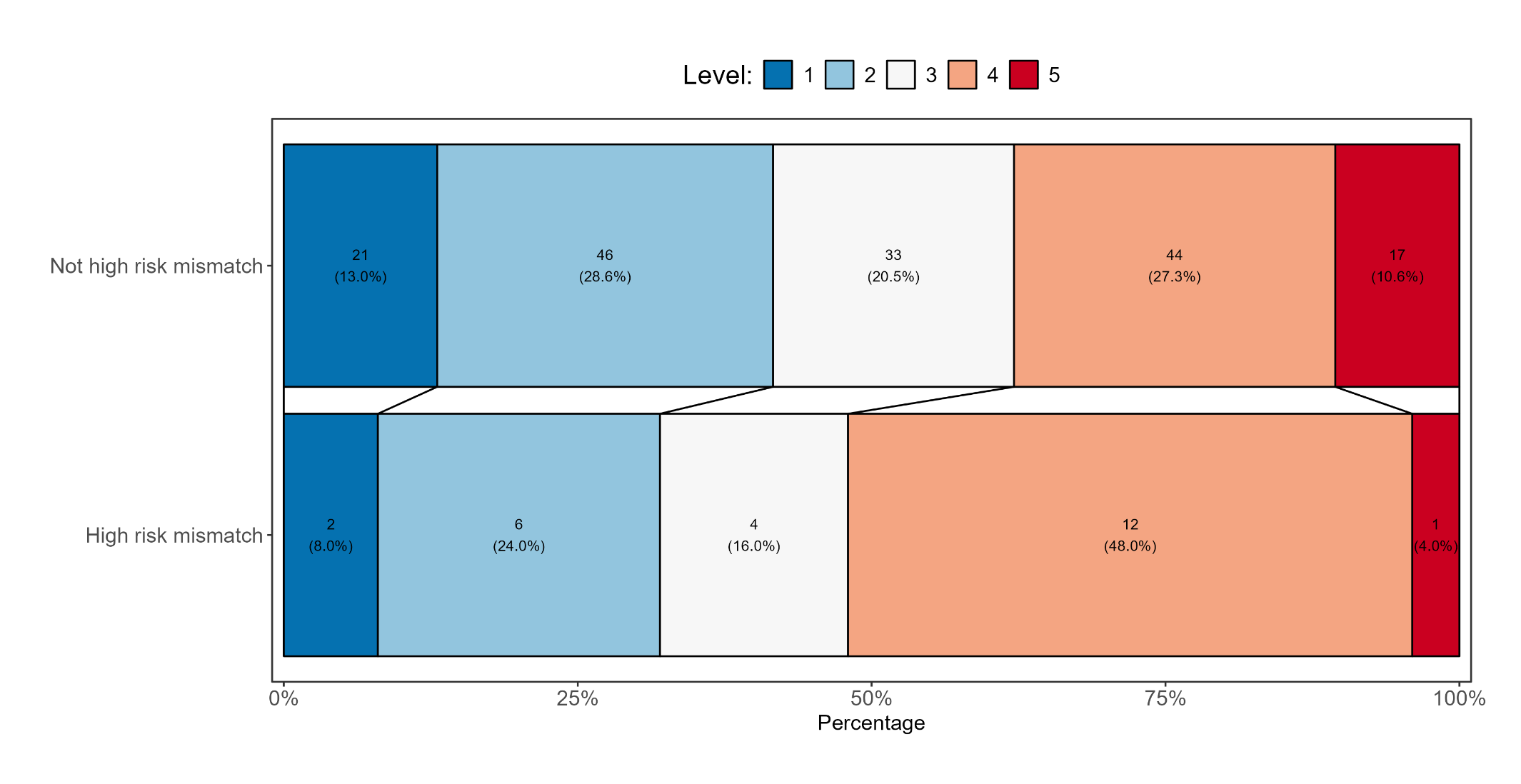
**Figure 12b:** Grotta bar for second ordinal endpoint by CMV mismatched status

**Figure 12c:** Grotta bar for third ordinal endpoint by CMV mismatched status

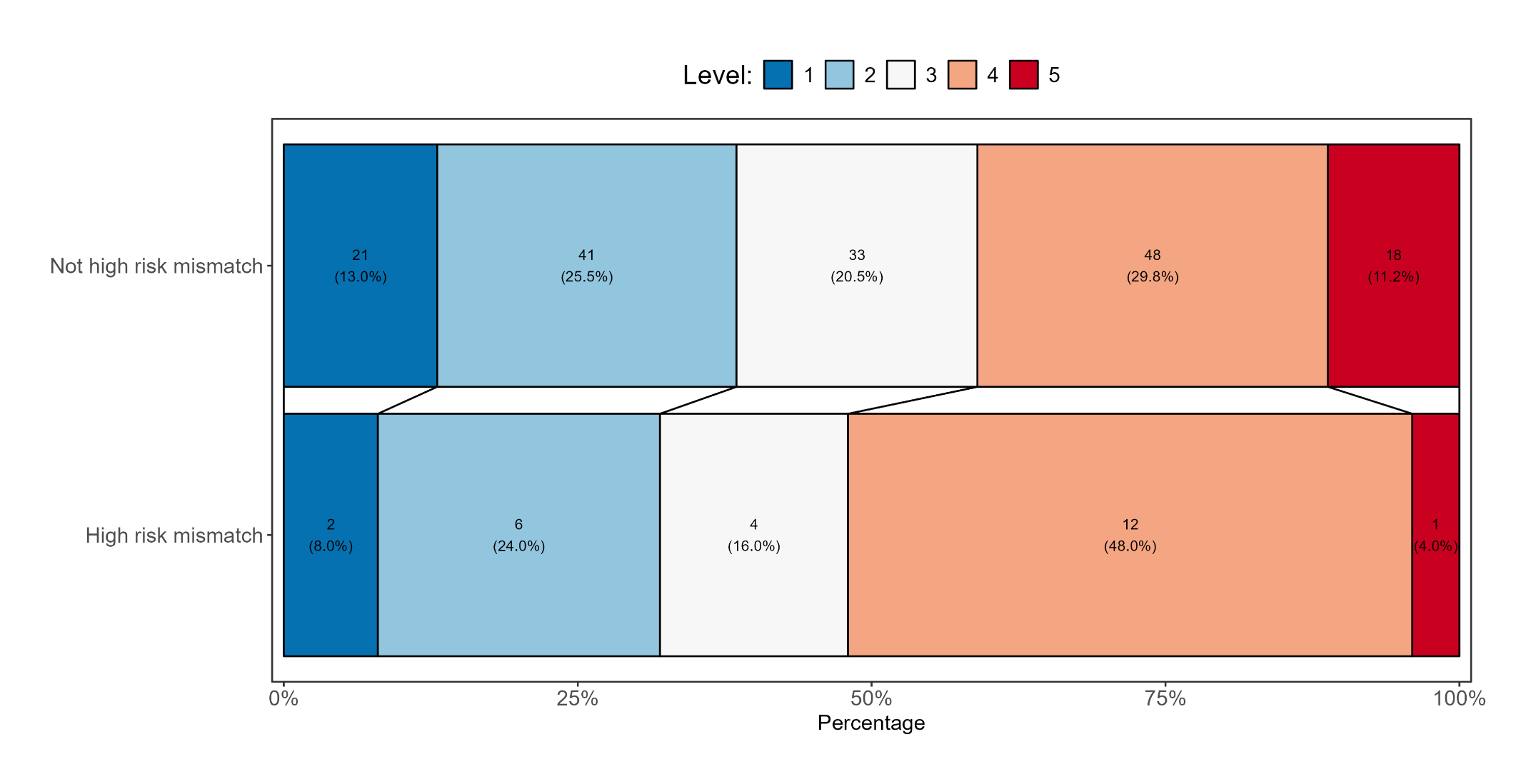

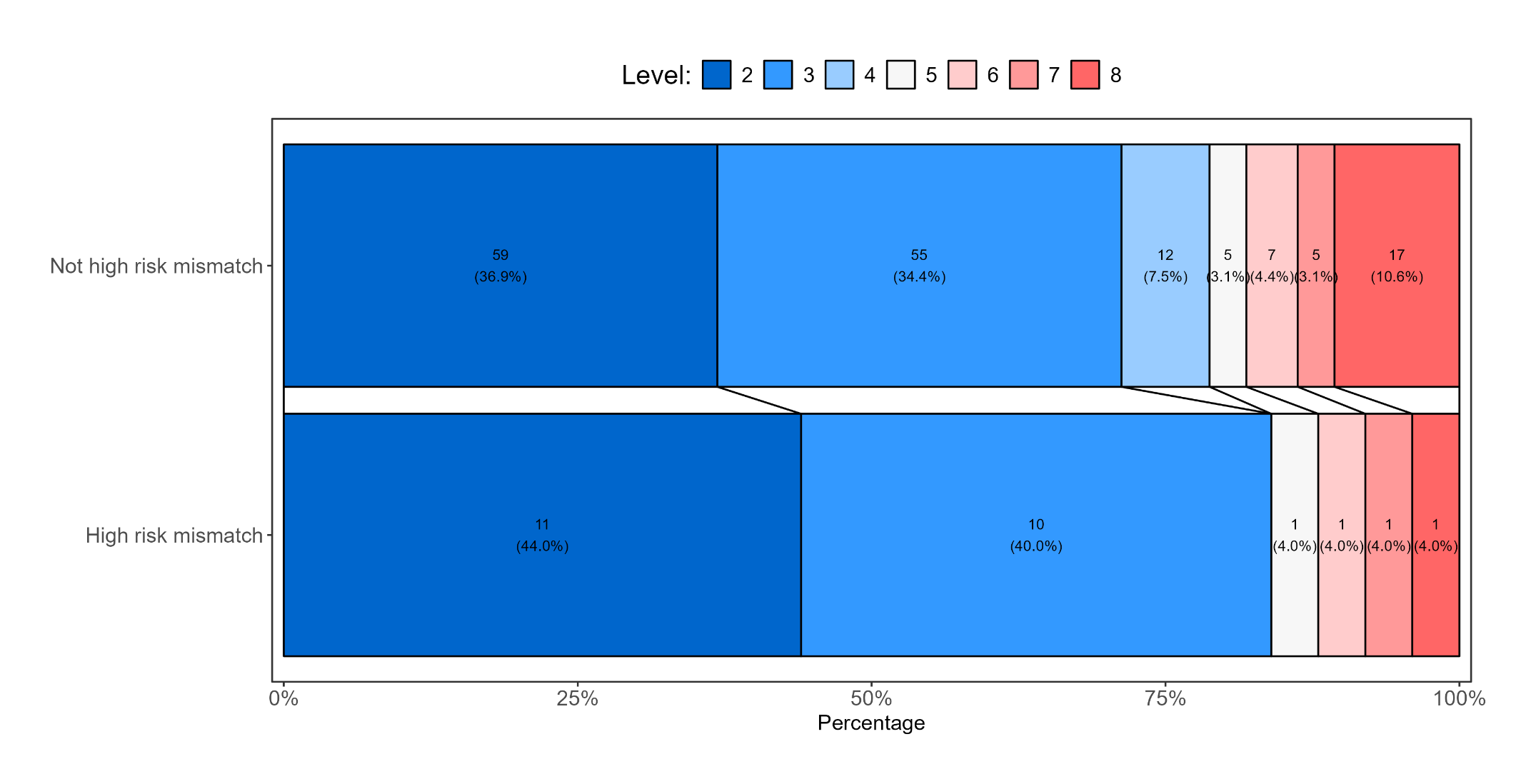
**Figure 12d:** Grotta bar for fourth ordinal endpoint by CMV mismatched status

2. ***Transplant type (for those with malignant indications)***

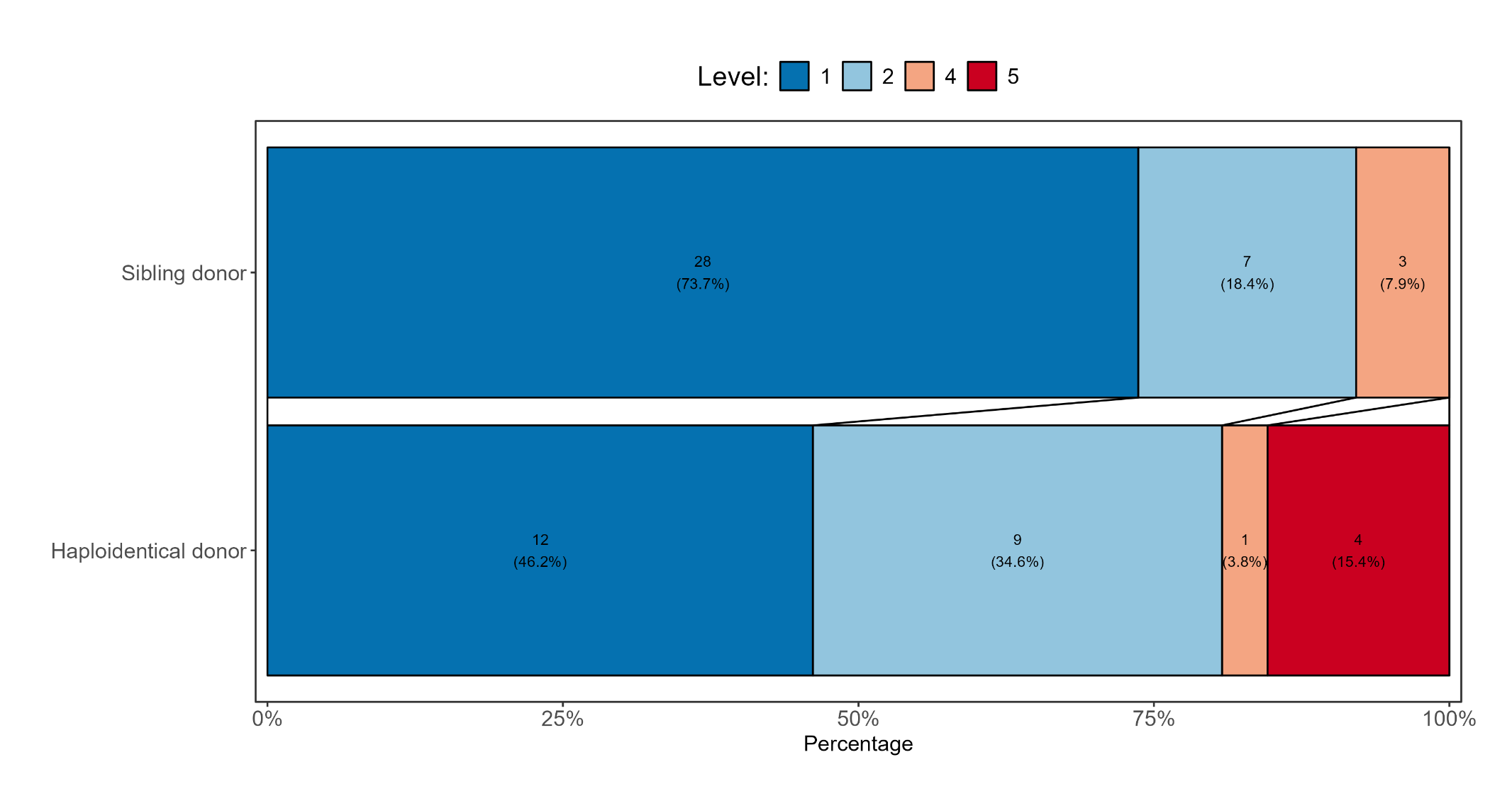
**Figure 13a:** Grotta bar for first ordinal endpoint by transplant type (haploidentical vs sibling donor)

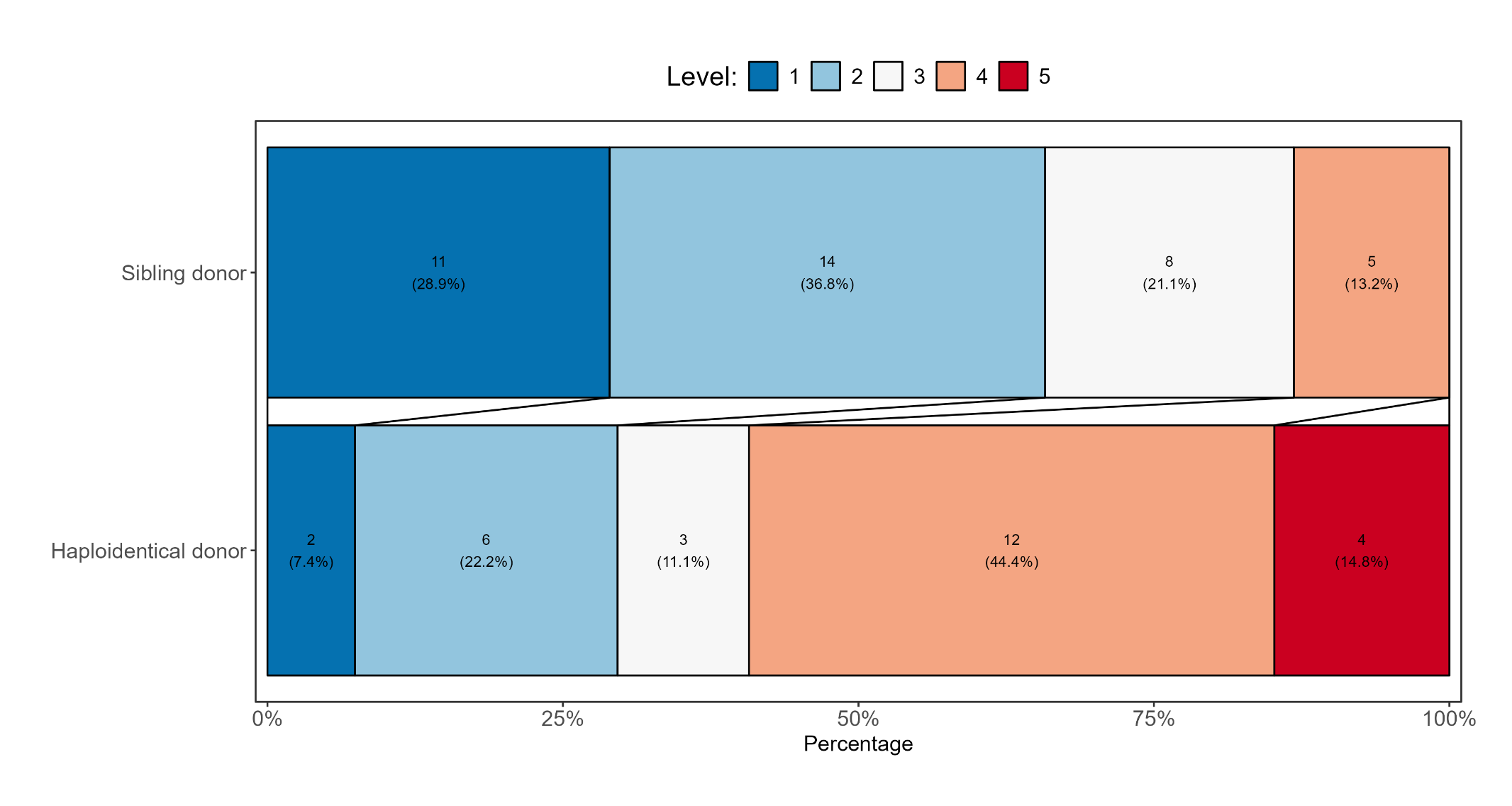
**Figure 13b:** Grotta bar for second ordinal endpoint by transplant type (haploidentical vs sibling donor)

**Figure 13c:** Grotta bar for third ordinal endpoint by transplant type (haploidentical vs sibling donor)

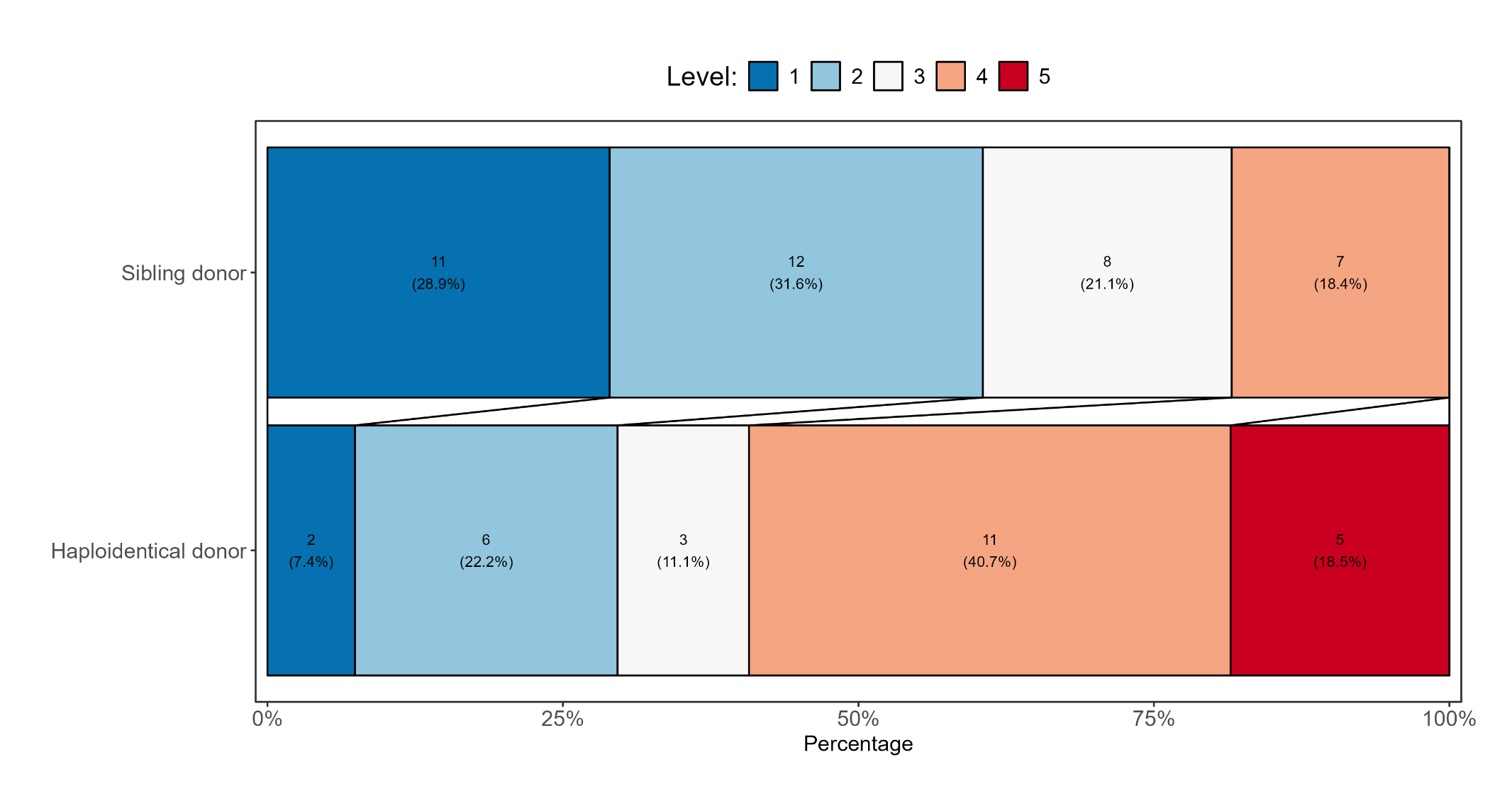

**Figure 13d:** Grotta bar for fourth ordinal endpoint by transplant type (haploidentical vs sibling donor)

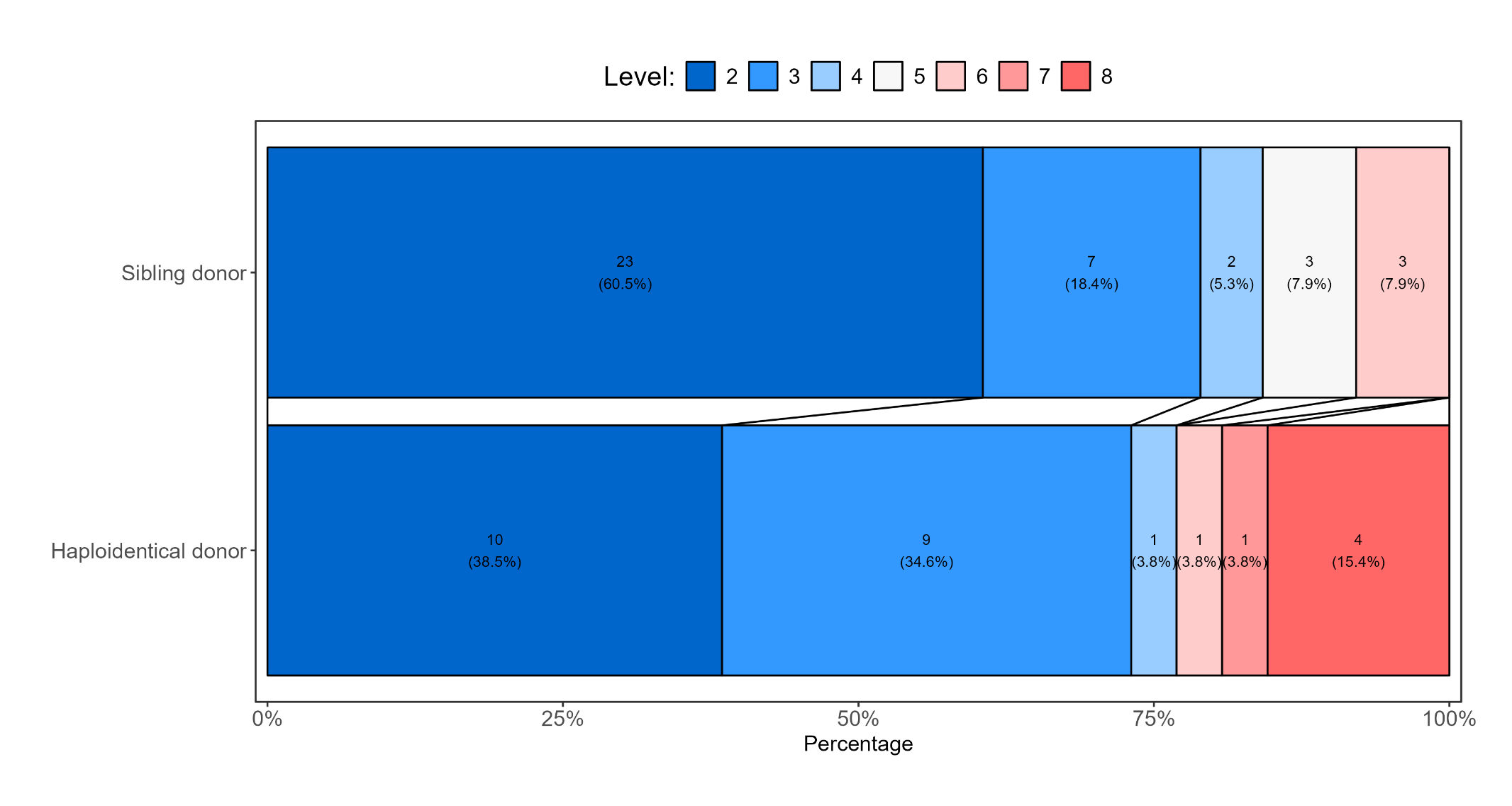

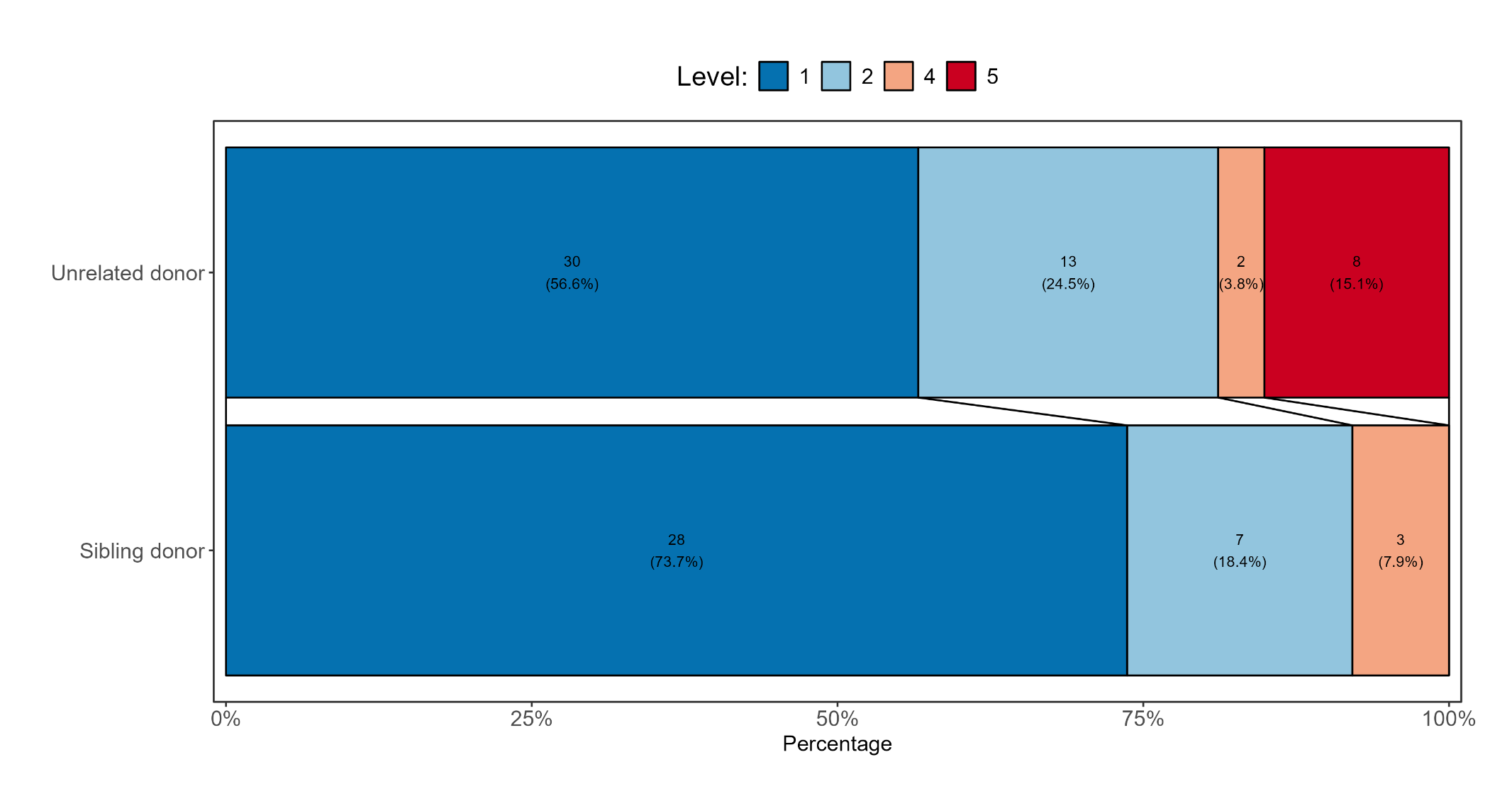
**Figure 13e:** Grotta bar for first ordinal endpoint by transplant type (unrelated vs sibling donor)

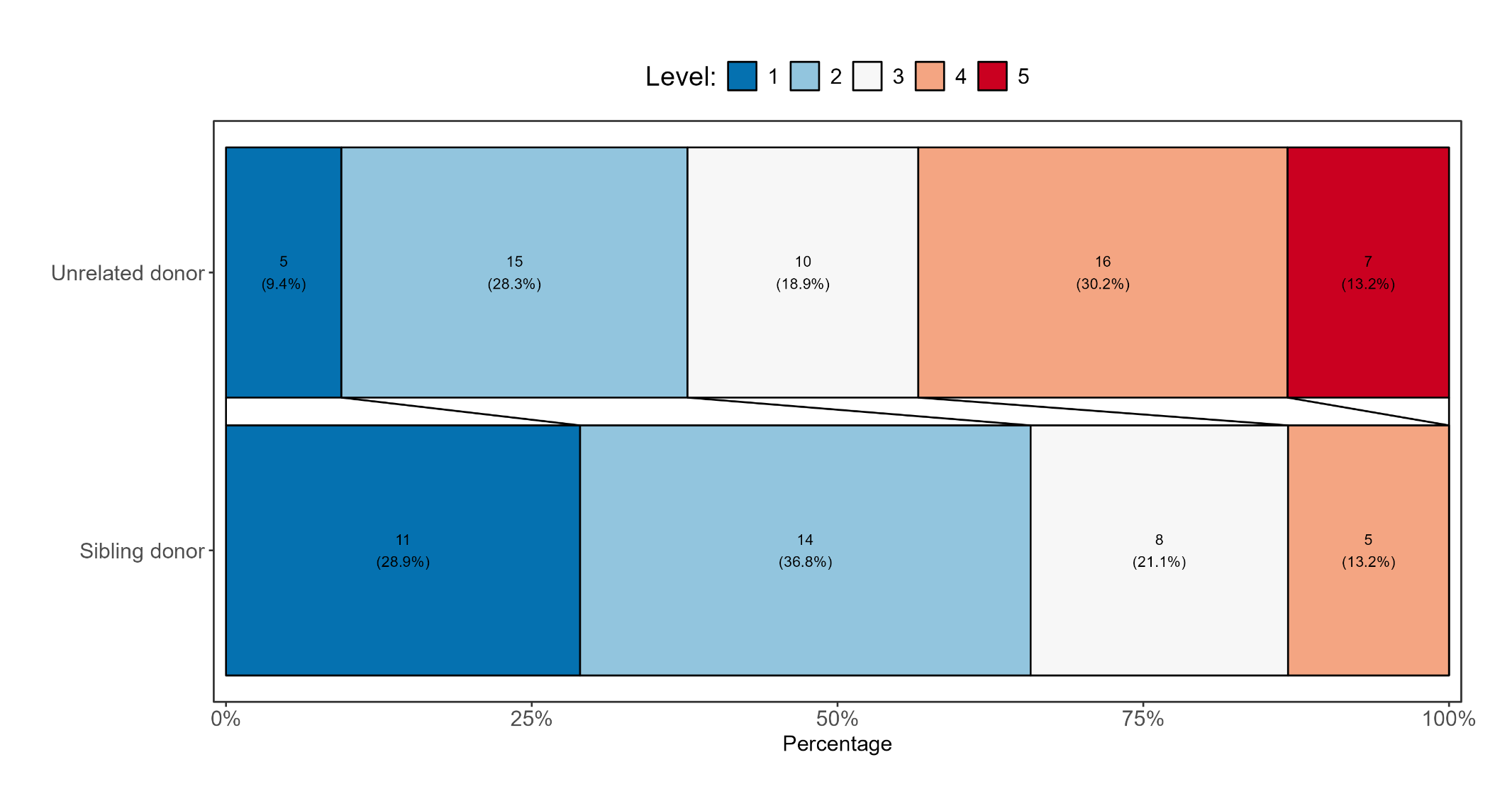
**Figure 13f:** Grotta bar for second ordinal endpoint by transplant type (unrelated vs sibling donor)

**Figure 13g:** Grotta bar for third ordinal endpoint by transplant type (unrelated vs sibling donor)

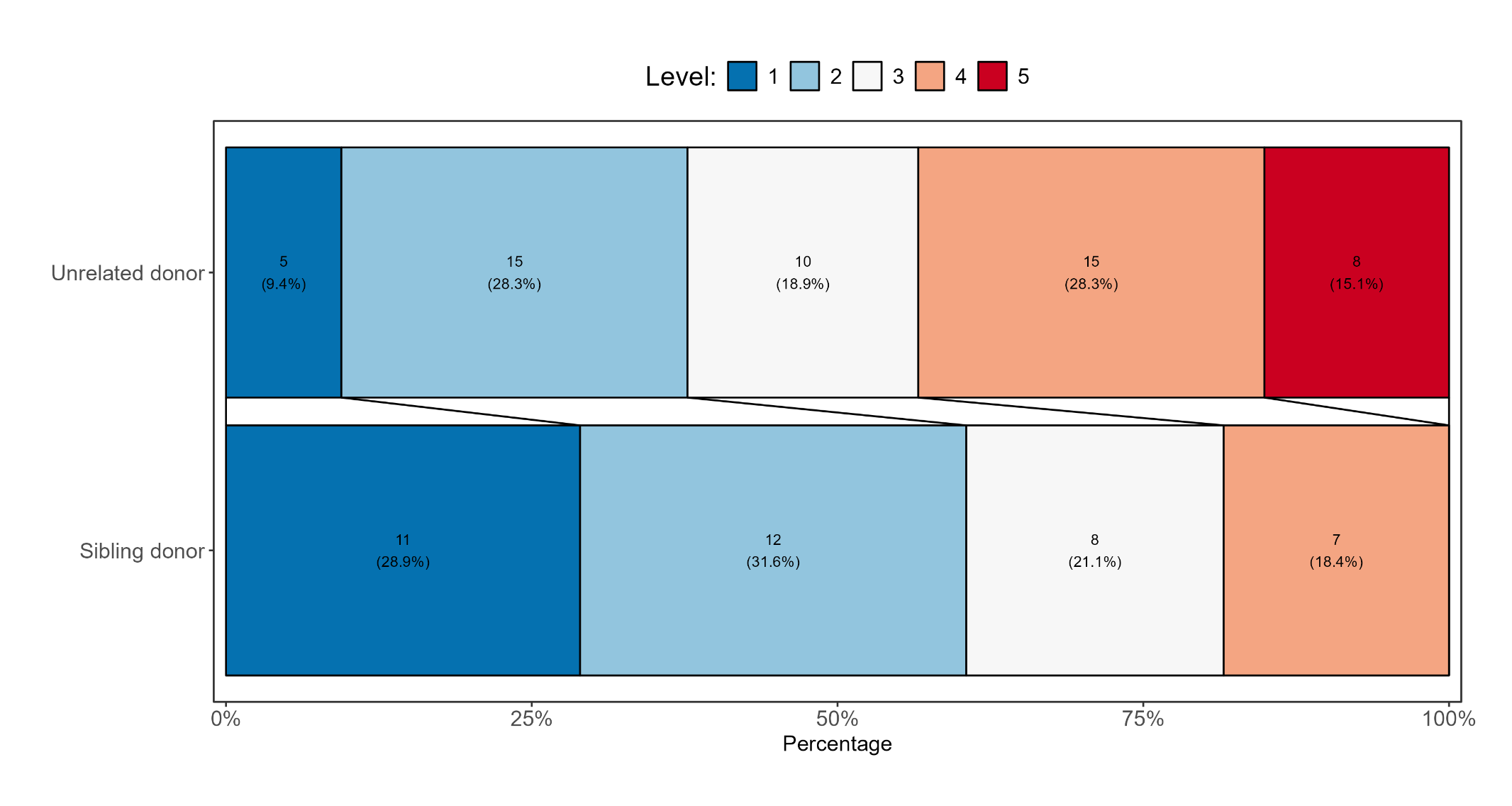

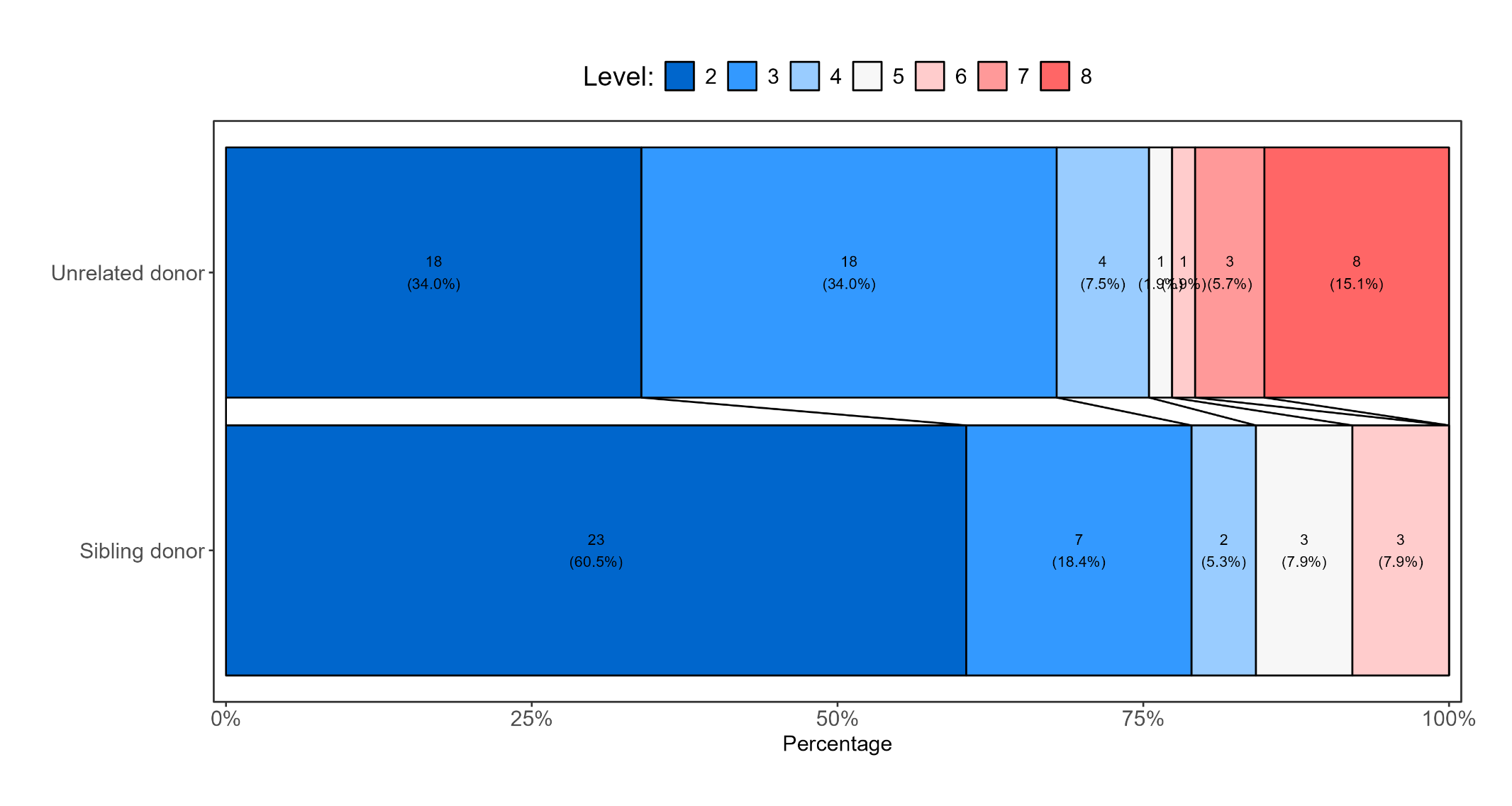
**Figure 13h:** Grotta bar for fourth ordinal endpoint by transplant type (unrelated vs sibling donor)

3. ***Transplant type (for those with non-malignant indications)***

**Figure 14a:** Grotta bar for first ordinal endpoint by transplant type (haploidentical vs sibling donor)

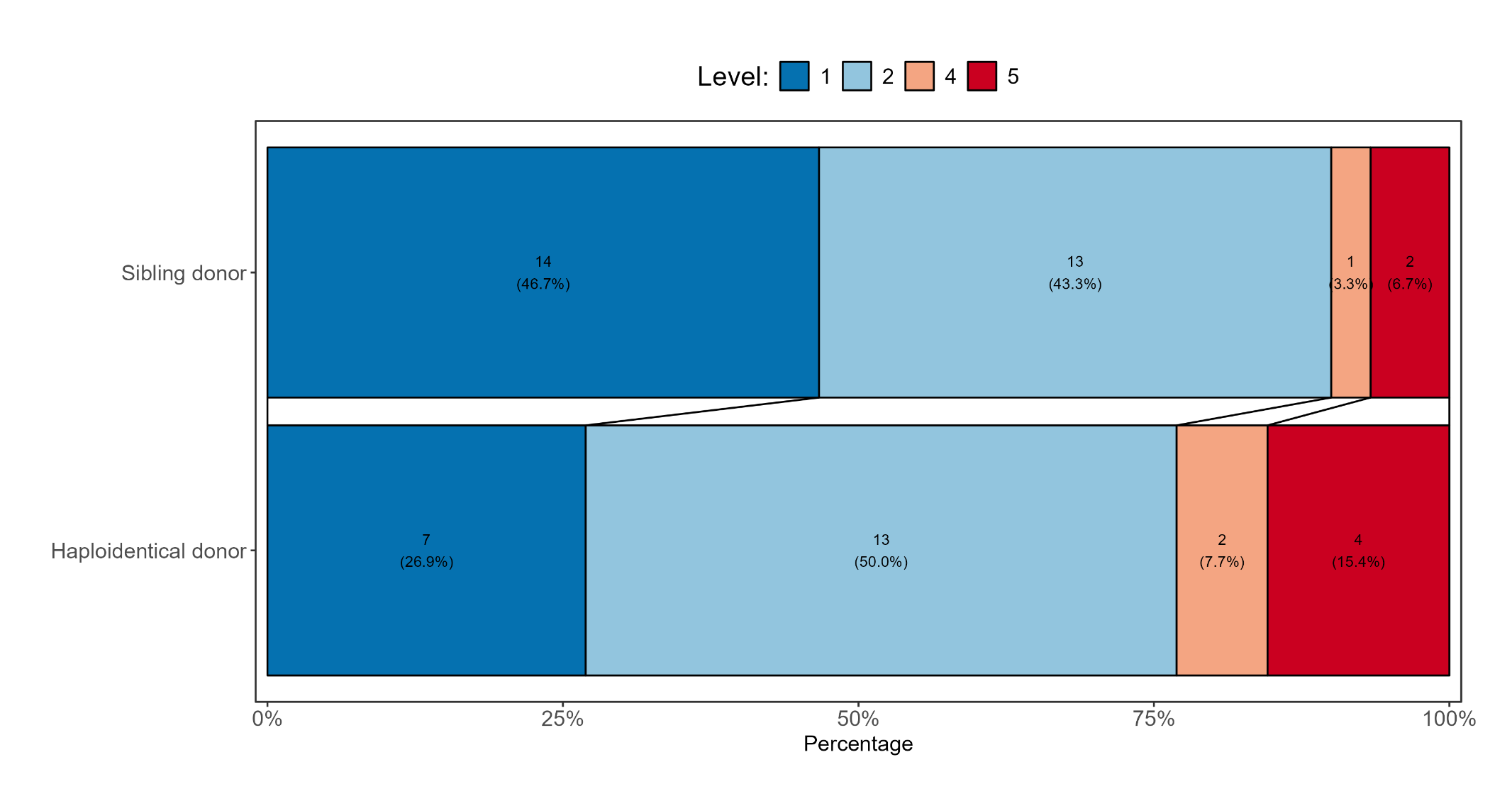

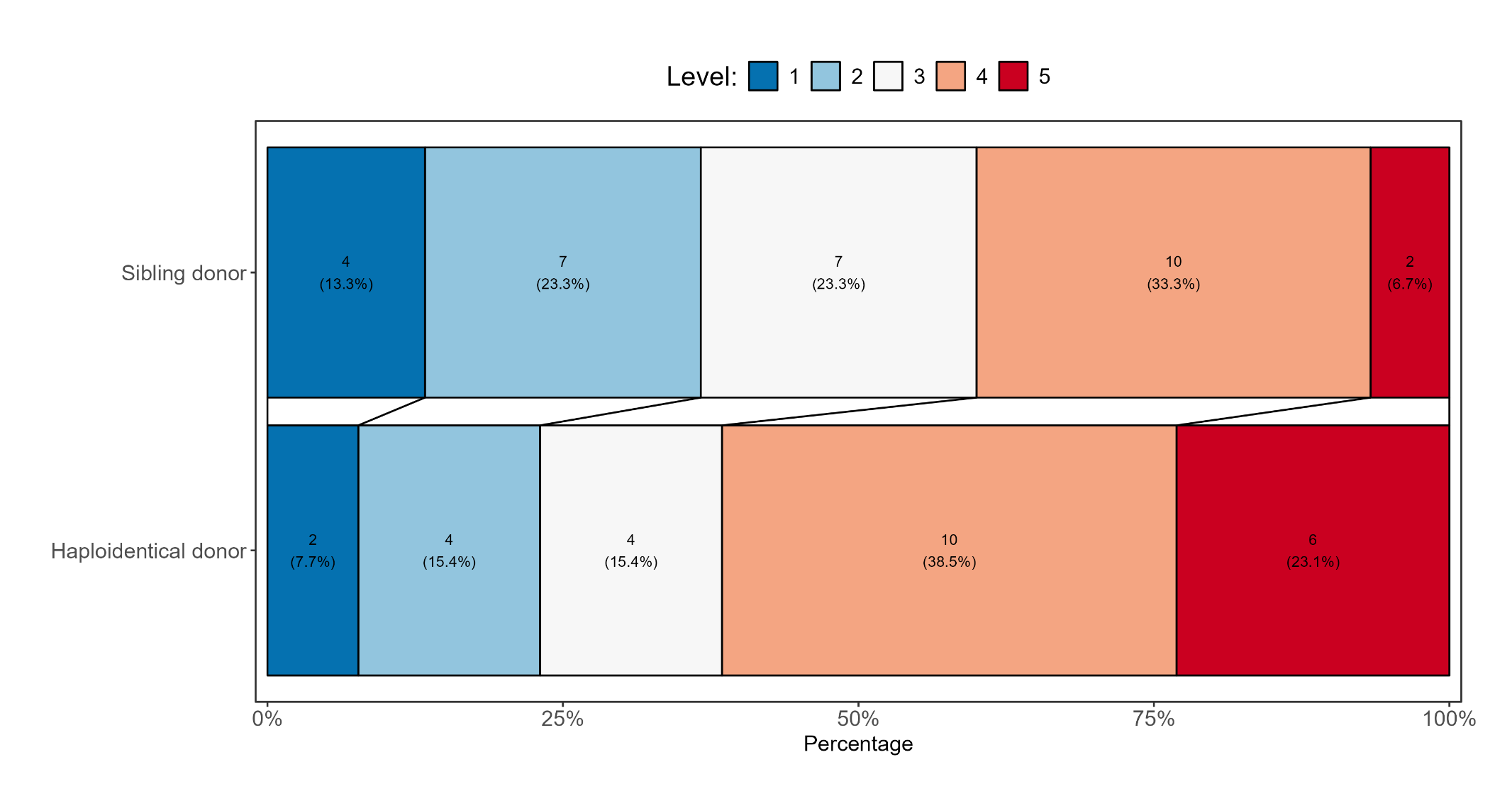
**Figure 14b:** Grotta bar for second ordinal endpoint by transplant type (haploidentical vs sibling donor)

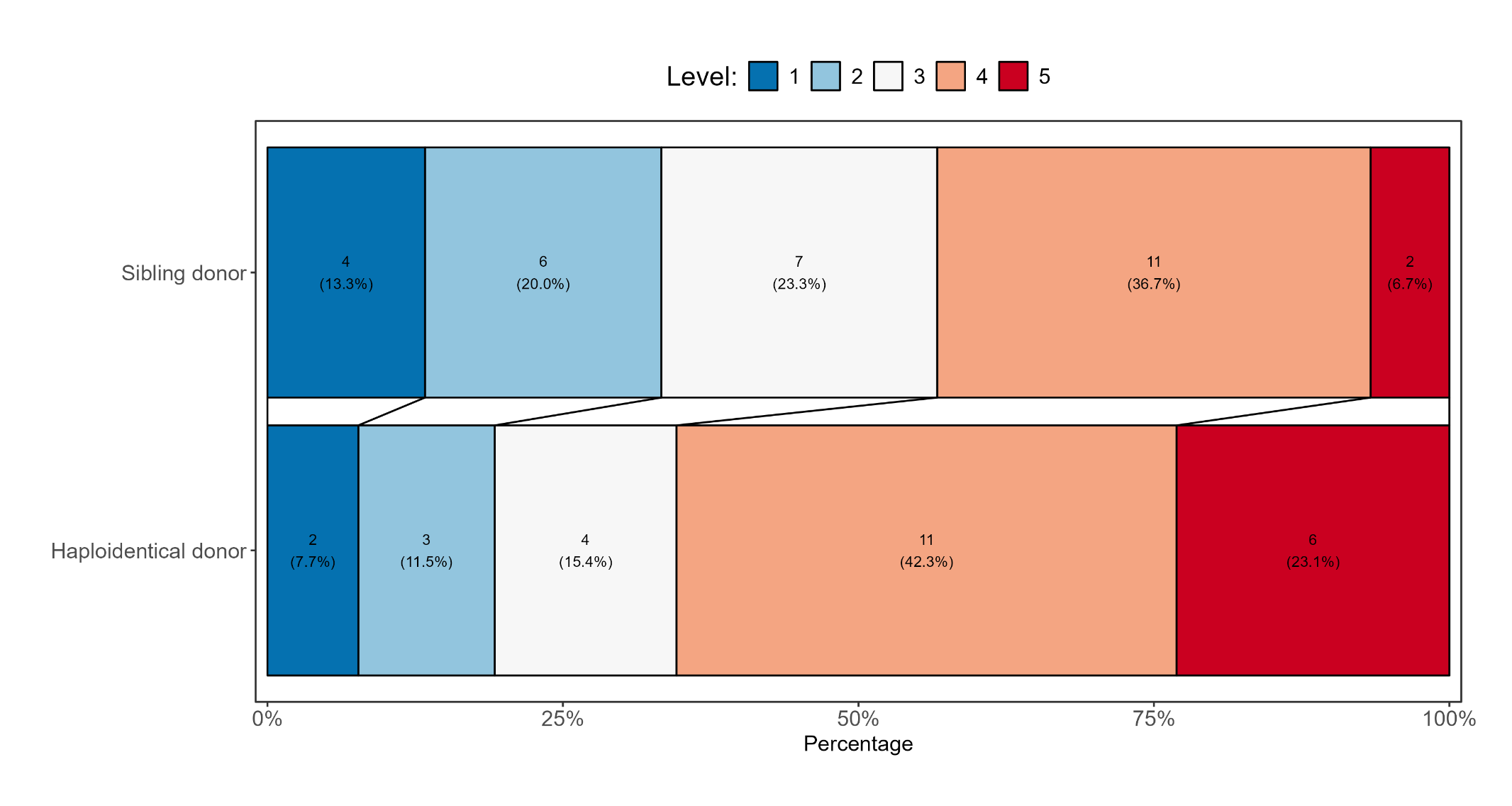
**Figure 14c:** Grotta bar for third ordinal endpoint by transplant type (haploidentical vs sibling donor)

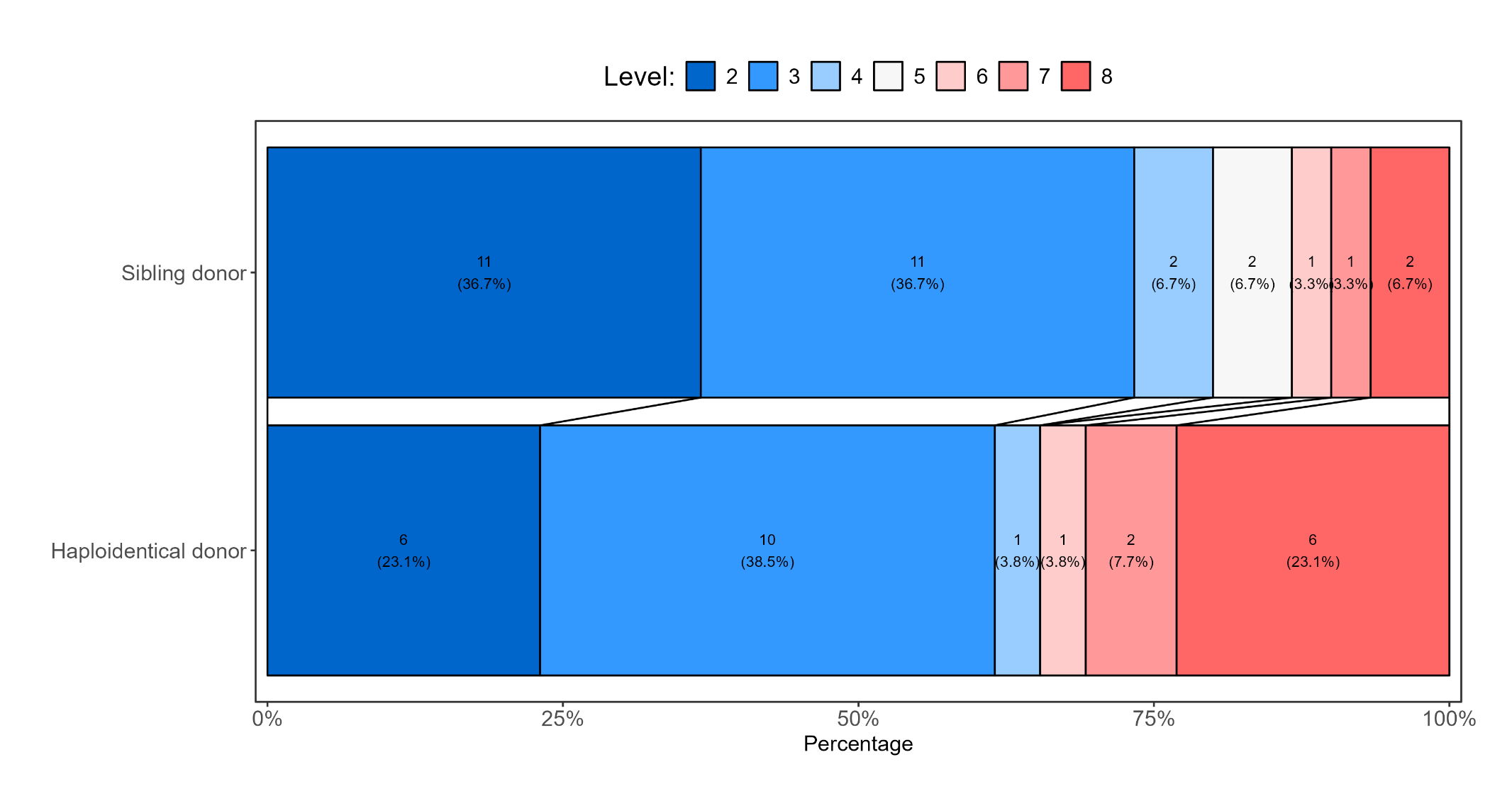
**Figure 14d:** Grotta bar for fourth ordinal endpoint by transplant type (haploidentical vs sibling donor)

**Figure 14e:** Grotta bar for first ordinal endpoint by transplant type (unrelated vs sibling donor)

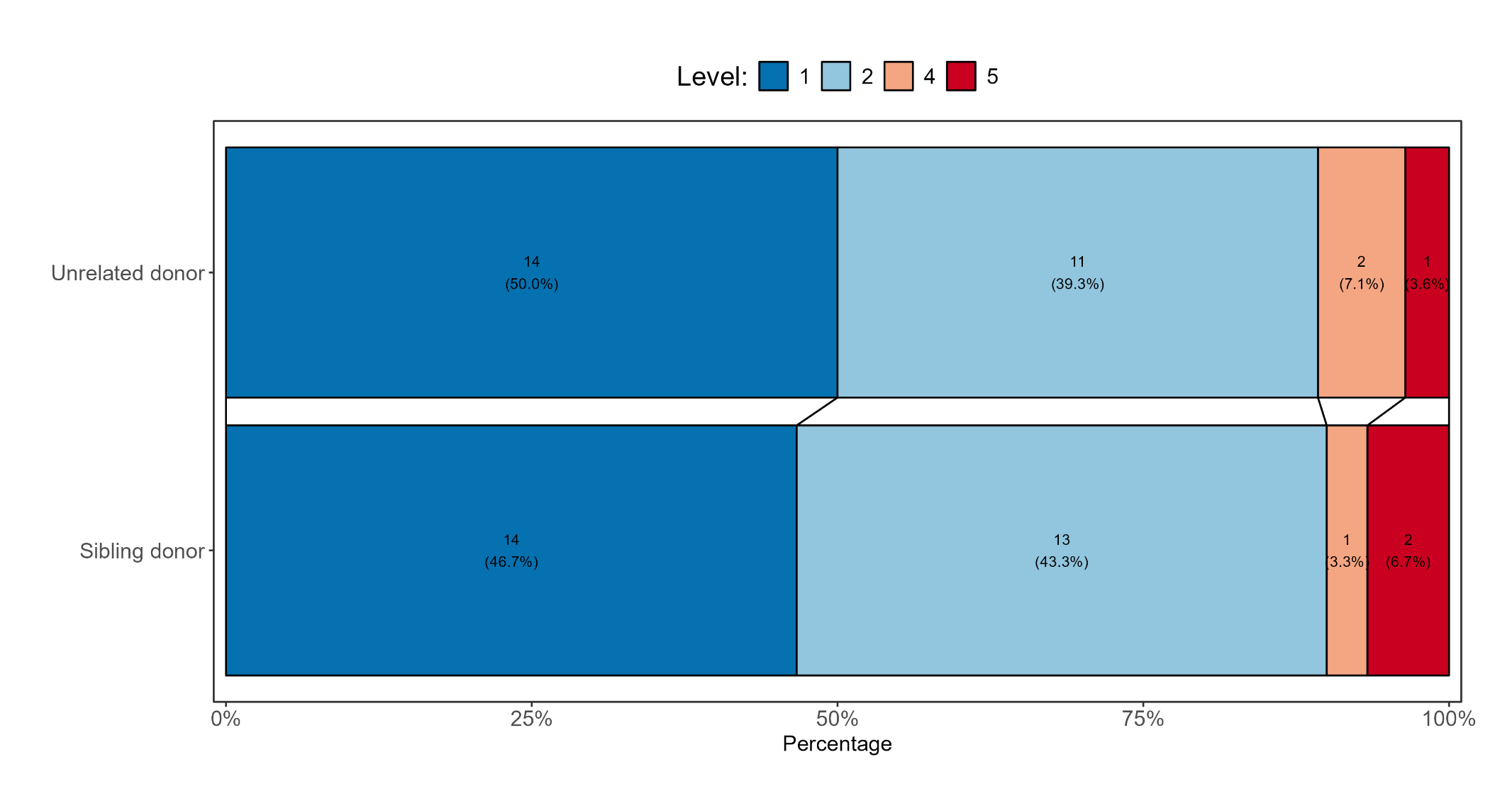

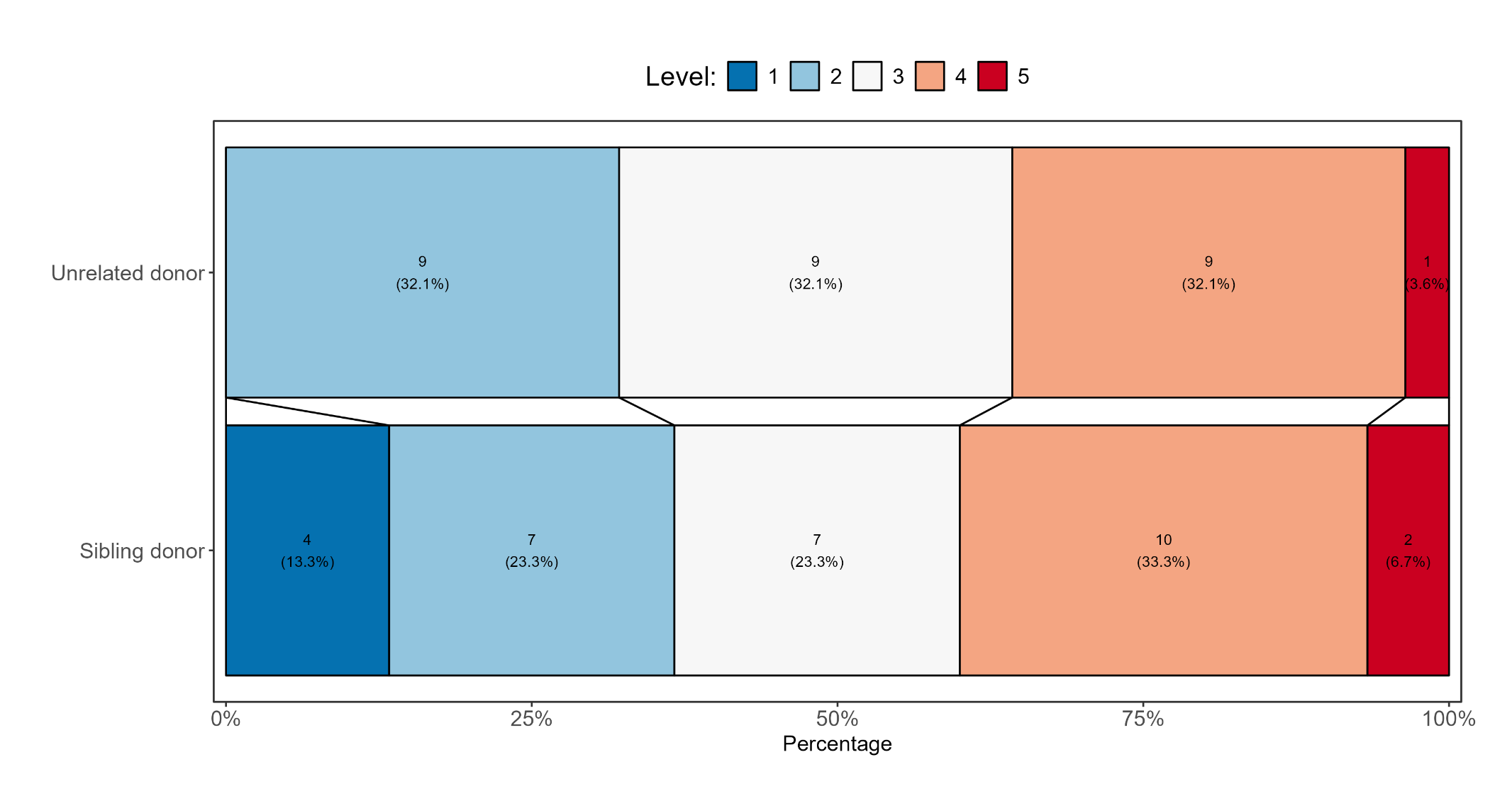
**Figure 14f:** Grotta bar for second ordinal endpoint by transplant type (unrelated vs sibling donor)

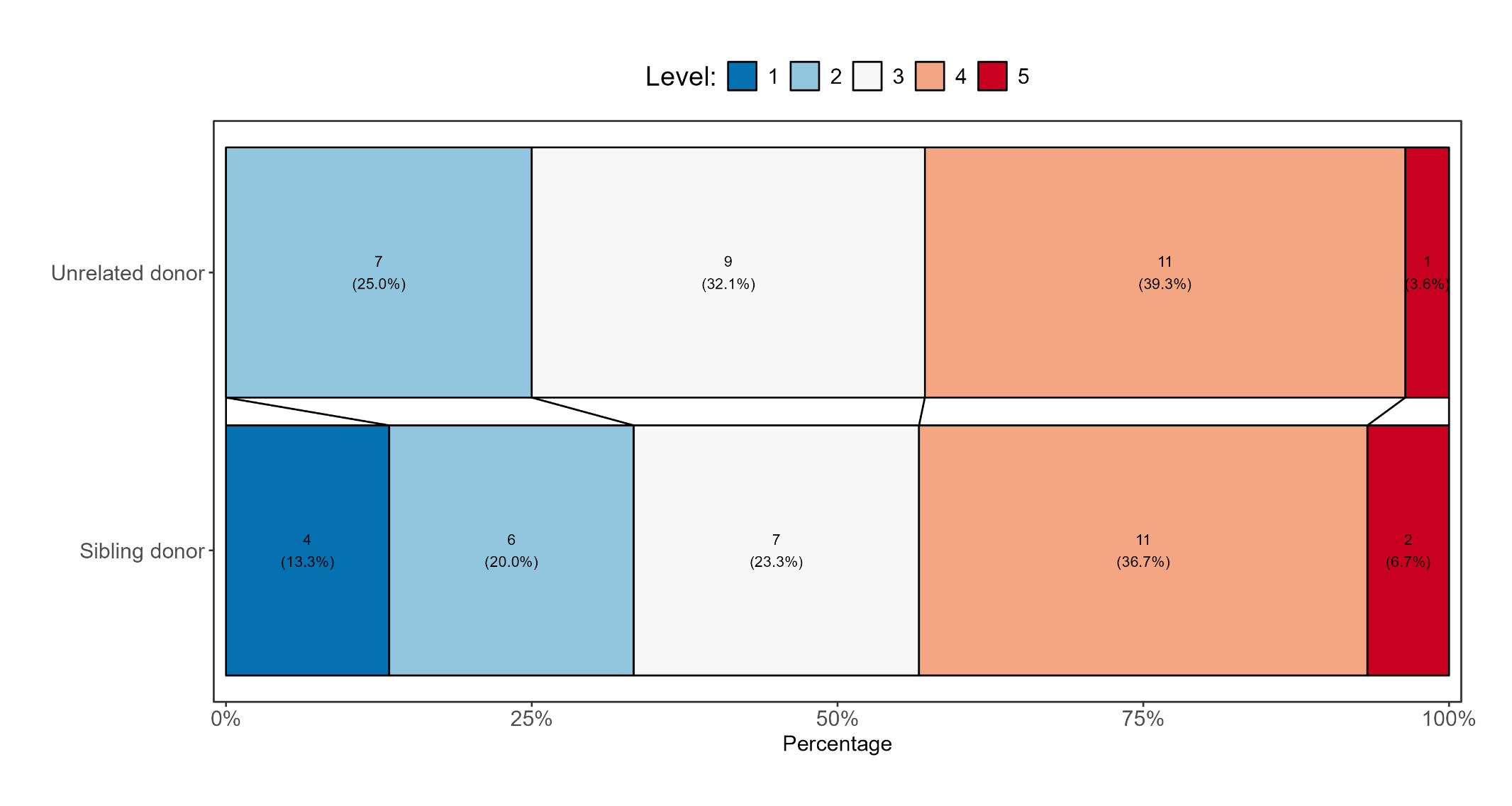
**Figure 14g:** Grotta bar for third ordinal endpoint by transplant type (unrelated vs sibling donor)

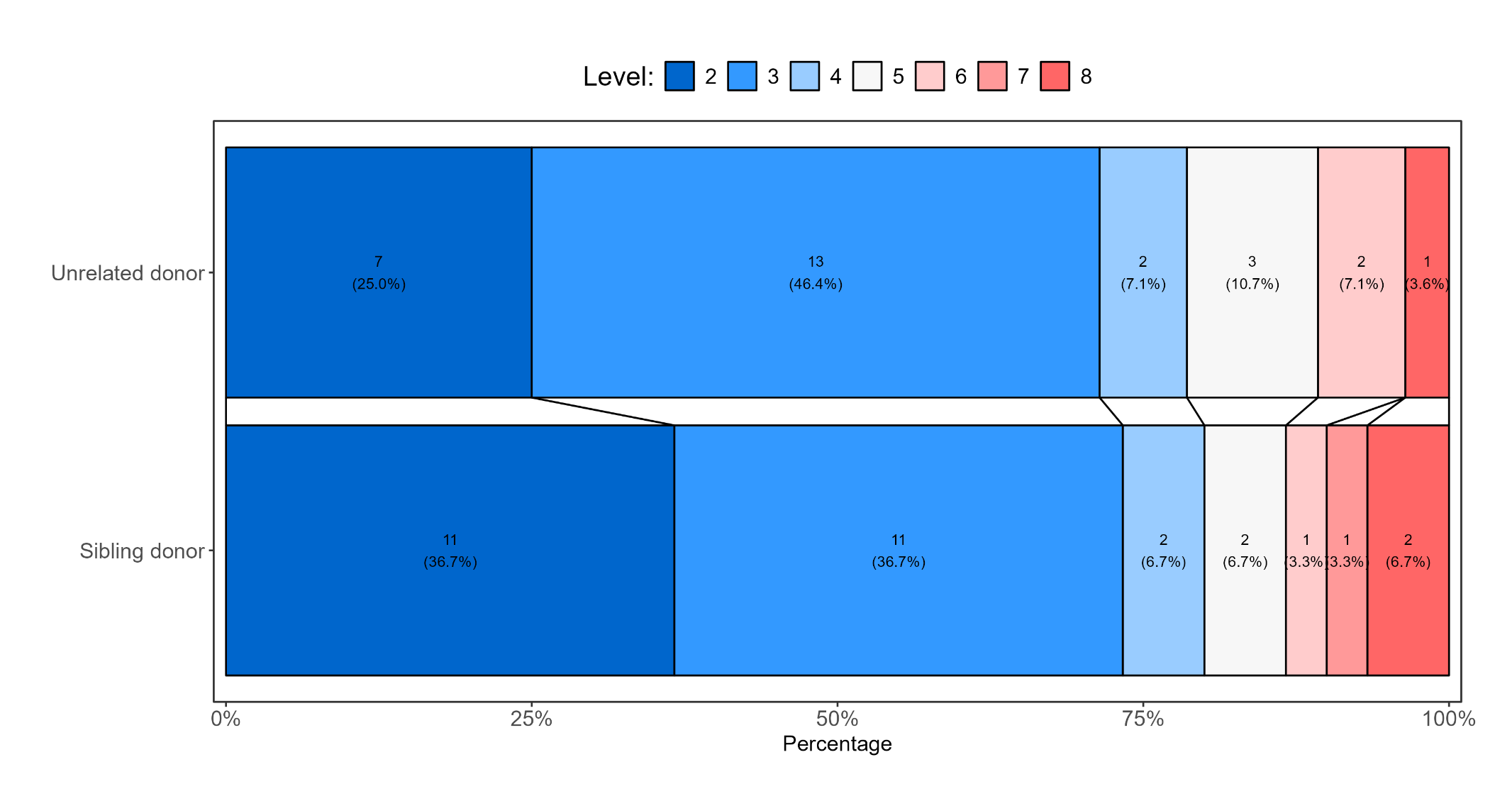
**Figure 14h:** Grotta bar for fourth ordinal endpoint by transplant type (unrelated vs sibling donor)

4. ***Malignant indication for HSCT***

**Figure 15a:** Grotta bar for first ordinal endpoint by malignant indication for HSCT (B-ALL vs AML/MDS)

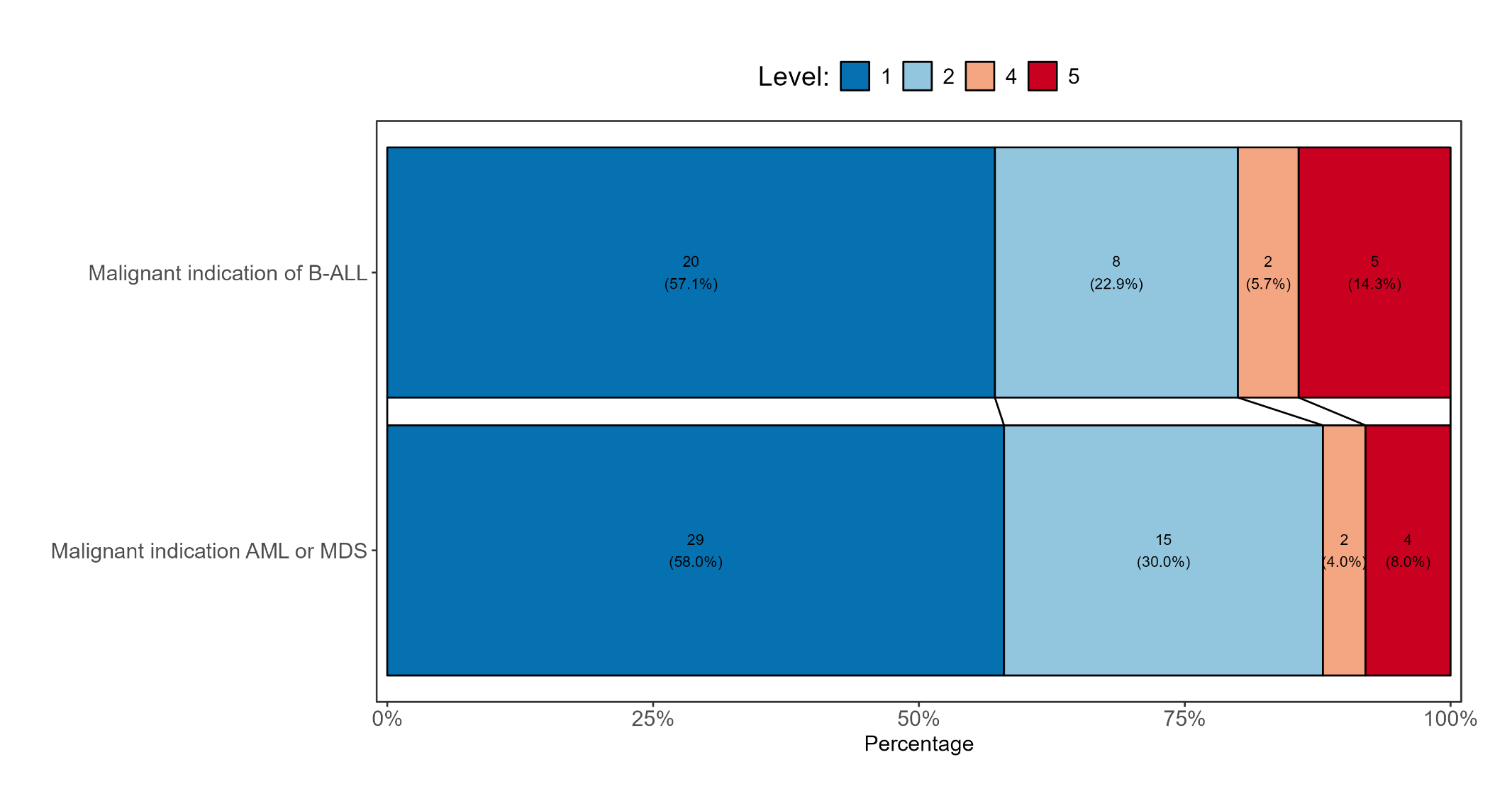

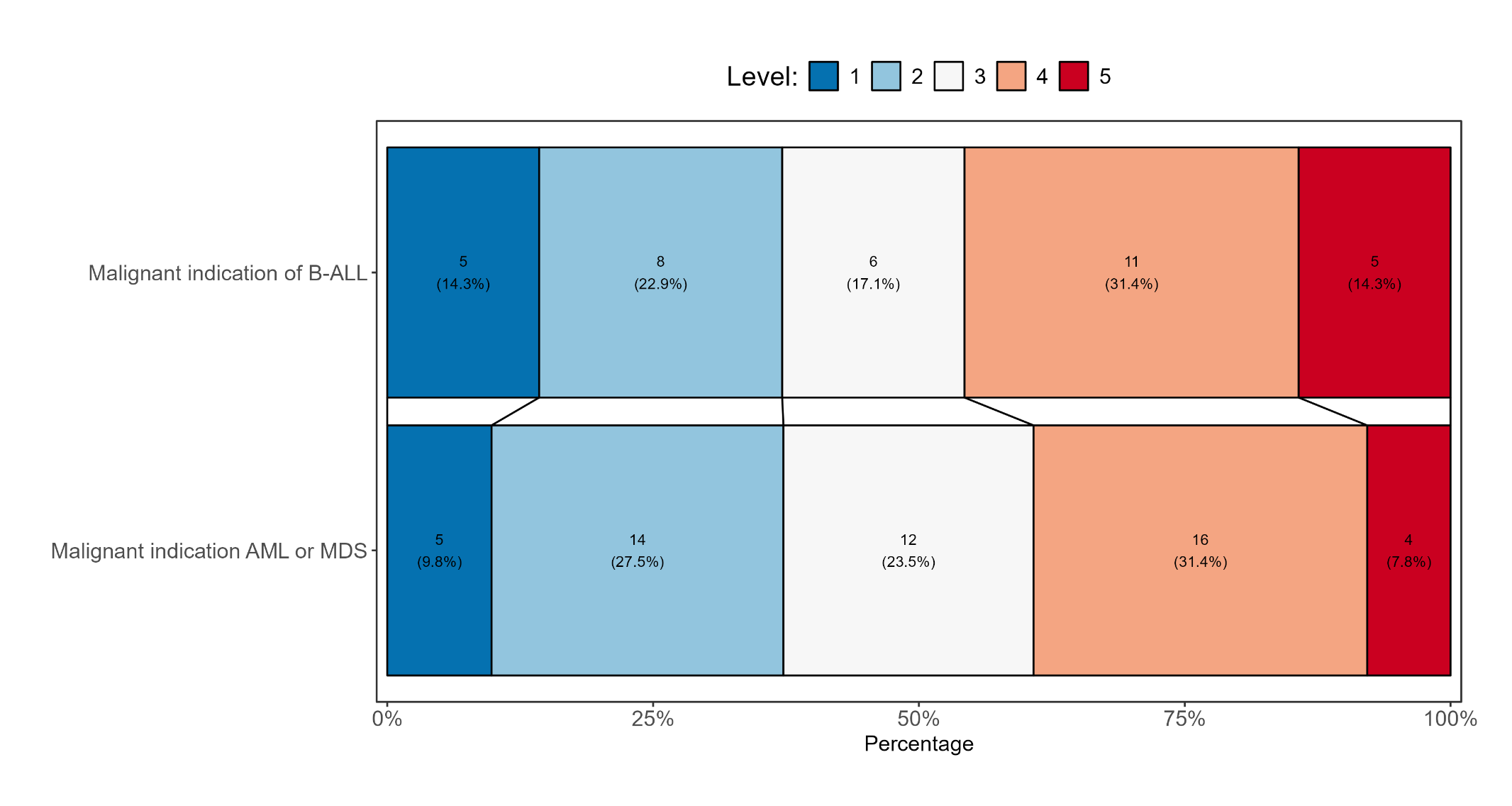
**Figure 15b:** Grotta bar for second ordinal endpoint by malignant indication for HSCT (B-ALL vs AML/MDS)

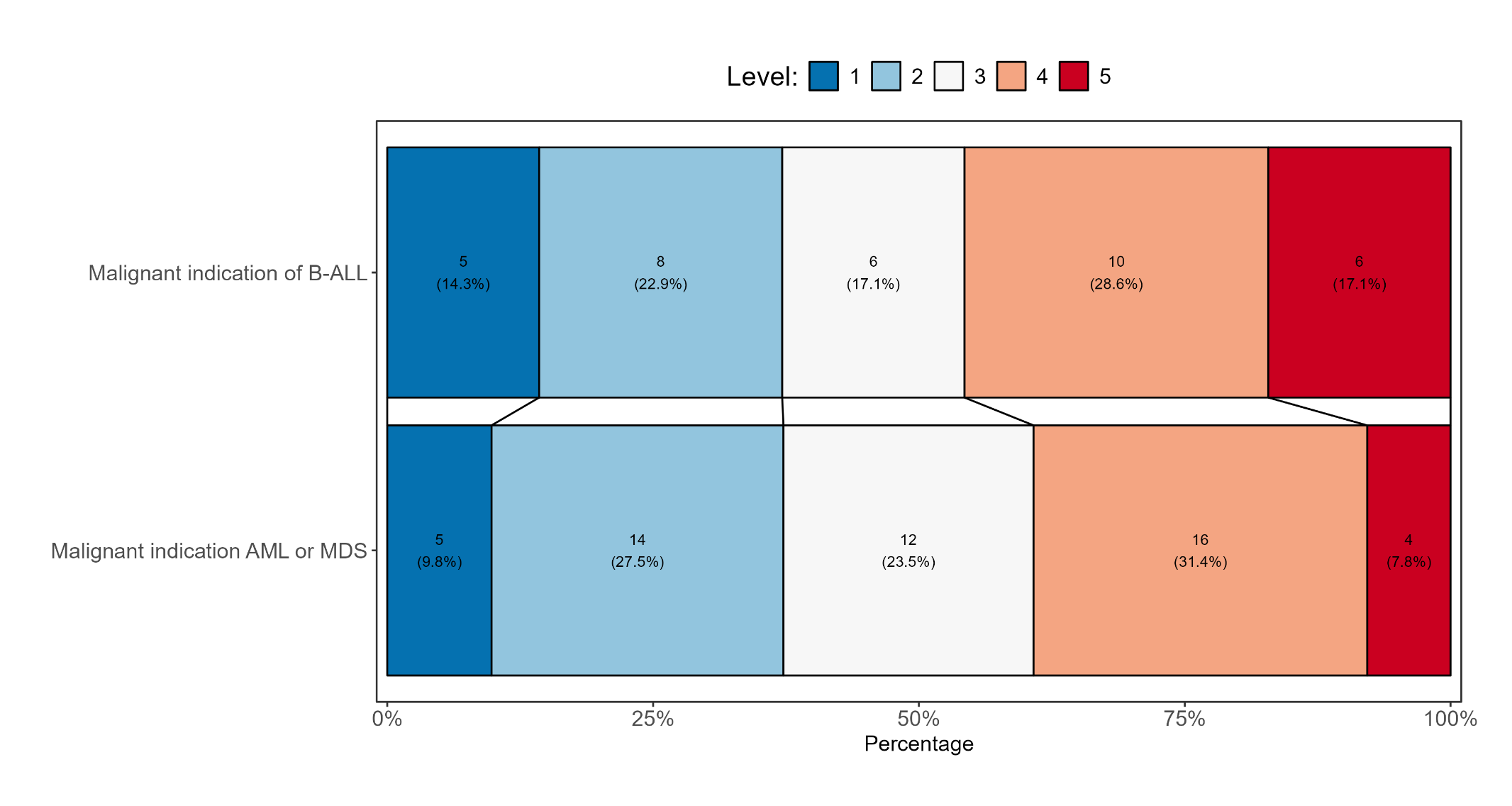
**Figure 15c:** Grotta bar for third ordinal endpoint by malignant indication for HSCT (B-ALL vs AML/MDS)

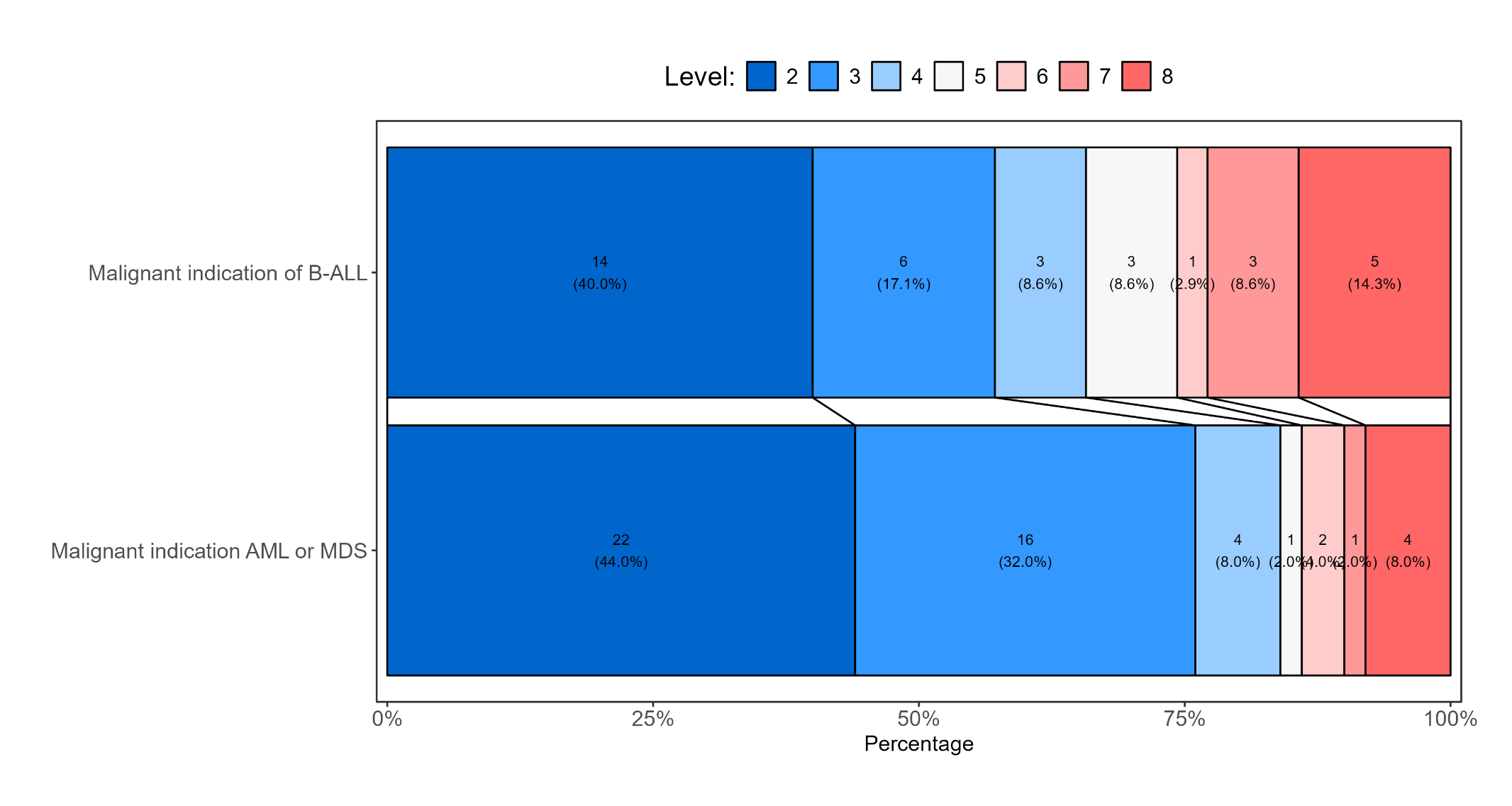
**Figure 15d:** Grotta bar for fourth ordinal endpoint by malignant indication for HSCT (B-ALL vs AML/MDS)

5. ***Cord blood transplant vs other (bone marrow or peripheral blood stem cell)***

**Figure 16a:** Grotta bar for first ordinal endpoint by cord blood transplant vs other transplant

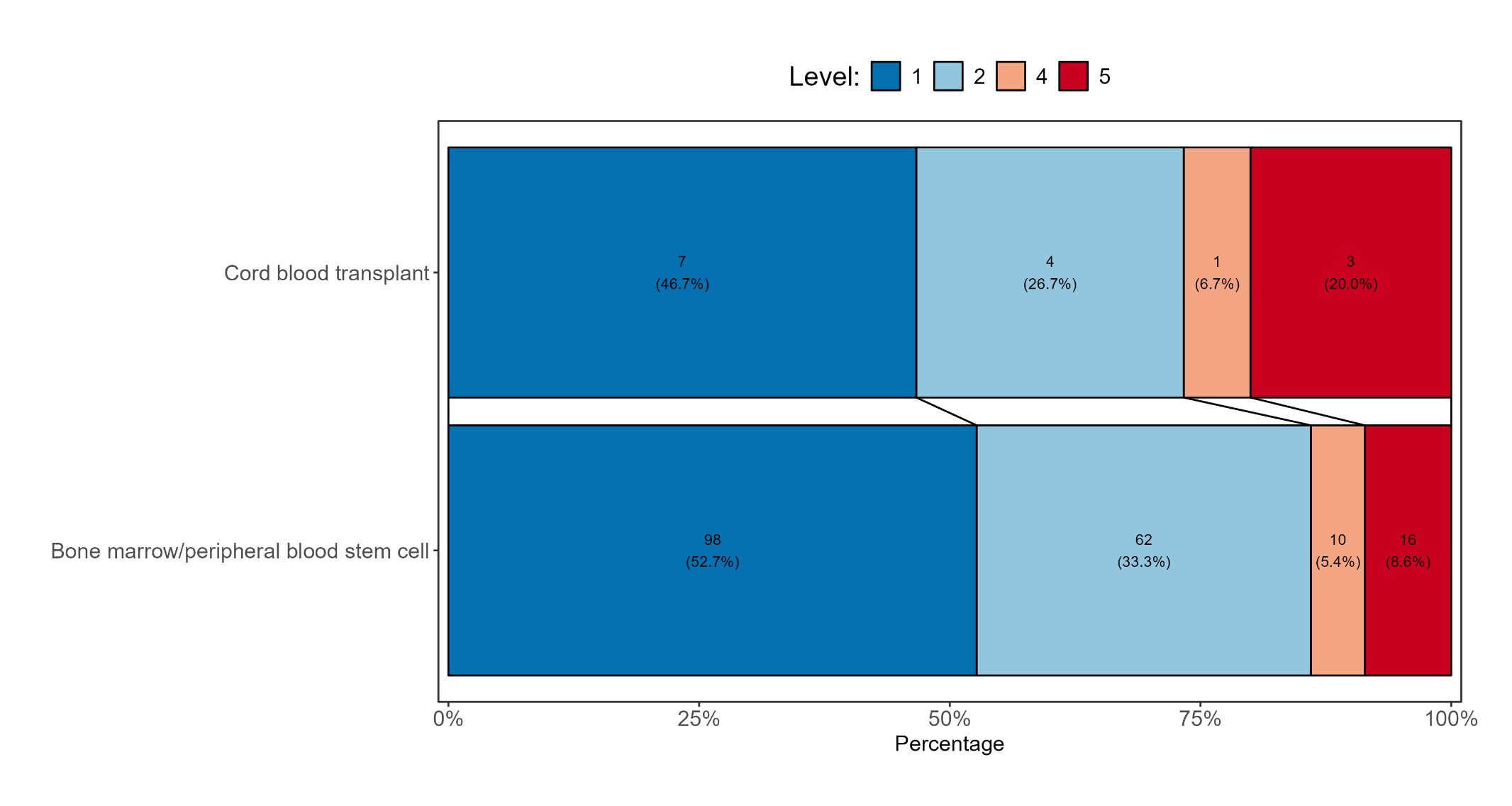

**Figure 16b:** Grotta bar for second ordinal endpoint by cord blood transplant vs other transplant

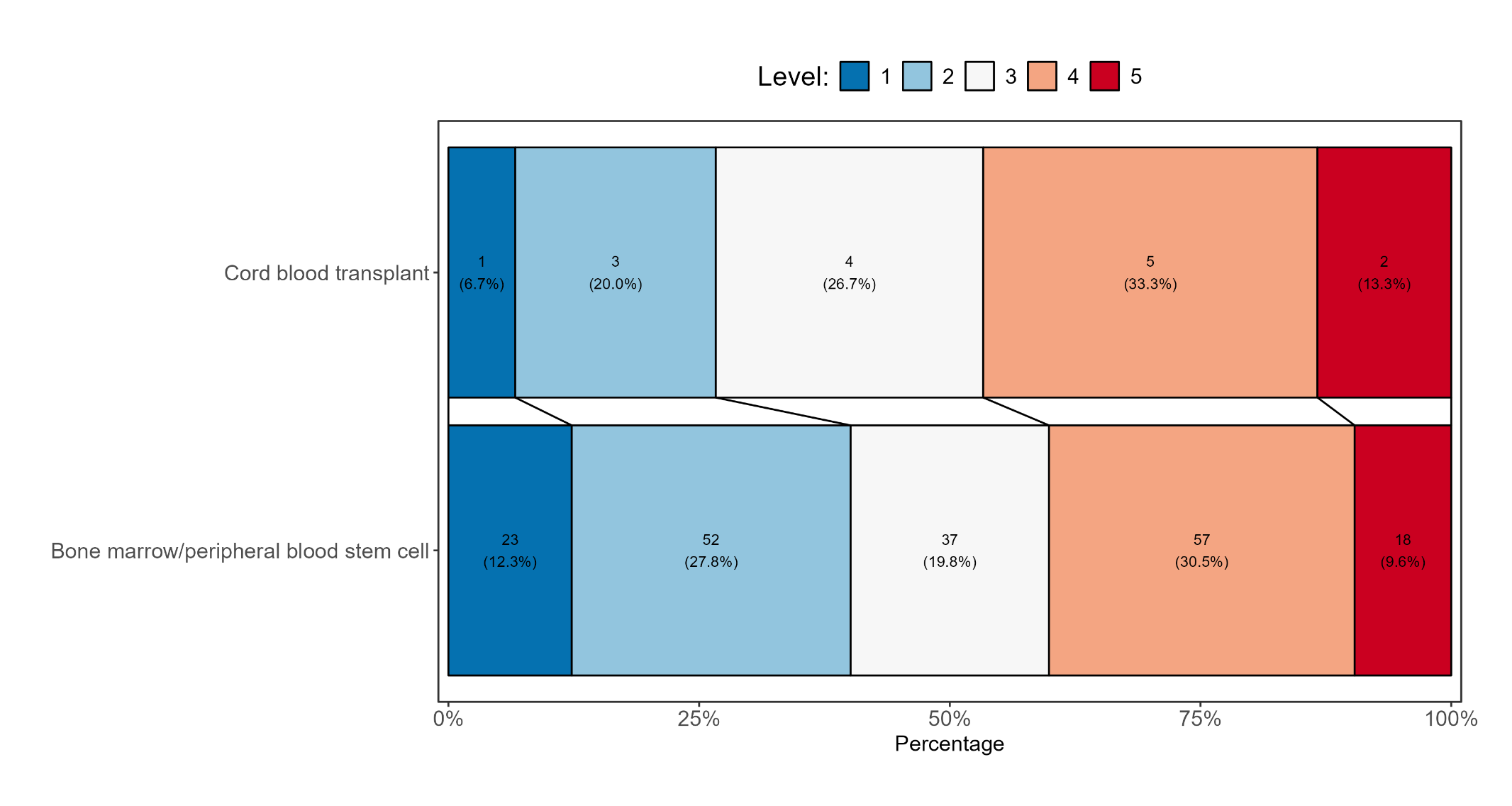

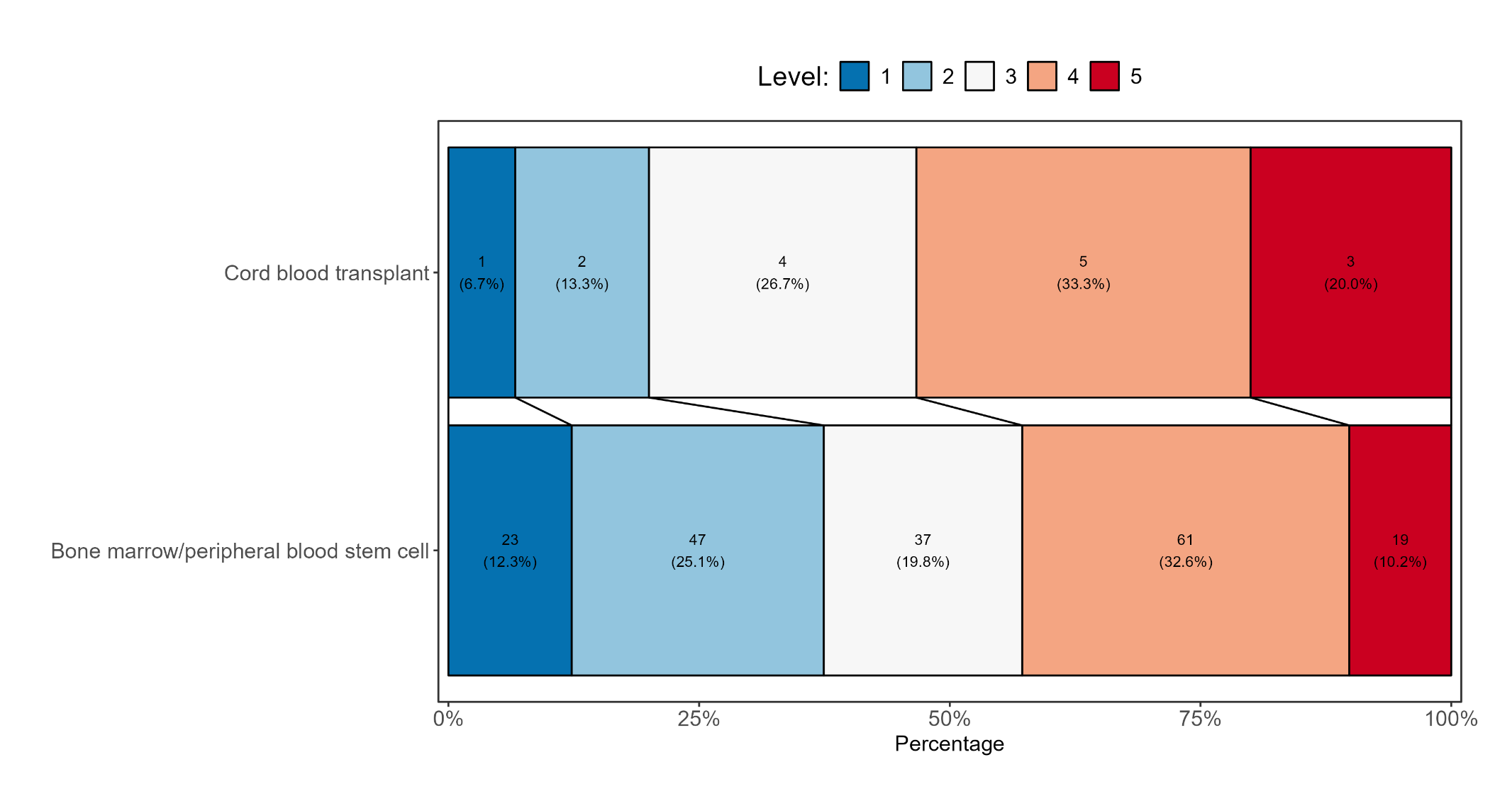
**Figure 16c:** Grotta bar for third ordinal endpoint by cord blood transplant vs other transplant

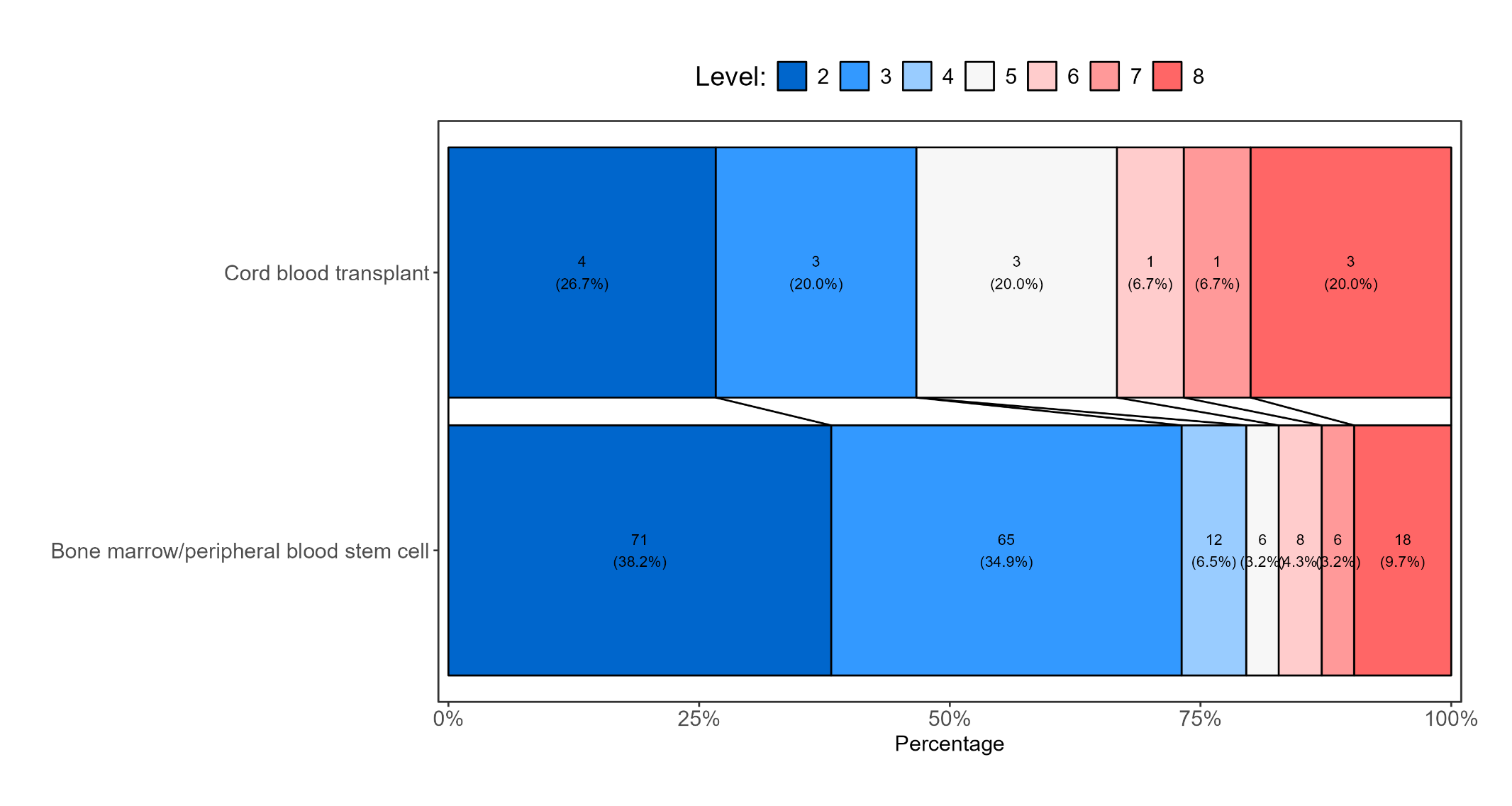
**Figure 16d:** Grotta bar for fourth ordinal endpoint by cord blood transplant vs other transplant

6. **Patients with B-ALL: comparing those with MRD or no MRD prior to transplant**

**Figure 17a:** Grotta bar for first ordinal endpoint by whether patient had MRD or not among those with B-ALL

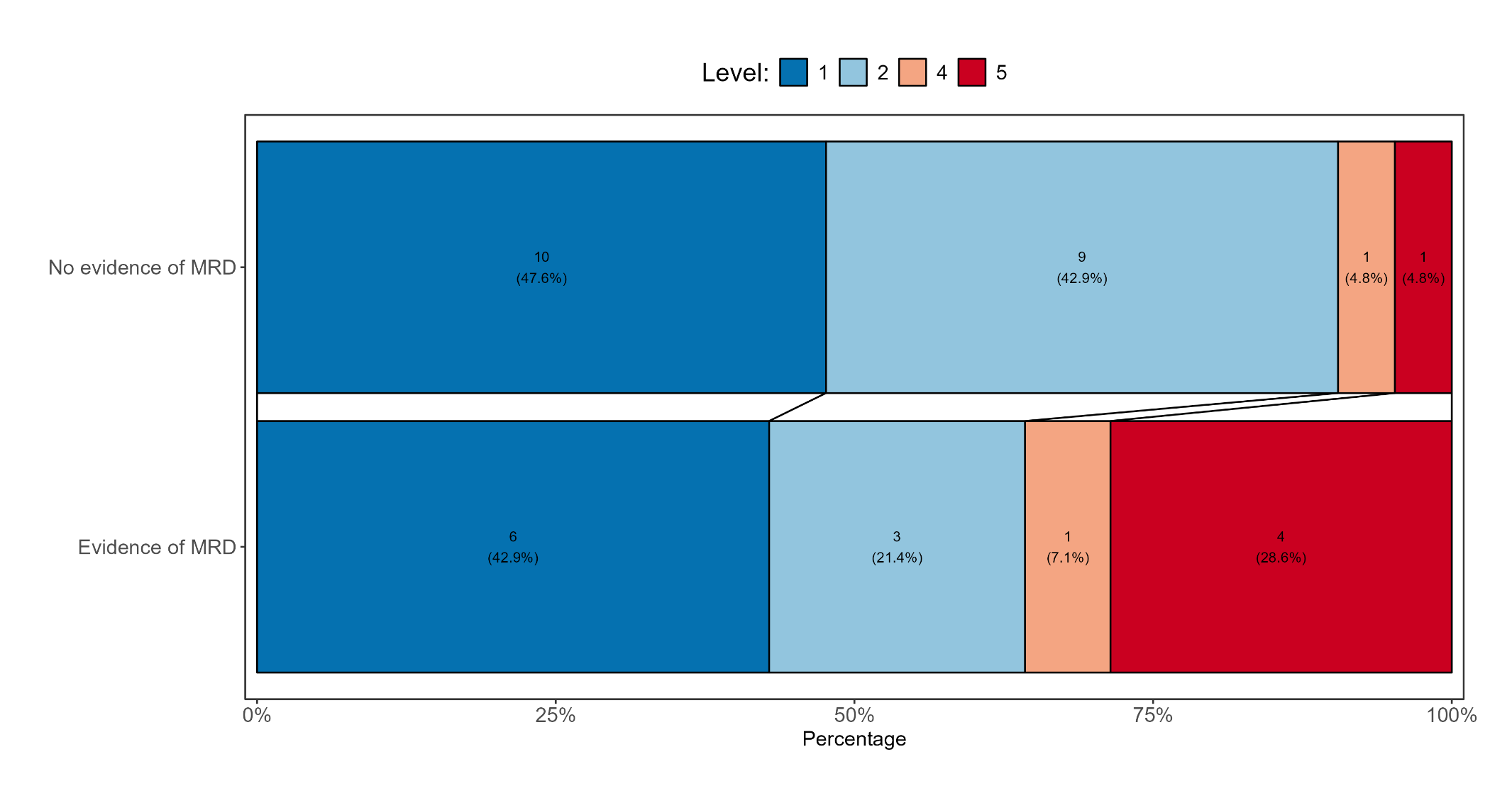

**Figure 17b:** Grotta bar for second ordinal endpoint by whether patient had MRD or not among those with B-ALL

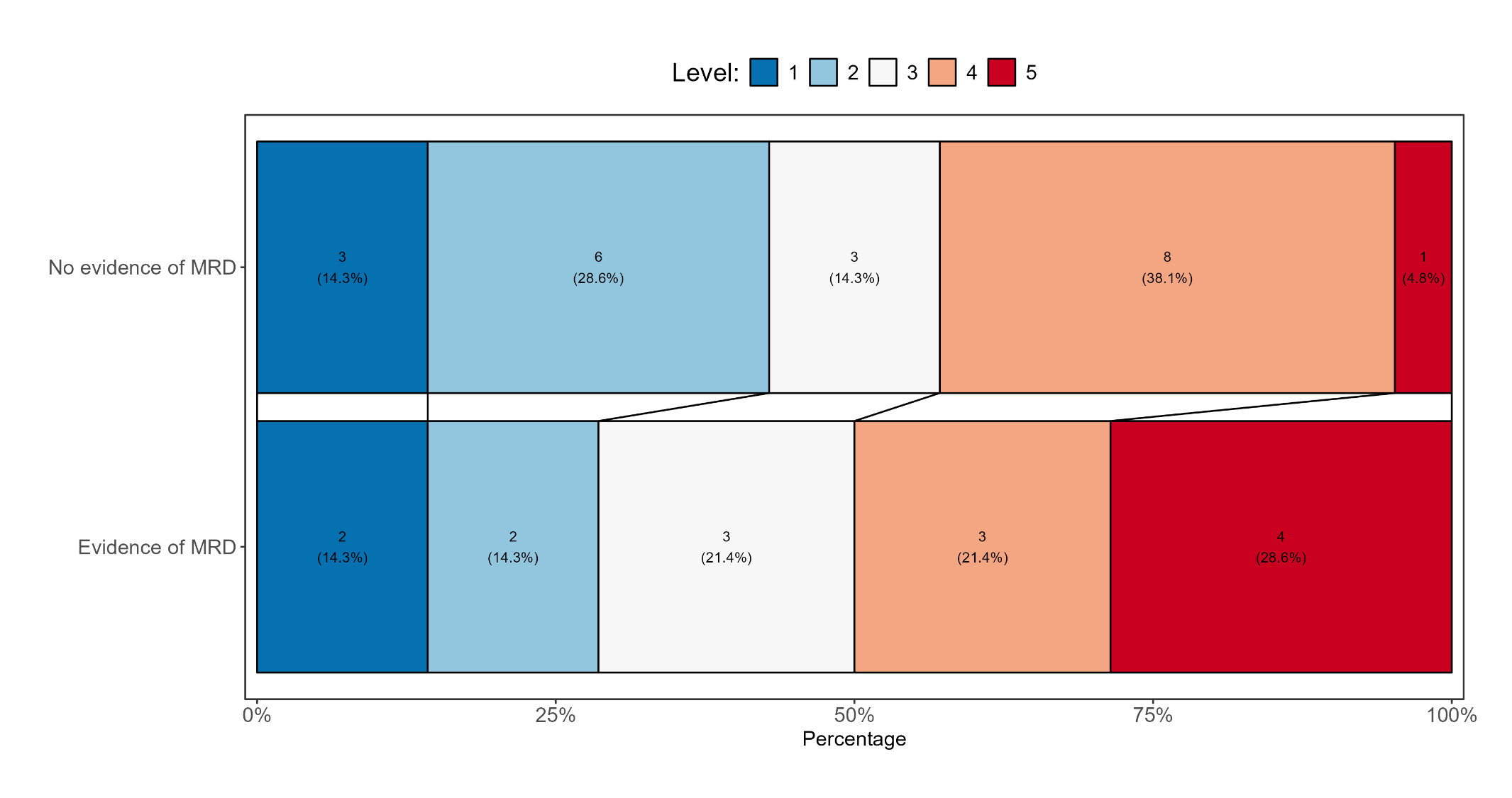

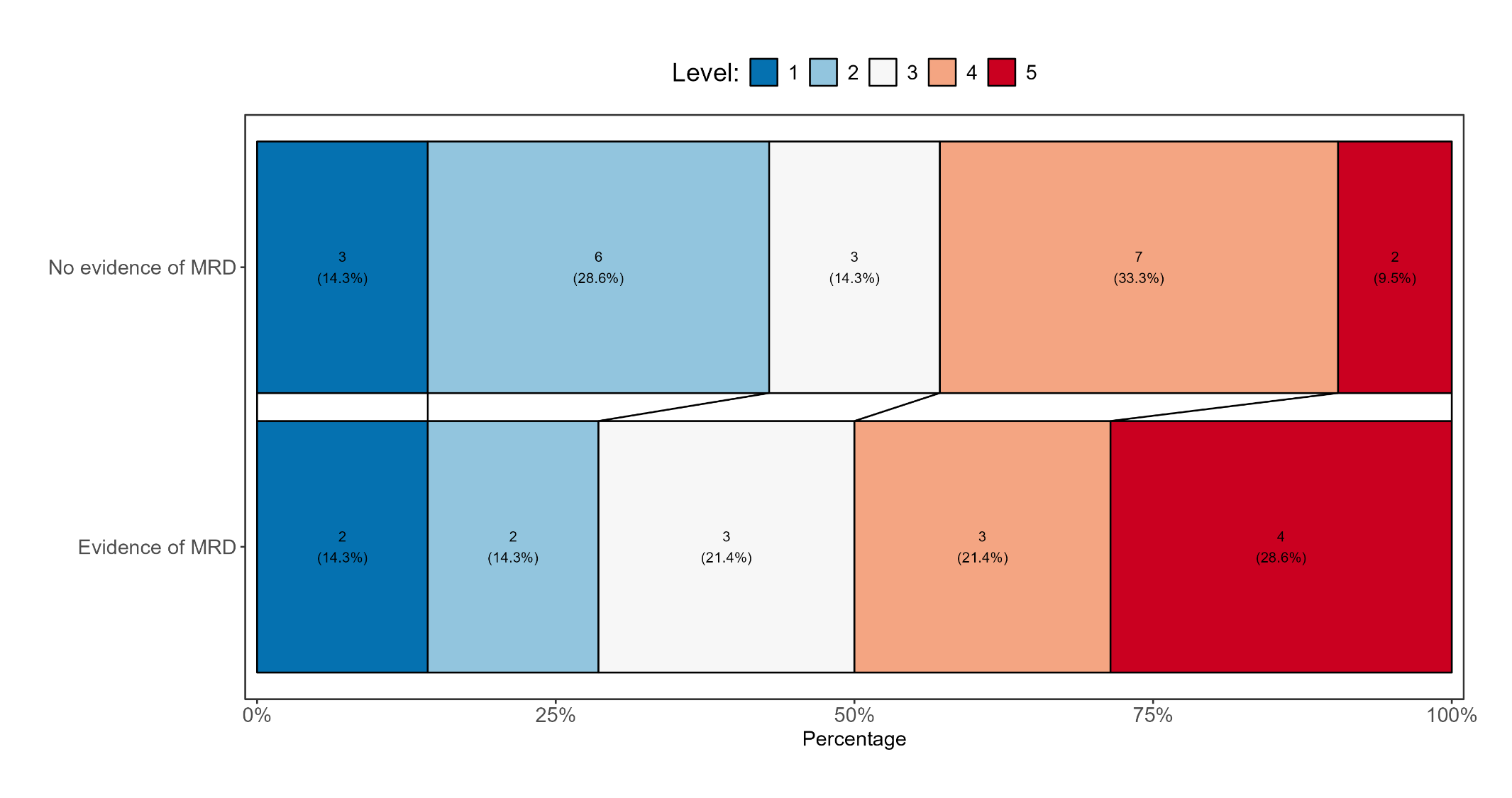
**Figure 17c:** Grotta bar for third ordinal endpoint by whether patient had MRD or not among those with B-ALL

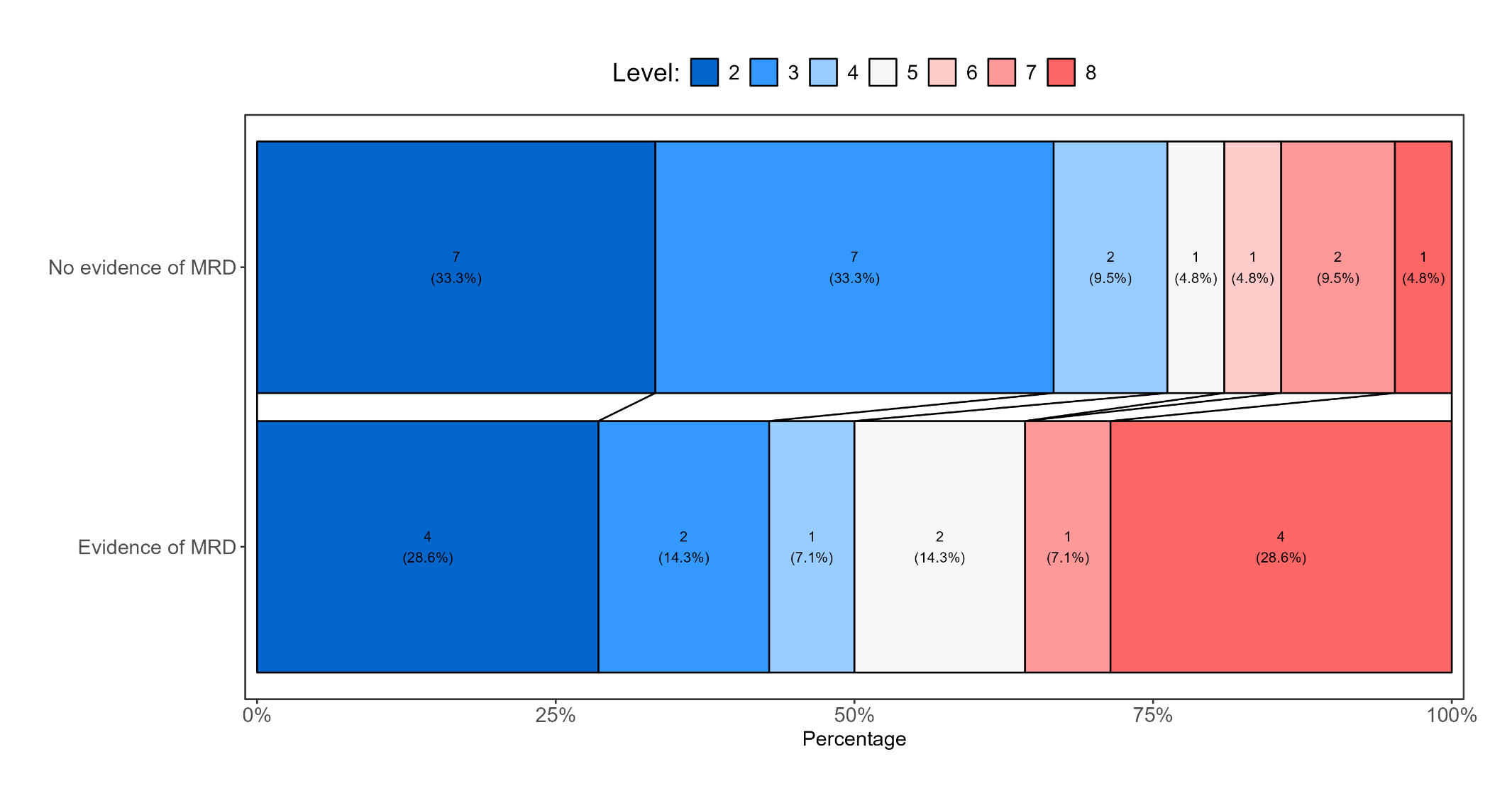
**Figure 17d:** Grotta bar for fourth ordinal endpoint by whether patient had MRD or not among those with B-ALL
